# Multi-omics analysis reveals drivers of loss of β-cell function after newly diagnosed autoimmune type 1 diabetes: An INNODIA^‡^ multicenter study

**DOI:** 10.1101/2023.03.22.23287261

**Authors:** Jose Juan Almagro Armenteros, Caroline Brorsson, Christian Holm Johansen, Karina Banasik, Gianluca Mazzoni, Robert Moulder, Karoliina Hirvonen, Tomi Suomi, Omid Rasool, Sylvaine FA Bruggraber, M Loredana Marcovecchio, Emile Hendricks, Naba Al-Sari, Ismo Mattila, Cristina Legido-Quigley, Tommi Suvitaival, Piotr J Chmura, Mikael Knip, Anke M Schulte, Jeong Heon Lee, Guido Sebastiani, Giuseppina Emanuela Grieco, Laura L Elo, Simranjeet Kaur, Flemming Pociot, Francesco Dotta, Tim Tree, Riitta Lahesmaa, Lut Overbergh, Chantal Mathieu, Mark Peakman, Søren Brunak, the INNODIA‡ investigators

## Abstract

**Background:** Heterogeneity in the rate of β-cell loss in newly diagnosed type 1 diabetes patients is poorly understood and creates a barrier to designing and interpreting disease-modifying clinical trials. Integrative analyses of complementary multi-omics data obtained after the diagnosis of T1D may provide mechanistic insight into the diverse rates of disease progression.

**Methods:** We collected samples in a pan-European consortium that enabled the concerted analysis of five different omics modalities in data from 97 newly diagnosed patients. In this study we used Multi-Omics Factor Analysis to identify molecular signatures correlating with post-diagnosis decline in β-cell mass measured as fasting C-peptide.

**Results:** Two molecular signatures were significantly correlated with fasting C-peptide levels. One signature showed a correlation to neutrophil degranulation, cytokine signaling, lymphoid and non-lymphoid cell interactions and G-protein coupled receptor signaling events that were inversely associated with rapid decline in β-cell function. The second signature was related to translation and viral infection were inversely associated with change in β-cell function. In addition, the immunomics data revealed a Natural Killer cell signature associated with rapid β-cell decline.

**Conclusion:** Features that differ between individuals with slow and rapid decline in β-cell mass could be valuable in staging and prediction of the rate of disease progression and thus enable smarter (shorter and smaller) trial designs for disease modifying therapies, as well as offering biomarkers of therapeutic effect.

**Funding:** This work is funded by the Innovative Medicine Initiative 2 Joint Undertaking (IMI2 JU) under grant agreement N° 115797 (INNODIA) and N° 945268 (INNODIA HARVEST). This Joint Undertaking receives support from the Union’s Horizon 2020 research and innovation program and ‘EFPIA’, ‘JDRF’ and ‘The Leona M. and Harry B. Helmsley Charitable Trust’.

## Introduction

Type 1 diabetes is an autoimmune disease involving environmental and genetic factors that trigger immune-mediated pancreatic β-cell dysfunction and destruction that results in insulin loss and symptomatic hyperglycemia requiring lifelong insulin therapy^1^. Globally, around 1.2 million people below the age of 20 years have type 1 diabetes, with an annually increasing incidence of 3% influenced strongly by geography^2^. Insulin replacement therapy is unable to fully mimic physiological control of blood glucose and therefore, many people living with type 1 diabetes develop severe disease complications that are directly attributable to prolonged glycemic exposure^3^, with markedly reduced life expectancy^4^. In the face of the disease burden and unmet need, international consortia are mobilizing to develop disease-modifying therapies. For example, therapies that maintain even minimal residual C-peptide secretion capacity have been found to have demonstrable clinical benefit^5^. An emerging barrier to this effort is disease heterogeneity. In particular, the rate of decline of β-cell capacity is highly variable, for reasons that remain unclear. As a result, clinical trial designs for disease-modifying therapies are necessarily cumbersome, requiring large sample sizes and prolonged observation. In addition, opportunities for tailored disease-modifying therapies are limited by an unclear understanding of the factors that drive diabetes progression after diagnosis. Gaining this knowledge could provide the attainment for improved participant inclusion in focused designed trials to foster their success and participant benefit^6^.

Studies with this goal conducted to date have typically been constrained by limitations to cohort size and the number of different data dimensions available for analysis^7, 8^. Thus, it has not been possible to conduct studies with an emphasis on hypothesis-generating, unbiased approaches, and integration of data across pathophysiological systems. These require large-scale, inter-disciplinary research efforts, in which carefully curated longitudinal clinical cohorts are aligned with multi-parametric technology platforms. Such a systems-based approach can address key questions with less bias and generate novel hypotheses on disease drivers. We used this strategy in the setting of a pan-European research consortium in which people with newly diagnosed type 1 diabetes as well as people at risk of developing type 1 diabetes (antibody positive) were enrolled into a master protocol^9^ to conduct a prospective study in search of factors that correlate with the rate of decline in β-cell mass/function. This endeavor was supported by the Innovative Medicines Initiative-2 Joint Undertaking, where INNODIA was created, being a private-public partnership of 40 partners in 16 countries. In the natural history study, people with newly diagnosed type 1 diabetes and unaffected family members are in follow up, allowing deep clinical characterization, as well as multi-omics analysis of samples (blood, urine, stool) collected and analyzed using standardized operating procedures (www.innodia.eu). Here we report the multi-omics analysis of the ‘first 100’ people with newly diagnosed disease. We report the existence of latent factors integrated from transcriptomic, small RNA, genomic, targeted proteomic, lipidomic, metabolomic, and immunomic-level data that show a relationship to subsequent rates of disease progression and have potential value as stratification and therapeutic target identification tools.

## Results

### C-peptide decline over time

The individual rates of decline in C-peptide levels over time were calculated, defined as the slope of C-peptide change over 12 months as described in Methods (Supplementary Fig. 1). Two individuals only had one available C-peptide measurement and were not assigned a group as C-peptide decline could not be determined. An overall trend of C-peptide decline over time was observed for the entire cohort (p-value 0.0001) (Fig. 2A). By calculating the C-peptide slopes using a linear mixed-effect model, the participants were divided into terciles (equal-sized), classified as rapid, slow, and increasing progression groups, yielding three groups with distinct progression patterns (Fig. 2B). All three progression groups had a similar estimated baseline C-peptide value (p-value 0.296), but significant C-peptide slopes (p-value < 0.0001). The clinical features for each of the progression groups are shown in Table 1.

**Fig. 1.**
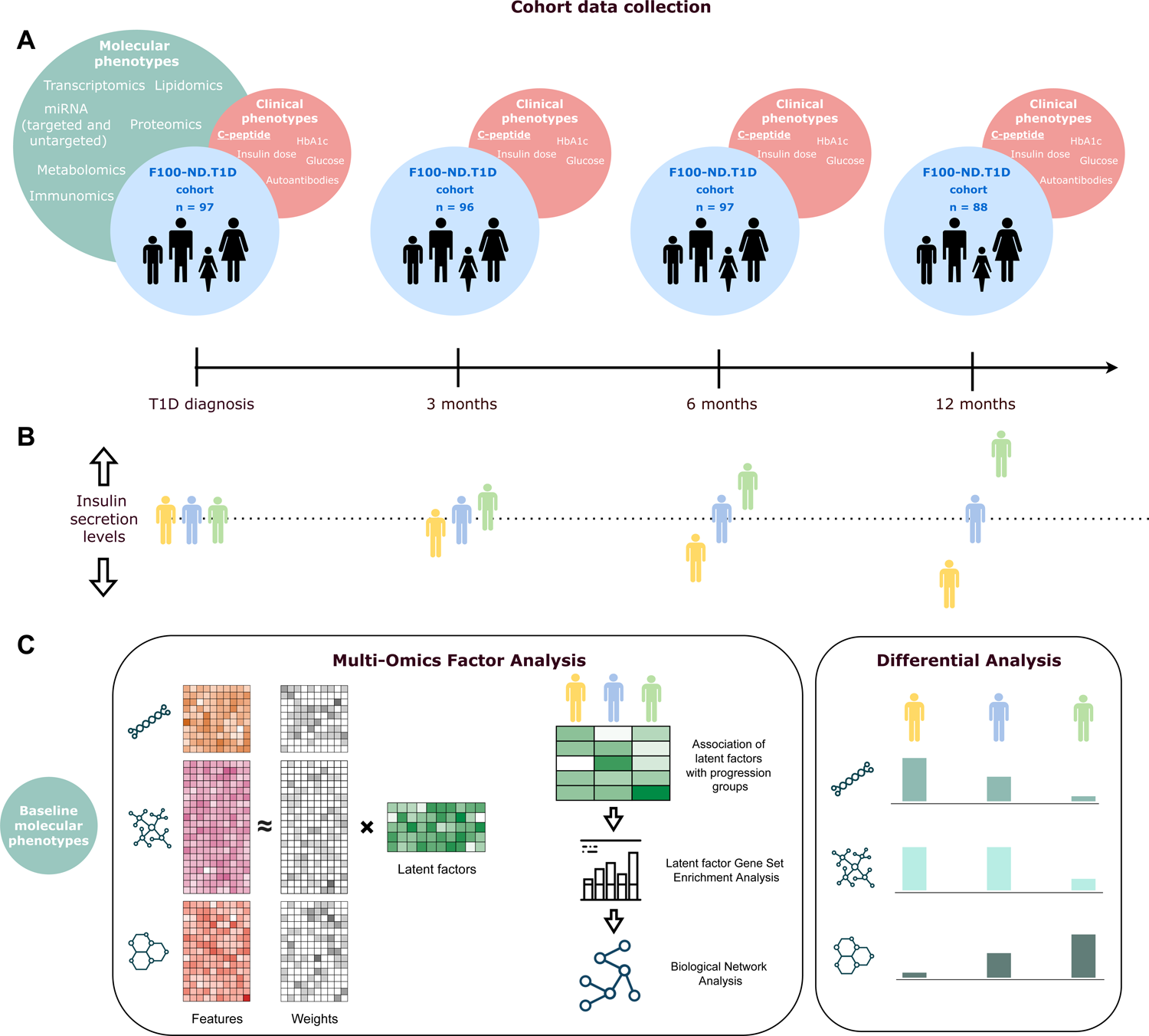
Cohort data and analysis overview. A) The cohort consists of 97 people with newly diagnosed type 1 diabetes. Multi-omics data were collected at baseline (within six weeks after diagnosis of type 1 diabetes) and clinical data were collected at baseline and at three, six, and 12 months. B) Participants were divided into three groups based on their change of insulin secretion levels (fasted C-peptide measurements) from baseline to 12 months. C) Multi-Omics Factor Analysis was performed to obtain an integrated signature across omics data types followed by differential expression analysis for each omics data type independently.

**Fig. 2:**
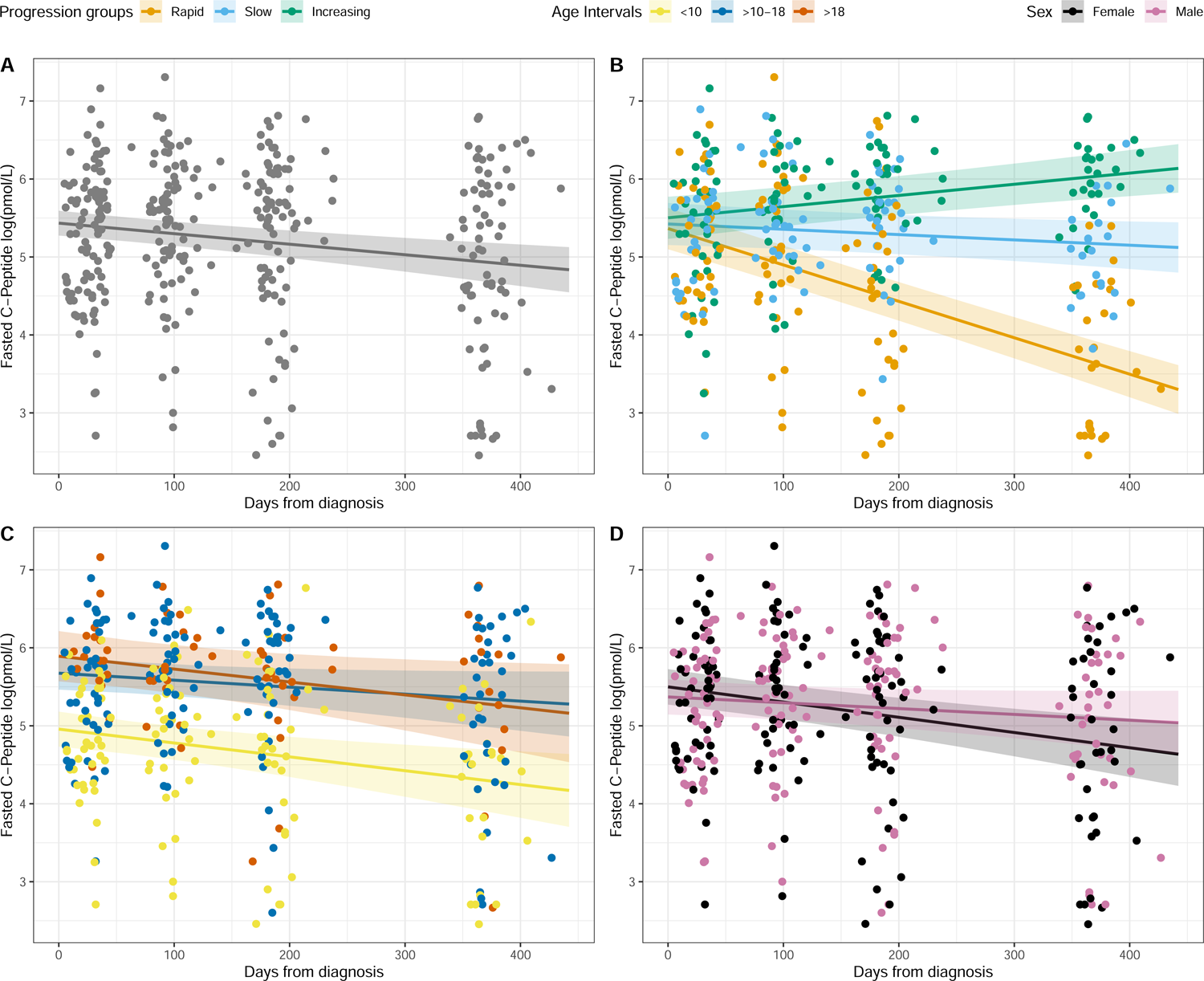
Fasted C-peptide trends over time. Time is represented as days from type 1 diabetes diagnosis. A) C-peptide values over time for all participants. B) C-peptide values over time divided into progression groups (progression terciles). C) C-peptide values over time for different age groups. D) C-peptide values over time for males and females, respectively.

**Table 1.**
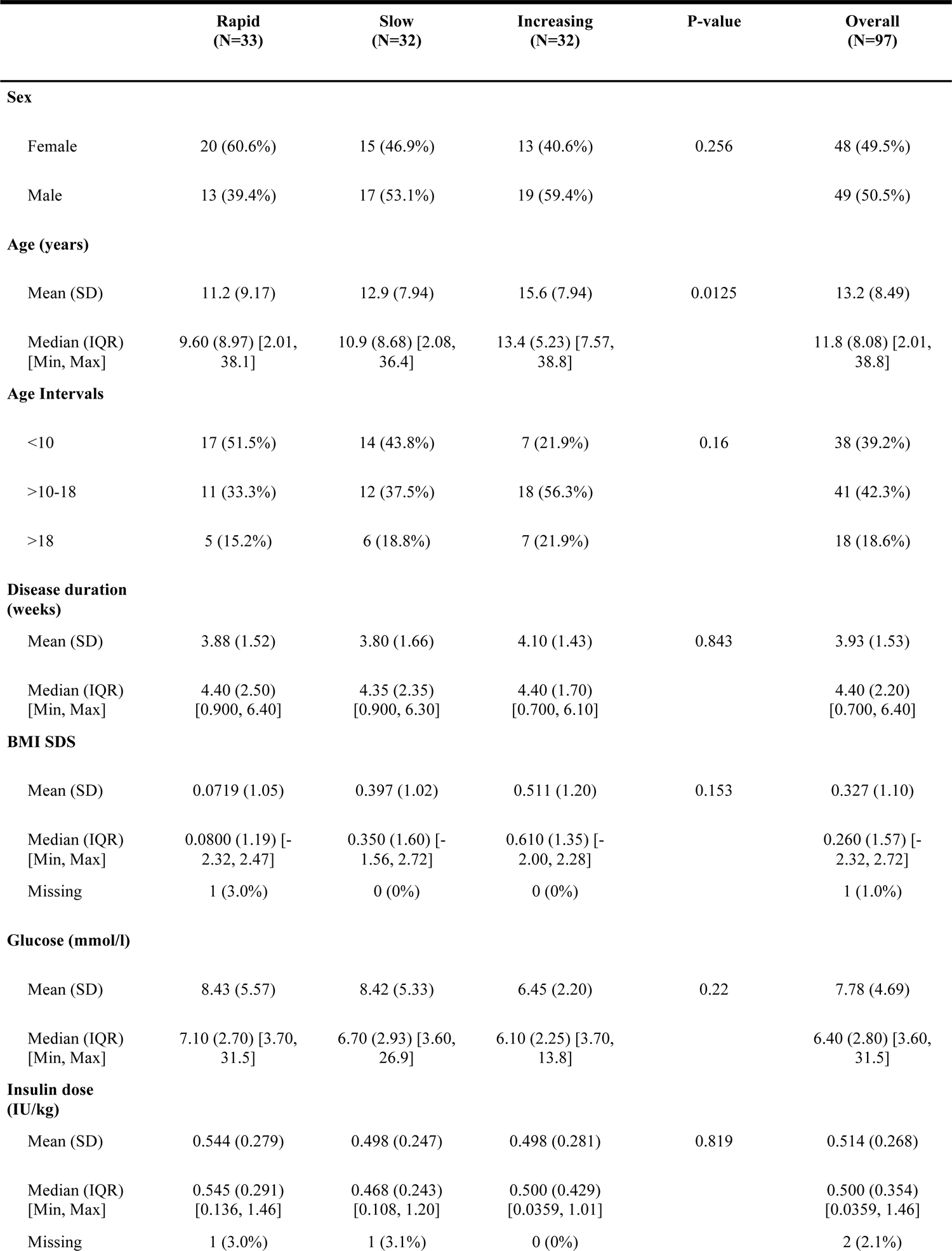
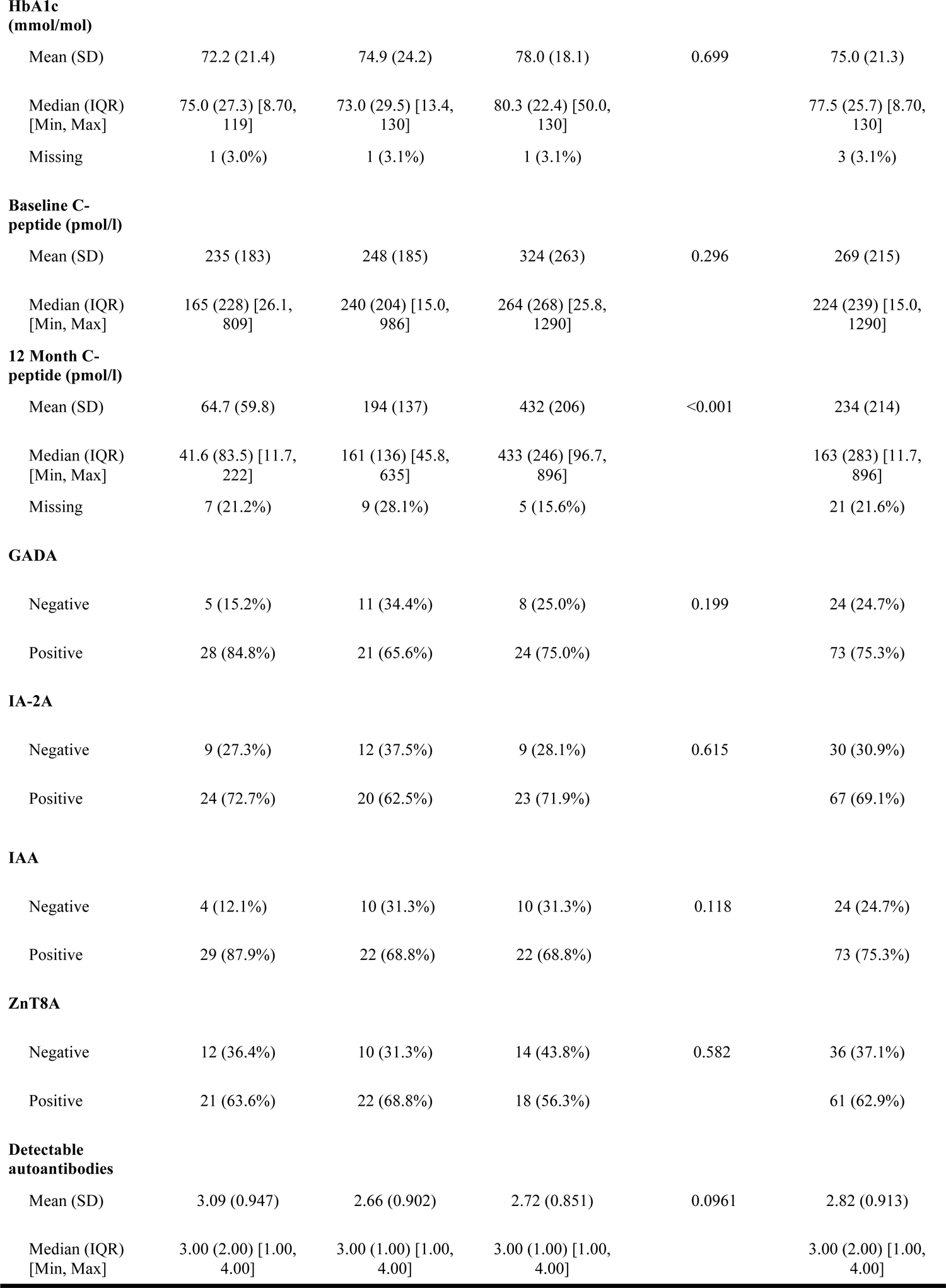
Clinical and demographic data for the type 1 diabetes cohort across progression groups. P-values were calculated using the Kruskal-Wallis test.

|  | Rapid<br>(N=33) | Slow<br>(N=32) | Increasing<br>(N=32) | P-value | Overall<br>(N=97) |
| --- | --- | --- | --- | --- | --- |
| **Sex** |  |  |  |  |  |
| Female | 20 (60.6%) | 15 (46.9%) | 13 (40.6%) | 0.256 | 48 (49.5%) |
| Male | 13 (39.4%) | 17 (53.1%) | 19 (59.4%) |  | 49 (50.5%) |
| **Age (years)** |  |  |  |  |  |
| Mean (SD) | 11.2 (9.17) | 12.9 (7.94) | 15.6 (7.94) | 0.0125 | 13.2 (8.49) |
| Median (IQR)<br>[Min, Max] | 9.60 (8.97) [2.01, 38.1] | 10.9 (8.68) [2.08, 36.4] | 13.4 (5.23) [7.57, 38.8] |  | 11.8 (8.08) [2.01, 38.8] |
| **Age Intervals** |  |  |  |  |  |
| <10 | 17 (51.5%) | 14 (43.8%) | 7 (21.9%) | 0.16 | 38 (39.2%) |
| >10-18 | 11 (33.3%) | 12 (37.5%) | 18 (56.3%) |  | 41 (42.3%) |
| >18 | 5 (15.2%) | 6 (18.8%) | 7 (21.9%) |  | 18 (18.6%) |
| **Disease duration (weeks)** |  |  |  |  |  |
| Mean (SD) | 3.88 (1.52) | 3.80 (1.66) | 4.10 (1.43) | 0.843 | 3.93 (1.53) |
| Median (IQR)<br>[Min, Max] | 4.40 (2.50) [0.900, 6.40] | 4.35 (2.35) [0.900, 6.30] | 4.40 (1.70) [0.700, 6.10] |  | 4.40 (2.20) [0.700, 6.40] |
| **BMI SDS** |  |  |  |  |  |
| Mean (SD) | 0.0719 (1.05) | 0.397 (1.02) | 0.511 (1.20) | 0.153 | 0.327 (1.10) |
| Median (IQR)<br>[Min, Max] | 0.0800 (1.19) [-2.32, 2.47] | 0.350 (1.60) [-1.56, 2.72] | 0.610 (1.35) [-2.00, 2.28] |  | 0.260 (1.57) [-2.32, 2.72] |
| Missing | 1 (3.0%) | 0 (0%) | 0 (0%) |  | 1 (1.0%) |
| **Glucose (mmol/l)** |  |  |  |  |  |
| Mean (SD) | 8.43 (5.57) | 8.42 (5.33) | 6.45 (2.20) | 0.22 | 7.78 (4.69) |
| Median (IQR)<br>[Min, Max] | 7.10 (2.70) [3.70, 31.5] | 6.70 (2.93) [3.60, 26.9] | 6.10 (2.25) [3.70, 13.8] |  | 6.40 (2.80) [3.60, 31.5] |
| **Insulin dose (IU/kg)** |  |  |  |  |  |
| Mean (SD) | 0.544 (0.279) | 0.498 (0.247) | 0.498 (0.281) | 0.819 | 0.514 (0.268) |
| Median (IQR)<br>[Min, Max] | 0.545 (0.291) [0.136, 1.46] | 0.468 (0.243) [0.108, 1.20] | 0.500 (0.429) [0.0359, 1.01] |  | 0.500 (0.354) [0.0359, 1.46] |
| Missing | 1 (3.0%) | 1 (3.1%) | 0 (0%) |  | 2 (2.1%) |
| **HbA1c (mmol/mol)** |  |  |  |  |  |
| Mean (SD) | 72.2 (21.4) | 74.9 (24.2) | 78.0 (18.1) | 0.699 | 75.0 (21.3) |
| Median (IQR) [Min, Max] | 75.0 (27.3) [8.70, 119] | 73.0 (29.5) [13.4, 130] | 80.3 (22.4) [50.0, 130] |  | 77.5 (25.7) [8.70, 130] |
| Missing | 1 (3.0%) | 1 (3.1%) | 1 (3.1%) |  | 3 (3.1%) |
| **Baseline C-peptide (pmol/l)** |  |  |  |  |  |
| Mean (SD) | 235 (183) | 248 (185) | 324 (263) | 0.296 | 269 (215) |
| Median (IQR) [Min, Max] | 165 (228) [26.1, 809] | 240 (204) [15.0, 986] | 264 (268) [25.8, 1290] |  | 224 (239) [15.0, 1290] |
| **12 Month C-peptide (pmol/l)** |  |  |  |  |  |
| Mean (SD) | 64.7 (59.8) | 194 (137) | 432 (206) | <0.001 | 234 (214) |
| Median (IQR) [Min, Max] | 41.6 (83.5) [11.7, 222] | 161 (136) [45.8, 635] | 433 (246) [96.7, 896] |  | 163 (283) [11.7, 896] |
| Missing | 7 (21.2%) | 9 (28.1%) | 5 (15.6%) |  | 21 (21.6%) |
| **GADA** |  |  |  |  |  |
| Negative | 5 (15.2%) | 11 (34.4%) | 8 (25.0%) | 0.199 | 24 (24.7%) |
| Positive | 28 (84.8%) | 21 (65.6%) | 24 (75.0%) |  | 73 (75.3%) |
| **IA-2A** |  |  |  |  |  |
| Negative | 9 (27.3%) | 12 (37.5%) | 9 (28.1%) | 0.615 | 30 (30.9%) |
| Positive | 24 (72.7%) | 20 (62.5%) | 23 (71.9%) |  | 67 (69.1%) |
| **IAA** |  |  |  |  |  |
| Negative | 4 (12.1%) | 10 (31.3%) | 10 (31.3%) | 0.118 | 24 (24.7%) |
| Positive | 29 (87.9%) | 22 (68.8%) | 22 (68.8%) |  | 73 (75.3%) |
| **ZnT8A** |  |  |  |  |  |
| Negative | 12 (36.4%) | 10 (31.3%) | 14 (43.8%) | 0.582 | 36 (37.1%) |
| Positive | 21 (63.6%) | 22 (68.8%) | 18 (56.3%) |  | 61 (62.9%) |
| **Detectable autoantibodies** |  |  |  |  |  |
| Mean (SD) | 3.09 (0.947) | 2.66 (0.902) | 2.72 (0.851) | 0.0961 | 2.82 (0.913) |
| Median (IQR) [Min, Max] | 3.00 (2.00) [1.00, 4.00] | 3.00 (1.00) [1.00, 4.00] | 3.00 (1.00) [1.00, 4.00] |  | 3.00 (2.00) [1.00, 4.00] |

We further analyzed the relationship between age and C-peptide change over time (Fig. 2C) as this was the only significant difference between the progression groups (Table 1). Participants less than ten years old had a significantly lower baseline C-peptide level compared to those older than ten years (p-value < 0.0001), whereas baseline values did not differ between the 10-18 years and >18 years age groups (p-value 0.85). However, the C-peptide change over time was not significantly different between the three age intervals (p-value 0.46). This indicated that age is associated with baseline C-peptide values but the decline in C-peptide over time is similar for all age groups. In addition, when investigating the distributions of C-peptide decline rates, children < 10 years old had a significantly different distribution of decline rates than those aged 10-18 years (p-value 0.0072). However, neither 10-18 year and >18 year groups nor the >18 year and < 10 year groups had different distributions (p-values 0.075 and 0.7, respectively) (Supplementary Fig. 2). These findings indicate a degree of association between the rate of decline and age among children, and as a result, age was included after log transformation as a covariate in our models to correct for potential effects on the relationship between C-peptide slopes and ‘omics. Evaluating the association of sex with the C-peptide change over time (Fig. 2D) we found no significant association with baseline C-peptide (p-value 0.64) or slope (p-value 0.16).

### Multi-omics integration analysis

The multi-omics data set overview is shown in Fig. 3A. Missing values, seen predominantly in the immunomics data set, are disregarded by MOFA and do not affect the decomposition of the data into latent factors. After training MOFA on the multi-omics data set (Fig. 3B) the latent factors that capture most of the variance across participants were represented by the mRNA and miRNA data. MOFA captures latent factors with common variance across the different data sets, even though certain data types appear to be responsible of most of the captured variance (Factors 1 to 7). This indicates that certain degree of heterogeneity exists across data sets, making the integration more challenging.

**Fig. 3.**
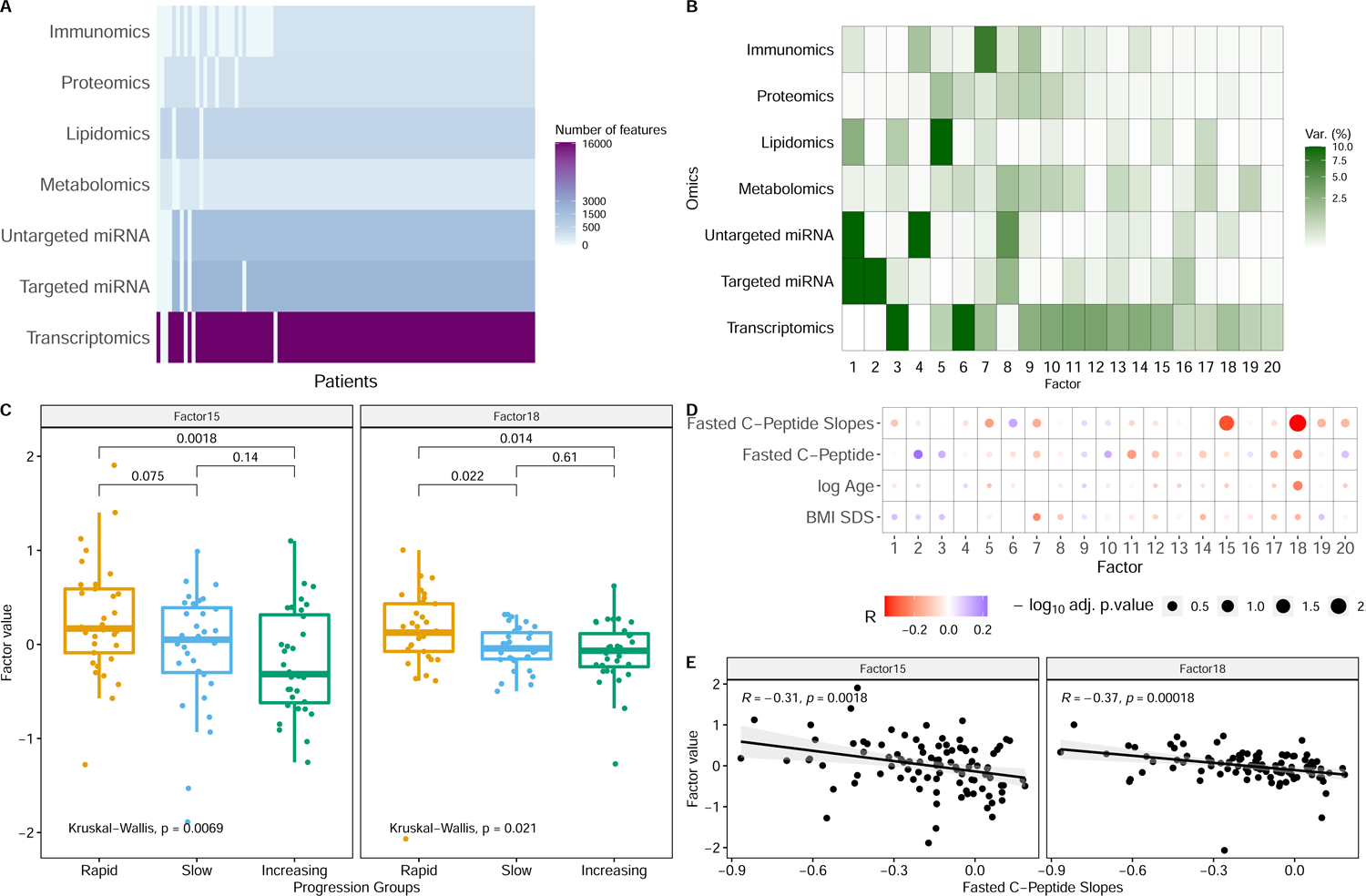
A) Overview of the multi-omics data sets, describing the number of features per data set and the level of missing data (white). B) Latent factors obtained from MOFA, the color scale represents the variance captured by each of the latent factors indicating the level of integration of the data types for each factor. C) Association of latent factors 15 and 18 values with the different progression groups. D) Spearman correlation of latent factors with the fasted C-peptide slopes, baseline fasted C-Peptide, Age (log scale), and BMI-SDS. P-values were adjusted by Benjamini-Hochberg. E) Spearman correlation of fasted C-peptide slopes against latent factor values (15 and 18).

Importantly, however, latent factors 15 and 18 were significantly associated (p-values < 0.1 adjusted by Benjamini-Hochberg) with C-peptide slopes (Fig. 3D) but not with age or baseline C-peptide. The amount of variance captured by latent factors 15 and 18 is 2.62% and 1.84%, respectively, indicating that the decline of C-peptide over time is not among the major sources of variance across the participants, but is sufficiently strong to be captured by this method. The differences in the strength of associations for latent factors 15 and 18 with the progression groups and C-peptide slopes indicated that latent factor 15 captures a non-linear association with the progression groups (Fig. 3C), and for latent factor 18 a linear association with the rate of C-peptide decline (Fig. 3E). As the two factors correlate with C-peptide decline, they may contain molecular signatures useful for explaining the differences in disease progression between patients. Therefore, we continued a thorough scrutiny of these factors.

### Differential gene expression analysis

Differentially expressed genes (DEGs) were identified between the different progression groups, with batch and age groups used as covariates (Fig. 4). P-values were adjusted for multiple testing using the Benjamini-Hochberg procedure and genes with an adjusted p-value < 0.1 were reported as differentially expressed genes (DEG). Fig. 4 shows the volcano plots of the different comparisons together with the genes belonging to latent factors 15 and 18. A total of 339 DEGs were observed comparing the rapid decline group and the increasing group (Fig. 4A, Supplementary Table 7), 33 DEGs between the rapid and slow decline group (Fig. 4B, Supplementary Table 8), and 1,205 DEGs between the slow decline group and the increasing group (Fig. 4C, Supplementary Table 9). Additionally, differential gene expression was performed for the C-peptide slopes, hence studying the linear change in gene expression with respect to the rate of decline of C-peptide over time. Here we found 484 DEGs (Fig. 4D, Supplementary Table 10).

**Fig. 4.**
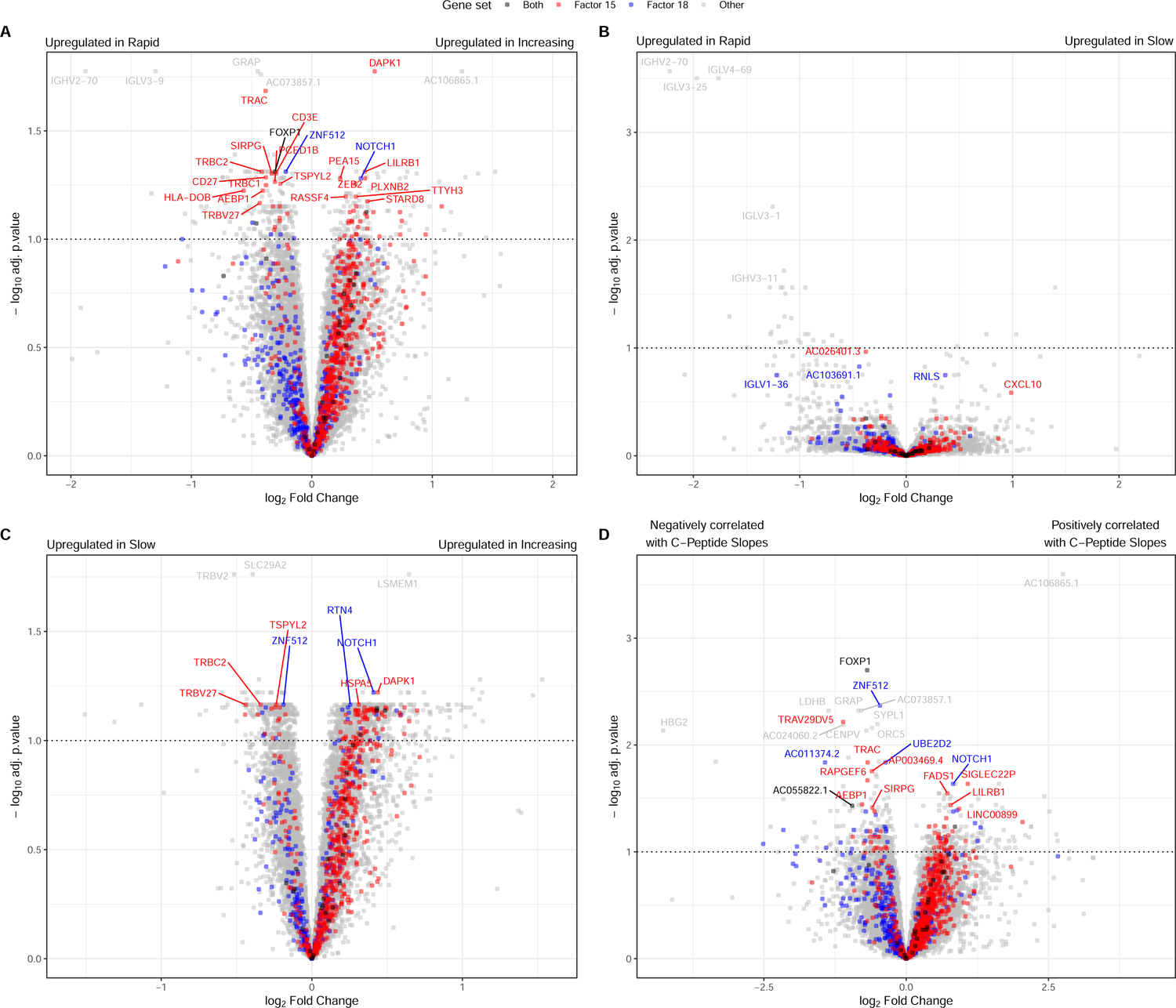
Volcano plot of differential gene expression between progression groups. The color indicates gene membership to either or both associated latent factors. A) DGE between rapid and increasing progression groups (339 genes are differentially expressed). B) DGE between rapid and slow progression groups (33 genes are differentially expressed). C) DGE between slow and increasing progression groups (1,206 genes are differentially expressed). D) DGE for C-peptide slopes (484 genes are differentially expressed).

More DEGs were observed when comparing the slow and increasing than between rapid and increasing progression groups, with little overlap between the significant top-ranking genes. Additionally, very few genes were differentially expressed between rapid and slow progression groups. Altogether, these results might indicate that even though the rapid and slow progression groups are more alike, the set of DEGs between these two groups and the increasing group are not the same. To further confirm this, two additional differential expression analyses were performed. One between the rapid-slow groups combined versus the increasing group and another between the slow-increasing groups combined versus the rapid. The analyses showed that the rapid-slow vs increasing comparison produced 1,804 DEGs (Supplementary Table 11), while the slow-increasing vs rapid comparison produced 18 DEGs (Supplementary Table 12). This indicates that the increasing progression group is much more dissimilar in its blood sample expression profile towards the other two groups at the early stage of type 1 diabetes manifestation. Therefore, the underlying biological processes involved in the developing disease progression do not vary much at this early time of the disease manifestation among people experiencing in the future different degrees of loss of β-cell function. At baseline they vary significantly more when comparing people experiencing in the future loss of β-cell function and those experiencing an improvement in β-cell function.

The similarity between DEGs found by the continuous change in C-peptide levels (n=484) and DEGs between the rapid and increasing group (n=339) showed an overlap of 209 genes. The rapid-slow vs increasing DEGs (n=1,804) had a bigger overlap with the continuous C-peptide levels where 313 of the same DEGs were found. In all cases, the DEGs had the same sign of their log2 fold changes for both analyses. Therefore, most DEGs were observed for the continuous change were also found when investigating progression groups. Nonetheless, we believe that the linear association of blood gene expression at baseline with the C-peptide slopes is more informative regarding the disease progression. We observe that change in β-cell function follows a gradient, so by categorizing participants into groups, we lose the resolution that the C-peptide slopes are providing.

### Annotation of latent factors

To examine the biological pathways in the two most relevant latent factors, we used gene set enrichment analysis. This analysis was divided into genes positively regulated in the rapid decline group and genes negatively regulated in the rapid decline group. Fig. 5 displays the top 15 significant pathways in each of the two associated latent factors (p-values < 0.1 adjusted by Benjamini-Hochberg). Negatively regulated genes in latent factor 15 are enriched in the immune system and signaling by G protein-coupled receptors (GPCR) pathways. Of specific interest are pathways associated with innate immunity, such as neutrophil degranulation, with high expression of granule proteins (e.g. CTSs, MPO) pointing to the presence of activated or degranulated neutrophils; and platelet activation, signaling and aggregation (the latter not shown). Also, several pathways pointing to cytokine signaling and interleukin (e.g. IL-1β) signaling emerge. Regarding the role of GPCRs, several pathways are associated with latent factor 15, such as signaling by GPCRs, downstream signaling and GPCR ligand binding. Positively regulated genes in latent factor 15 are also enriched in immune system pathways, with again an important contribution of the innate immune system, although here it seems that the increase is mainly attributed to resting neutrophils, with for instance high expression of LY96. Collectively these data show that GSEA pathways in innate immunity are mainly associated with the activation status of the neutrophils, with shift in the balance of resting neutrophils versus activated or degranulated neutrophils.

**Fig. 5.**
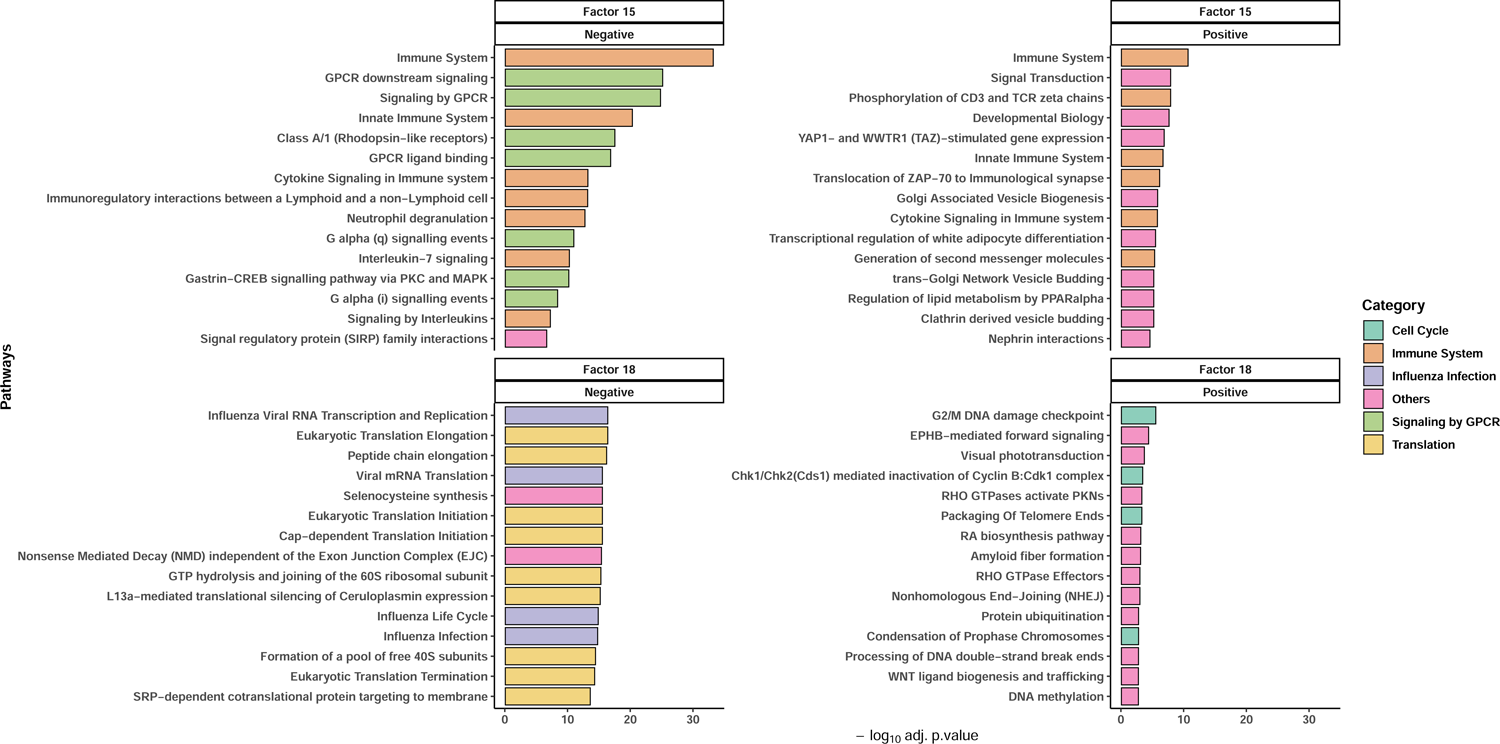
GSEA was performed separately for genes with positive weights in the latent factor (upregulated in rapid decline) and genes with negative weights in the latent factor (downregulated in rapid decline). Pathways are coloured depending on the main pathway they belong to, according to Reactome. P-values were adjusted using Benjamini-Hochberg.

Negatively regulated genes in latent factor 18 are enriched in influenza infection pathways and mRNA translation pathways, suggesting that viral mRNA replication and translation by host cell machinery are major pathological features of this association. Positively regulated genes in latent factor 18 did not show a particular pattern of enrichment.

Furthermore, the relation of the latent factors’ genes to previous type 1 diabetes publications was studied. Using the Open Targets database^10^, a total of 174 genes out of the 668 top genes (three-fold higher than expected by chance) in the latent factor 15 have been previously associated with type 1 diabetes (p-value 0.03). On the other hand, the overlap between the top genes in the latent factor 18 and associated disease genes was not significant. These results indicate that latent factor 15 is capturing a set of genes composed of known disease targets (e.g. INSR, NDUFA4, CTSH) as well as potential new candidate genes already detectable in blood in the early phase of type 1 diabetes.

### Interpretation of biological networks

Biological networks were constructed for latent factors 15 and 18 separately, based on protein-protein or protein-protein-miRNA interactions (Supplementary Fig. 3 and 4). As the two network types yielded similar results, we selected the protein-protein-miRNA networks for the focus of our interpretations. The latent factor 15 network revealed a diverse set of biological functions (Supplementary Fig. 5), some of them overlapping with the pathways shown in Fig. 5. Immune system responses, signaling by different receptors and lipid metabolism are (widely) represented in these clusters. Notably there is an enrichment of lipid metabolism pathways as the lipidomic data also influenced latent factor 15. Some of the genes significantly associated with the C-peptide slopes also appear in several of the clusters, which further validates the biological processes captured by the latent factor. The latent factor 18 network was smaller and had more loosely defined clusters (Supplementary Fig. 6). Nonetheless, it captured a similar set of biological functions compared to the latent factor 15 network. Eukaryotic/viral mRNA translation is the main difference between the two networks, which appear as the biggest and more interconnected cluster of the latent factor 18 network. In this case, only one of the genes in this network was significantly associated with the C-peptide slopes.

### Immunomics signature

Analysis of immune cell populations identified based on standard markers and their association with the C-peptide slopes revealed that Natural Killer (NK) cells were significantly associated after p-value adjustment, (Fig. 6A), with higher levels of NK cells observed in people with slow disease progression (Fig. 6B). Examination of the relationship between C-peptide slope and NK cell frequency in individual progression groups indicated the strongest correlation was observed among rapid progressors (Fig. 6C). Unsupervised analysis of the immunomics data revealed distinct clusters among the progression groups. Fig. 7A shows a FlowSOM color-density map of CD16 expression levels with node sizes representing the frequency of cells in each cluster. Meta-cluster 19 (MC-19) was assigned as the primary NK (CD56loCD16+) subset based on lineage marker expression and was significantly more abundant in the increasing versus rapid progression groups (8.8 vs 6.3% p-value=0.013). Examination of markers of NK cell activation and differentiation (KLRG1, TIGIT, CD38, and CD57) in the different progression groups showed higher expression of KLRG1, in the increasing group but lower levels of CD38.

**Fig. 6.**
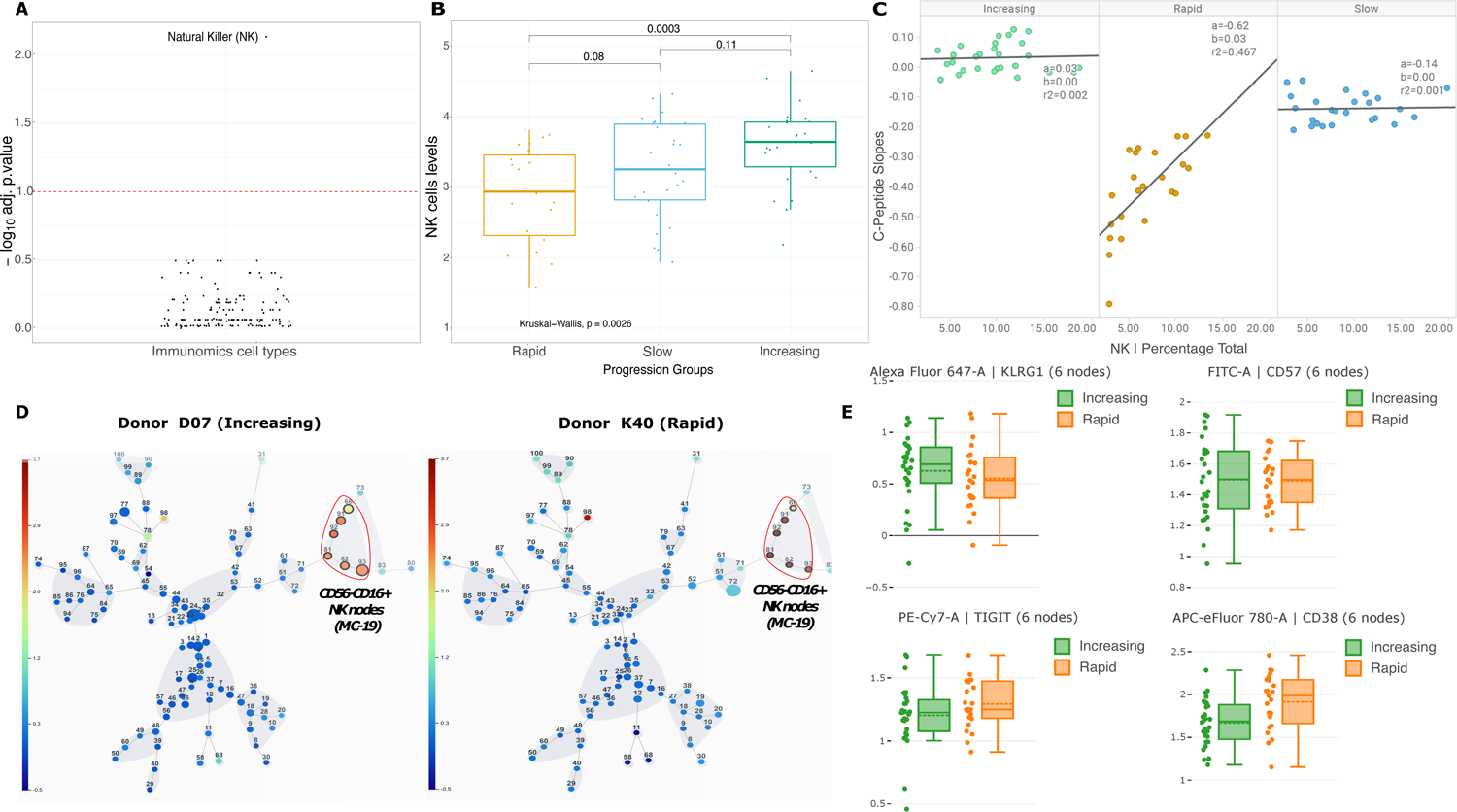
Immunomics association with C-peptide slopes. A) Benjamini-Hochberg adjusted p-value for the linear association of immune cells abundances with the C-peptide slopes (correcting for batch effects and age groups). B) NK cell levels for the different progression groups (as a frequency of total live mononuclear cells). C) Spearman correlation of fasted C-peptide slopes versus NK cell frequency for the different progression groups. D) FlowSOM unbiased cluster analysis on live CD45^+^ PBMCs of Donor D07 (Increasing, left) and K40 (Rapid, right) highlighting Metacluster-19 (red-circle) as a primary NK (CD56^lo^CD16^+^) subset. Colour-density and circle size were overlaid as CD16 intensity and number of events, respectively. E) Bar charts representing NK marker expression on the selected clusters in each progression group.

Comparison of the genes associated with latent factor 15 and the leukocyte gene signature matrix LM22^11^ revealed an overlap of 52 genes and enrichment in immune cell-specific genes (p-value 0.0001). The genes were representative of the following five main groups, T-cell specific, macrophage M1 specific, monocyte specific, neutrophil specific, and eosinophil specific (Supplementary Fig. 7). In this way, in addition to capturing immune-related pathways, the latent factor 15 broadly represented immune cell-specific genes.

### Other ‘omics associations with disease progression

The association of factors derived from running MOFA without the transcriptomics data, did not yield any significant associations with the progression groups nor the C-peptide slopes. We can draw two conclusions based on these results. Firstly, the variance of the miRNA and lipidomics captured by latent factor 15 might indicate that these two omics data sets are only informative of the disease progression in combination with transcriptomics. Interestingly, an enrichment on lipid metabolism pathways was apparent among the genes associated with latent factor 15, indicating that these pathways might have an effect of disease progression. This shows how integration of both lipidomics and transcriptomics data aid in discovering enrichment in specific pathways. Similarly, the incorporation of transcriptomics and miRNA data allowed the discovery of enrichment in viral infection pathways.

Secondly, the main difference between transcriptomics and the other omics data was that it was determined from whole blood. The remaining omics are collected either from serum, plasma, or PBMCs. Even though some analytes may give similar results, samples obtained from the same medium are more easily comparable. Therefore, the disparity between omics that we observe in the latent factors and in the linear association of each omics data set might be caused by the source medium. This could be considered both a drawback and an advantage. On one hand, it is undesirable that this disparity exists because correlated analytes across omics cannot be studied. This makes the data integration challenging as we cannot observe the joint effect of multiple omics nor validate whether the analytes associated with the disease progression in one data type can also be observed in a different one. On the other hand, the heterogeneity across omics can be seen as complementary information. Each omics data set is capturing a different source of variation, thus, providing additional information not captured by the other omics types. Based on the current data, we cannot conclude whether the plasma and serum omics were not associated with the disease progression due to the medium or the analytes themselves.

## Discussion

In this study, we identified two latent factors associated with β-cell decline. These factors were predominantly influenced by transcriptomics, with secondary contributions of miRNA and lipidomics, respectively. Latent factor 15 revealed an enrichment of immune system pathways, the most significant being associated with neutrophil degranulation, cytokine signaling, and immunoregulatory interactions between lymphoid and non-lymphoid cells. Moreover, there were multiple pathways associated with GPCR signaling events. More detailed GSEA revealed that disease progression (C-peptide slopes) was associated with an altered balance between resting and activated/degranulated neutrophils. This is in keeping with previous studies that showed a temporary decline in the number of circulating neutrophils in people with newly diagnosed type 1 diabetes, compared to healthy controls, as well as high circulating levels of neutrophil extracellular traps (NETs)^12–14^. The previous detection of neutrophils and NETs in the pancreas of deceased subjects affected by type 1 diabetes, and a correlation between circulating neutrophil numbers with C-peptide levels in pre-symptomatic subjects (non-diabetic, at-risk) has implicated that activated neutrophils play a pathogenic role in type 1 diabetes^15^. Our data add significantly to this hypothesis since we show in longitudinal follow-up that the neutrophil profile at diagnosis associates with rate of disease progression.

Of further interest, GSEA shows platelet activation to be a feature of latent factor 15, linking our findings to the recent demonstration of a role for activated platelets in the formation of platelet-neutrophil aggregates (PNAs), which are increased in the circulation of subjects during the development of type 1 diabetes^16^. It is tempting to speculate that activation of GPCR pathways (also associated with latent factor 15) may play a role in these events since it is a response to a variety of stimuli (chemokines, cytokines, complement fragments) and can trigger neutrophil degranulation^17^.

In contrast, latent factor 18, which was also associated with β-cell decline, is characterized by features of enrichment of viral mRNA translation and subsequent translation by the host cell machinery. There is a considerable body of literature that associates viral infections with early events in type 1 diabetes development as well as peri-diagnosis events^18^. As a result, virus infection has often been cited as an autoimmunity-triggering event as well as a disease-precipitating event. Our findings in the context of the present study design are entirely consistent with the latter hypothesis, which could be addressed in follow-up viromic studies targeted to samples in which both the relevant viral mRNA translation signals and negative slope of C-peptide decline are prominent.

Molecular and cellular signatures of adaptive immune responses were by and large not observed to be associated with β-cell decline in our study, which might at first sight be considered a surprise, given the strong credentials, at genomic, pathological and mechanistic levels, for type 1 diabetes being the archetype of an organ-specific autoimmune disease. However, it is entirely plausible that the detection of such associations is challenging in whole blood or whole mononuclear cell analyses, both because the disease-relevant, β-cell antigen-specific lymphocytes are rare, and even more importantly, because they may be sequestered at inflammatory sites. Certainly, smaller scale studies focused on using appropriately sensitive technologies have identified that the activation and differentiation state of circulating antigen-specific cytotoxic T lymphocytes, for example, correlate with changes in β-cell function after the diagnosis of type 1 diabetes^8^. Amongst the lymphocyte studies presented here, our observation of a prominent NK cell signal related to rapid β-cell decline is of considerable interest. NK cells have features of both innate and adaptive immune cells and play a key role in anti-viral responses. Both pro-inflammatory and regulatory functions have been ascribed to these cells, and functional subtypes can be partially differentiated by surface markers. Previous studies examining the frequency of NK cells in individuals affected by type 1 diabetes have consistently reported lower circulating levels of both proinflammatory and regulatory NK cells when compared to aged-matched non-diabetic subjects^10, 19, 20^, potentially reflecting homing to inflammatory sites in the pancreas. Consistent with this, we observed lower circulating levels of NK cells (both effector and regulatory subtypes according to surface markers) in the rapid decline group. Of interest, reduced circulating NK cell levels are also associated with viral infection, linking this observation to the viral signatures already described. Future functional studies will be required to explore the pathological implications of these findings, since the immune phenotyping performed here was limited to expression of CD38 (NK cell activation) and KLRG1 (an inhibitory receptor associated with an exhausted phenotype).

Key strengths of the study include (i) the setting of a large, longitudinal natural history study conducted across multiple European sites according to standardized clinical and laboratory protocols; (ii) access through the INNODIA network to highly specialized, systems-based technology platforms for parallel multi-parametric analysis; (iii) leverage of new tools in integrated multi-omics factor analysis to discover signatures that are significantly associated with rate of disease progression for the year following diagnosis. This period of the disease is important since it represents the phase during which disease-modifying immunotherapies are typically trialed for their effect on arresting β-cell decline. Factors identifiable at baseline that associate with faster β-cell decline could be used to conduct shorter and smaller trials (e.g. by enrollment of a rapid-decline group), an important step towards de-risking the investment needed to bring disease modifying strategies into clinical use.

Finally, we want to highlight specificities of the present study that can be interpreted as limitations or strengths. Our INNODIA natural history collection involves individuals from the ages of one up to 45 years, and thus includes the whole lifespan of people living with type 1 diabetes. Although this may render interpretation of findings in this clearly heterogenic disease more complex, it may add to the identification of common factors that drive disease, irrespective of age. We used fasting C-peptide as read-out for β-cell function rather than stimulated C-peptide and could demonstrate similar trends in decline of function using this simple parameter. Although collecting throughout Europe, the population studied is almost completely white Caucasian, thus limiting the generalizability of the findings to a global population, where type 1 diabetes is becoming more prevalent in non-Caucasian people. Confirmation of our observations will therefore be needed in more diverse cohorts. Our work reports on a small cohort of just under 100 people with newly diagnosed disease. However, we demonstrate that even in such small numbers, using deeply phenotyped individuals and standardized operating procedures, application of systems biology techniques can lead to significant associations. Here we believe that the collaboration between academics, foundations, industry and people affected by type 1 diabetes and their families within INNODIA was a unique driver. We created strict protocols for follow up where people could be convinced to participate with support of materials created by the PAC (People with diabetes Advisory Committee), we set up a highly standardized sample collection (e.g. even standardizing the pipet tips for miRNA sample collection) and applied homogeneous laboratory procedures. Importantly, we brought all data into a GDPR-conform centralized database, allowing clean data collection and high quality data for input into the analysis.

In summary, the presented study addressed the drivers of disease heterogeneity in type 1 diabetes by leveraging opportunities presented by a highly integrated clinical network featuring embedded research platforms with the capability to generate large systems-level datasets. One of the two factors identified showed correlation to neutrophil degranulation, cytokine signaling, lymphoid and non-lymphoid cell interactions and G-protein coupled receptor signaling events, while the second signature, pathways related to translation and viral infection were inversely associated with change in β-cell function. The derived latent factors were used to identify specific signatures, which were further investigated for biomarker opportunities and mechanistic pathways that correlate with β-cell decline. This shows how multi-omics analysis can be used as an important foundation for the development and testing of disease-modifying therapies in the future.

## Methods

### Subjects with type 1 diabetes

For this in depth analysis, we included the first 100 subjects with newly diagnosed (<6 weeks) type 1 diabetes enrolled in the INNODIA natural history study. Using a consecutive recruitment approach, subjects were included based on baseline omics data availability, an even gender distribution and biosample availability, positivity for at least one diabetes-related autoantibody (GADA, IA-2A, IAA, ZnT8A), and age between one to 45 years. Two subjects were excluded due to incomplete ‘omics datasets and one following the detection of a MODY gene mutation. The final analysis cohort comprised 49 male and 48 female study participants (Table 1), the average age at diabetes diagnosis of 13.2 years (SD 8.5; two-ten years n=38; ten-18 years n=41, 18-39 years n=18), mean disease duration of 3.9 weeks (SD 1.5) and at baseline an average total daily insulin dose of 0.51 IU/kg (SD 0.27), HbA1c of 75 mmol/mol (SD 21.3), fasting C-peptide level of 269 pmol/l (IQR 25.7), fasting glucose level of 7.78 mmol/l (IQR 2.8) and BMI SDS of 0.327 units (SD 1.1). Fasted C-peptide measurements were made at four visits (Fig. 1A). To define the rate of C-peptide decline over time, we utilized linear mixed-effect models to fit the log-transformed fasted C-peptide from day of diagnosis to 12 months (Fig. 1B). The model was fitted using subject-level random effects and the rate of C-peptide change over time. Mixed-effect models were fit using the lme4 R package^21^ with an unstructured random effects variance-covariance matrix.

Some individuals did not complete all visits. A total of 69 individuals completed the four visits (baseline, three, six, and 12 months), 21 individuals completed three visits, five individuals completed two visits, and two individuals completed only the baseline visit (not necessarily consecutive visits). At each visit HbA1c was measured and stimulated C-peptide response was determined by mixed meal tolerance test (MMTT) from individuals of at least five years of age. The islet autoantibodies GADA, IA-2A, IAA, and ZnT8A were quantified with the use of specific radiobinding assays as described earlier^22^.

### Targeted serum proteomics by liquid chromatography-mass spectrometry (LC-MS/MS)

Targeted proteomics of baseline serum samples from 91 individuals was performed using liquid chromatography with selected reaction monitoring (SRM) mass spectrometry (LC-MS/MS) to measure 195 peptides, representing 98 target proteins and retention time markers. The list of peptides and proteins measured is provided in Supplementary Table 1.

The samples were reduced, digested, and alkylated, then spiked with isotopically labelled synthetic analogues of the targets, as previously described^23^. The batchwise analyses of the samples were made with the periodic inclusion of three internal QC reference controls. Skyline software^24^ was used to both develop the acquisition method and perform primary processing of the data, including peak integration and quality assessment. The un-normalized data was subsequently exported and prepared for batch correction and normalization^25^.

### Whole blood transcriptomics

We performed transcriptome analysis on whole blood samples collected from 92 individuals as well as four reference RNAs (two anonymous donors) used for normalization and assessing batch effects across sample pools. Prior to RNA extraction, frozen whole blood PAXgene samples were thawed at room temperature for 2 hours and subjected to RNA extraction using PAXgene Blood miRNA Kit (PreAnalytix/QIAGEN, Cat# 763134). Total RNA, including RNA longer than approximately 18 nucleotides, was purified according to the manufacturer’s protocol. Sample concentration was measured with a Nanodrop 2000 spectrophotometer and Qubit Fluorometric Quantitation (Thermo Fisher Scientific).

The quality of the samples was ensured with the Experion Automated Electrophoresis System (Bio Rad) and Agilent 2100 Bioanalyzer RNA Pico chip. Library preparation and sequencing were carried out at the Finnish Functional Genomics Centre (FFGC). Before starting library preparation, ERCC Spike-in Mix 1 (Invitrogen P/N 4456739) was added to 100 ng RNA according to the kit’s protocol. RNAseq libraries were prepared using the TruSeq stranded mRNA HT kit and protocol # 15031047 (Illumina). The quality and quantity of the amplified libraries were measured using Advanced Analytical Fragment Analyzer (Agilent) and Qubit Fluorometric Quantitation, respectively. Pooled libraries were sequenced on an Illumina NovaSeq 6000 instrument, using 2×50 bp paired-end sequencing. bcl2fastq2 Conversion Software v2.20.0.422 was used to convert base call files into FASTQ files. The quality of the raw sequencing reads was checked using the FastQC tool (v. 0.11.14). The sequencing data preprocessing was carried out using R (v. 3.6.1) and the related Bioconductor module (v. 3.9). The reads were aligned to UCSC hg38 human reference genome (downloaded from Illumina iGenomes site https://support.illumina.com/sequencing/sequencing_software/igenome.html) using Rsubread package (v. 1.34.7) and the same package was used to produce the read counts for the RefSeq annotated genes. The gene-wise counts per million (CPM) were generated using the edgeR package (v. 3.26.8).

### Circulating Small RNAs sequencing

Expression levels of circulating Small RNAs were analyzed from baseline plasma samples of n=91 subjects on the same sample set adopting two different Next Generation Sequencing (NGS) approaches: *(i)* probes-based sequencing focused on miRNAs (targeted) through HTG EdgeSeq miRNA whole transcriptome assay (Supplementary Table 2); *(ii)* Small RNAs sequencing using the QiaSeq miRNA/small RNA library preparation kit (untargeted).

Plasma samples were shipped to HTG Molecular Inc. in order to be analyzed using rigorous standard procedures. The HTG EdgeSeq miRNA whole transcriptome assay method is an RNA extraction free approach that exploits quantitative nuclease protection assay chemistry using sequence-specific nuclease-protection probes (NPPs). This was followed by an NGS step, in order to allow semi-quantitative analysis of a panel of n=2102 targeted miRNAs (including n=13 housekeepings, n=5 negative process controls, n=1 positive process control and n=2083 targeted miRNAs) from 15 µL of plasma. HTG EdgeSeq Plasma Lysis buffer was added to 15 µL of each plasma sample. Lysed samples were then transferred to a standard 96-well plate. The NPPs were added to the lysed samples followed by the addition of S1 nuclease to digest non-hybridized RNA. The nuclease digestion reaction was then stopped and each processed sample was used as a template for PCR-based library preparation using specifically designed primers (tags), which share common sequences complementary to 5’-end and 3’-end “wing” sequences of the probes and common adapters required for cluster generation on Illumina NGS platform. Libraries were prepared in accordance with HTG standard procedures, HTG EdgeSeq PCR processing, and HTG EdgeSeq AMPure cleanup of Illumina Sequencing Libraries. Libraries concentration was evaluated by the HTG EdgeSeq KAPA Library quantification kit, and each library was normalized and pooled using the HTG EdgeSeq RUO library calculator. Then, pooled libraries were denatured in 2 N NaOH and sequenced (final concentration 4 pM) onto Illumina NextSeq550 platform (High Output kit v.2 cat. FC-404-2005). Data were returned from the sequencer as demultiplexed FASTQ files. The resulting reads (on average 79% passing filter, corresponding to a total of 8.33×10^8^ passing filter reads) were aligned referring to miRbase v.20 using HTG Parser software. Raw reads were standardized into CPM and filtered through the “limma Edge R” R Bioconductor package. For the QiaSeq Small RNA sequencing, total RNA extraction was performed from 200 µL of plasma through Serum/Plasma Norgen kit (cat. 55000). Small RNA Libraries were prepared using the QiaSeq miRNA library kit (cat. 331505) following the manufacturer’s instructions. QiaSeq strategy assigns Unique Molecular Index (UMI) during reverse transcription step to every mature miRNA molecule, in order to enable unbiased and accurate Small RNAome-wide quantification of mature miRNAs and additional small RNAs by NGS. Libraries quality control (QC) was performed by quantifying their concentration through QUBIT 3.0 spectrofluorometer (Qubit™ dsDNA HS Assay Kit, cat. Q32854) and assessing their quality using capillary electrophoresis in Bioanalyzer 2100 (Agilent High Sensitivity DNA kit cat. 5067-4626). The quality of libraries was evaluated considering electropherograms showing a peak comprised between 175 and 185 bp as high quality. Following QC, all libraries were normalized until 2 nM and pooled, denatured in 0.2 N NaOH, and further sequenced (final concentration 175 pM) using the Illumina NovaSeq 6000 platform (NovaSeq 6000 SP Reagent Kit (100 cycles) cat. 20027464, NovaSeq XP 2-Lane Kit cat. 20021664, using the XP protocol applying 75×1 single reads). Data were returned from the sequencer as demultiplexed FASTQ files. Resulting reads (on average 87% passing filter, corresponding to a total of 1.11×10^9^ passing filter reads) were mapped using QIAGEN Gene Globe data analysis center software, which adopts a sequential alignment strategy to map to different databases (perfect match to miRbase v.21 mature, miRBase hairpin, noncoding RNA, mRNA and other RNA, and ultimately a second mapping to miRBase mature, where up to two mismatches are tolerated) using bowtie (bowtie-bio.sourceforge.net/index.shtml). At each step, only mapped sequences were passed to the next step.

### Plasma Metabolomics (GCxGC-MS)

83 metabolites, covering various amino acids, fatty acids and sugars etc. (Supplementary Table 3), was generated from baseline plasma samples as described before^26^. In brief, 30 μL blood samples from 93 participants were spiked with 10 μL of the internal standard mixture (d4-succinic acid, d5-glutamic acid, d8-valine, and d33-heptadecanoic acid.; Sigma Aldrich). Samples were vortex-mixed and incubated on ice for 30 min and centrifuged (10,000 rpm, 3 min, 4◦C). Finally, 180 µL of the filtered extracts were transferred to glass vials and evaporated dry before derivatization.

The samples were derivatized using a previously described procedure^27^, where reactive groups are converted into trimethysilyl derivates, which increases the volatility of the biomolecules. The polar metabolites were then analyzed using a Pegasus 4D (LECO; Saint Joseph; USA) system, which combines two-dimensional chromatographic separation with time-of-flight (TOF) mass spectrometric detection. Identifications were assigned using the National Institute of Standards (NIST) database and Steno Diabetes Center Copenhagen in-house libraries. After data acquisition, the raw data were pre-processed into a peak table with ChromaTOF (LECO; Saint Joseph; USA). Finally, data were post-processed in R by batch correction, truncation of outliers, and imputation of missing values.

### Plasma Lipidomics (LC-MS)

The lipidome (Supplementary Table 4) was measured from baseline plasma samples of 94 individuals and two unrelated control samples in four replicates following lipid extraction from a 10 μL plasma sample using a chloroform:methanol (2:2 V/V) lipid extraction method^28^. Nine stable isotope labelled and non-physiological lipid species were spiked as internal standards. (Supplementary Table 5).

Samples were analysed in positive and negative ion modes of ultra-high-performance liquid-chromatography mass-spectrometry (UHPLC-MS; Agilent Technologies; Santa Clara, CA, USA) at Steno Diabetes Center Copenhagen as described previously^29–30^. After data acquisition, the raw data were pre-processed into a peak table with MZmine 2. Finally, data were post-processed in R by denoising as normalization to internal standards, batch correction, truncation of outliers, and imputation of missing values.

Nine stable isotope labelled and non-physiological lipid species were spiked as internal standards (Supplementary data table 2).

### PBMC (cryopreserved) multi-dimensional flow cytometry (Multi-FACS) immunomics

The immunome was examined using cryopreserved peripheral blood mononuclear cells (PBMCs; that were freshly isolated and cryopreserved in Cryostor) from 76 baseline blood samples (Na-heparin) shipped in liquid nitrogen and processed at a single site by Multi-FACS. 36-marker Multi-FACS Cytek Aurora panel (plus one viability dye) was set up and a specific flow gating strategy that allowed enumeration of 150 cell populations was applied (Supplementary Table 6).

All flow data were also analyzed by OMIQ platform (www.omiq.ai), including unbiased clustering (FlowSOM) and dimensionality reduction (UMAP). In FlowSOM analysis, the median intensity of 36 markers were used to generate 100 nodes, which were further clusters as meta-cluster (k=30, grey area on FlowSOM map under randomized speed and Euclidean distancing metric). FlowSOM map provides a concise representation of the number of cell types and visualization of the differential marker profiles by color-density.

Samples were processed in five batches (between 12 to 16 samples per batch, consisting of a mixture of samples from each of the five INNODIA immune laboratories) together with two unrelated control samples in each batch using one 2.5×10^6^ PBMCs per sample except for two samples where two aliquots were used due to poor percentage of recovery (range from 11.2 to 228.8%, average of 68.1%). Viability assessed by trypan blue ranged between 70 and 100% (average 94%). An average of 1.51×10^6^ (range 0.54-2.8×10^6^) cells were used for staining. List of Multi-FACS flow panel antibodies and reagents used is detailed in Supplementary data table 3. PBMCs were first stained using Live/dead blue for 15 min at room temperature, washed with FACS buffer (PBS with 0.2% BSA and 2mM EDTA), and incubated with Fc receptor blocker (TruStain FcX Fc; BioLegend) for 10 min at room temperature. Without wash, samples were stained in a 37°C waterbath for 15 min using mastermix 1 (containing antibodies against CXCR3. CD117, CD294/CRTH2, and CD161). Samples were further stained in waterbath for 15 min using mastermix 2 (containing antibodies against CXCR5, ICOS, CCR7, and CCR6), followed by 30 min at room temperature using mastermix 3 (Supplementary data table 3). Finally, samples were washed using FACS buffer, then fixed and resuspended in PBS containing 1% paraformaldehyde (Alfa Aesar). Single colour controls were made using PBMC for all colours except for CD294, CD117, CD161, and TCRgd where BD mouse or rat comp beads were used instead due to low cell expression. Single colour controls were subjected to the same buffer and fixed as the multi-colour stained samples. SpectroFlo QC beads were run daily and single colour controls were acquired in the reference library, which was subsequently used for live unmixing during sample acquisition on a Cytek Aurora cytometer. Flow data were analysed using FlowJo software (an example of the gating strategy is shown in Supplementary Fig. 8 and checked by an independent reviewer).

### Multi-omics data pre-processing, integration, and analysis

For transcriptomics and miRNA data, the DESeq 2 package^31^ was used to normalize the counts using the variance stabilizing transformation (VST). The transcriptomics and miRNA data sets were filtered for low counts (features with less than 10 counts in total or features with zero counts in more than 90% of the samples). The proteomics, metabolomics, lipidomics and immunomics data sets were log2 transformed. The transcriptomics, proteomics, and immunomics data were corrected for batch effects associated with dataset-specific factors (sequencing at different days or different handling of the samples) using the limma package in R. Finally, all the data sets were corrected for age. The age was log-transformed to account for the growth effect in children (one year difference in adults is not equivalent to one year difference in children). This was necessary due to the high degree of age heterogeneity present in the cohort and the association of the fasted C-Peptide slopes with age.

The omics data sets were integrated using the general framework in the Multi-Omics Factor Analysis (MOFA) package from 2018^32^. MOFA performs a dimensionality reduction of the omics data into a lower-dimensional latent space (Fig. 1C). The latent factors generated by MOFA capture sources of global variability across the different omics data sets. Each factor has an underlying weight for every feature, which can be used to annotate the factors in terms of omics features, yielding a specific molecular signature for each factor. MOFA was run with default parameters and 20 latent factors. The model was initialized with different random seeds yielding similar results, generally only altering the number assigned to each factor associated with the C-peptide slopes.

Latent factors were associated with the C-peptide slopes using the Spearman correlation. Other covariates were also analyzed such as age at baseline and C-peptide at baseline. The association of the latent factors with the progression groups was calculated using the Kruskal-Wallis test and each group was compared using a Mann-Whitney U test.

Gene Set Enrichment Analysis (GSEA)^33^ was performed in order to better characterize the genes with the largest weight in the latent factors. This analysis was performed using the MOFA GSEA function that utilizes a modified version of the principal component gene set enrichment scheme (PCGSE)^34^. The Reactome database was the gene set annotation used for this analysis. The GSEA was performed separately for genes with a positive and negative weight in each latent factor. This was done to avoid combining genes that are upregulated (positive weight) and downregulated (negative weight) in the latent factor, as these two groups of genes might be involved in different biological pathways. The top 15 significant pathways for each latent factor (positive and negative weights) were selected and grouped by biological pathway.

Differential gene expression of individual genes was determined with the DESeq 2 package^31^. Models included covariates for the batch variable (different runs) and the age of the people. Age was encoded as a categorical variable defined in three groups, less than ten years, between ten and 18 years, and more than 18 years. Differential gene expression was assessed between rapid and slow groups, rapid and increasing and slow and increasing, respectively.

For biological network analysis, two types of interaction networks were constructed. One was compiled from the STRING^35^ database to study protein-protein interactions or associations only. Another network was compiled from the mirTarBase^36^ (miRNA-gene), which was combined with the STRING network to study protein-protein-miRNA interactions or associations. The STRING database was filtered for high-confidence interactions (combined score above 0.7) and miRNA-gene interactions had to be reported by at least two publications and two non-high-throughput methods. We decided to focus on genes and miRNAs only because those were the omics types with higher weights in the most relevant latent factors.

The association between the biological networks and the latent factors was performed using the PCSF graph optimization approach. This method allows us to interpret the biological landscape of the interaction network based on the importance/weight of each gene/miRNA in the latent factor. The output is a subnetwork that captures interactions between the genes/miRNAs with a higher importance in the latent factor. In order to select a subset of genes and miRNAs to construct the network, only genes with normalized absolute weights three-fold higher than expected by chance and miRNAs with normalized absolute weights two-fold higher than expected by chance were selected. Grid-search was performed to select the best parameters based on the network that had a high number of genes/miRNA from the latent factor while keeping number of genes/miRNA not observed in the latent factor low (μ=0.005, χο=1, β= 5000). The final network was constructed using 20 runs with noise to edge costs (r=0.1) that were combined and clustered using the edge-betweenness algorithm. Gene set enrichment analysis was performed on the clusters obtained from the network.

### Other

#### Data Availability

The data generated and analysed is person-sensitive as it can be used to identify people based on their sequence variation and can be accessed in secure environments only. Access to data can be provided by application to the INNODIA Data Access Committee by emailing Professor Lut Overbergh. Processed results of GSEA analysis is available as supplementary material.

#### Code Availability

The primary software used for this work was Multi-Omics Factor Analysis (MOFA). The code for this software is available at https://github.com/bioFAM/MOFA2.

#### Ethics Declaration

The INNODIA study protocol was approved by the London – City & East Research Ethics Committee on 28 October 2016 (REC 16/LO/1750) IRAS Project ID 210497. Subsequently, after translation of the participants’ documentation, approval was obtained from local Ethic authorities throughout the entire INNODIA clinical network.

## Data Availability

The data generated and analysed is person-sensitive and can be accessed in secure environments only. Access can be provided by application to the INNODIA Data Access Committee.

## Acknowledgments

We gratefully acknowledge all participants of the INNODIA natural history study which built the basis for the work presented here. Without the outstanding engagement of the people with type diabetes and their relatives and friends participating in the INNODIA clinical efforts, this data and new knowledge gain would not have been possible. The authors also want to thank all the clinical personnel for their dedication in the participant recruitment, characterization, sample collection and preparation that was at the basis of this analysis. We dedicate this work to the late Professor David Dunger, who inspired INNODIA and lay at the basis of this analysis.

## Author contributions

JJAA, CB, SFAB, DBD, PJC, MK, AMS, GS, LLE, FD, TT, RL, LO, CM, MP, SB designed the study. JJAA, CB, KB, RM, NA-S, IM, CL-Q, TS, JHL, GEG, MKH, TS, OR, PJC, JHL, GS, TT collected data and performed analyses. JJAA, CHJ, TT, LO, CM, MP, SB wrote the manuscript. All authors revised the manuscript for crucial content and approved the final version. All authors had full access to all the data and had final responsibility for the decision to submit for publication. A complete list of the partners of the INNODIA consortium is included separately.

## Declaration of interests

CM serves or has served on the advisory panel for Novo Nordisk, Sanofi, Merck Sharp and Dohme Ltd., Eli Lilly and Company, Novartis, AstraZeneca, Boehringer Ingelheim, Roche, Medtronic, ActoBio Therapeutics, Pfizer, Imcyse, Insulet, Zealand Pharma, Avotres, Mannkind, Sandoz and Vertex. Financial compensation for these activities has been received by KU Leuven; KU Leuven has received research support for CM from Medtronic, Imcyse, Novo Nordisk, Sanofi and ActoBio Therapeutics; CM serves or has served on the speakers bureau for Novo Nordisk, Sanofi, Eli Lilly and Company, Boehringer Ingelheim, Astra Zeneca and Novartis. Financial compensation for these activities has been received by KU Leuven. S.Br. reports having received funding from INNODIA (grant agreement No 115797), having ownerships in Intomics A/S, Hoba Therapeutics Aps, Novo Nordisk A/S, Lundbeck A/S, ALK A/S and managing board memberships in Proscion A/S and Intomics A/S. MK reports ownership and managing board membership in Vactech Oy. CLQ reports serving on advisory boards of Fondation Alzheimer’s and Institute Pasteur, and an affiliation with Novo Nordisk. LO reports having received funding from INNODIA and INNODIA Harvest.

## Supplementary information

Supplementary information is available for this paper as an appendix.

## Notes

### Funding Statement

This project has received funding from the Innovative Medicines Initiative 2 Joint Undertaking under grant agreement No 115797 (INNODIA) and No 945268 (INNODIA HARVEST). This Joint Undertaking receives support from the Union's Horizon 2020 research and innovation programme, "EFPIA", "JDRF" and "The Leona M. and Harry B. Helmsley Charitable Trust". Novo Nordisk Foundation grants (NNF17OC0027594 and NNF14CC0001) to S.B. are also acknowledged.

### Author Declarations

NHS Health Research Authority London - City & East Reasearch Ethics Committee gave ethical approval for this work (IRAS Project ID 210497) Subsequently, after translation of the participants' documentation, ethical approval was obtained from the local Ethic authorities throughout the entire INNODIA clinical network listed below. Commissie Medische Ethiek UZ KU Leuven / Onderzoek ga Ethikkommission Medizinische Universitat Graz Center for sundhed Videnskabsetisk Komite COMITE DE PROTECTION DES PERSONNES SUD MEDITERRANEE III Helsingen Ja Uudenmaan Sairaanhoitopiiri MHH Ethikkommission OE 9515 Ethik-Kommission der Bayerischen Landesarztenkammer Uulm Ethikkommission Comitato Etico Regionale per la Sperimentazione Clinica della Regione Toscana Comitato Etico delle Province di Chieti e Pescara Ospedala San Raffaele, il Comitato Etico Il comitato Etico Bambino Gesu Ospedale Pediatrico Comite National d`ethique de recherche Regional Ethics Committee Ethics Committee of Silesia in Katowice Republic of Slovenia, THE NATIONAL MEDICAL ETHICS COMMITTEESökande forskningshuvudman, Region Skåne

