## Supplementary figures and images for "Multi-omics analysis reveals drivers of loss of β-cell function after newly diagnosed autoimmune type 1 diabetes: An INNODIA^‡^ multicenter study"

### Supplementary Figure 1

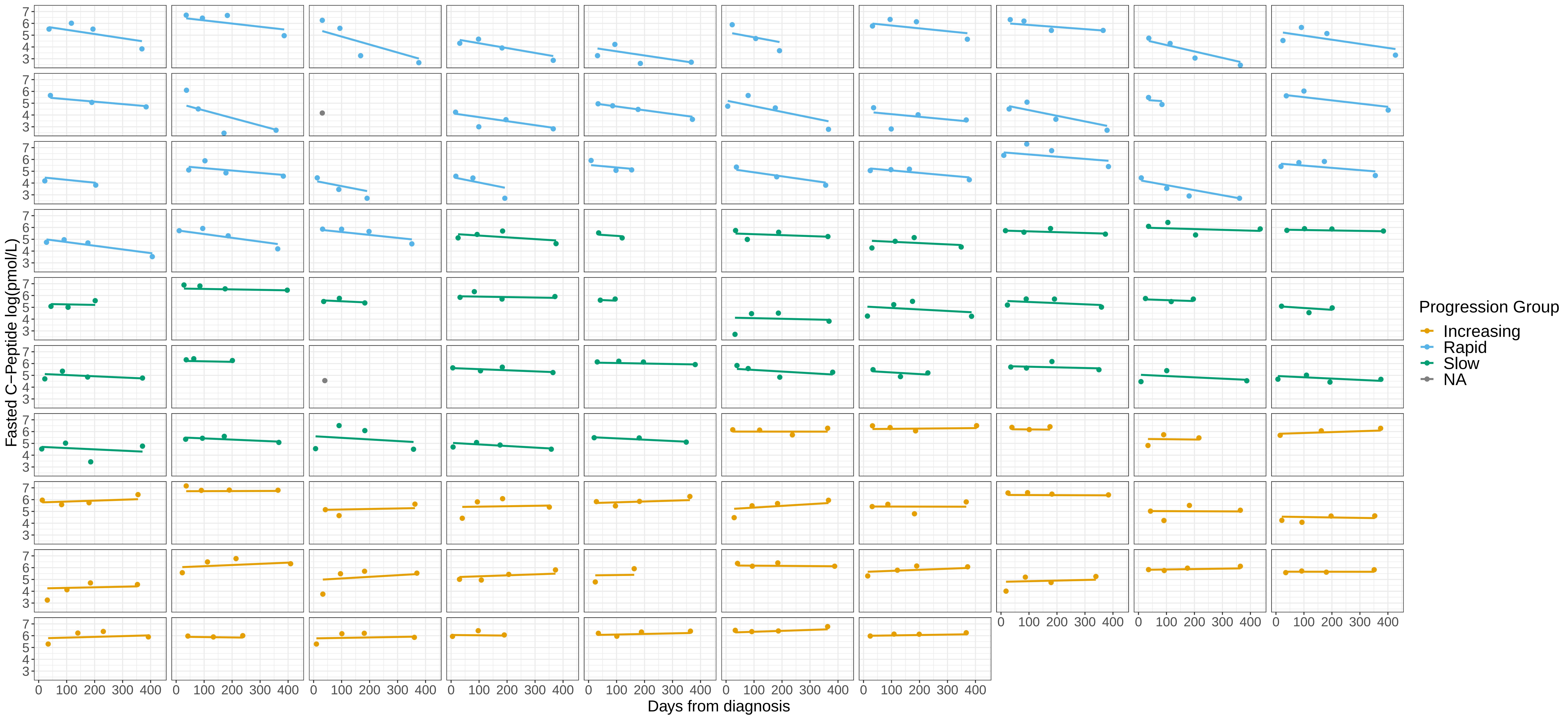

### Supplementary Figure 2

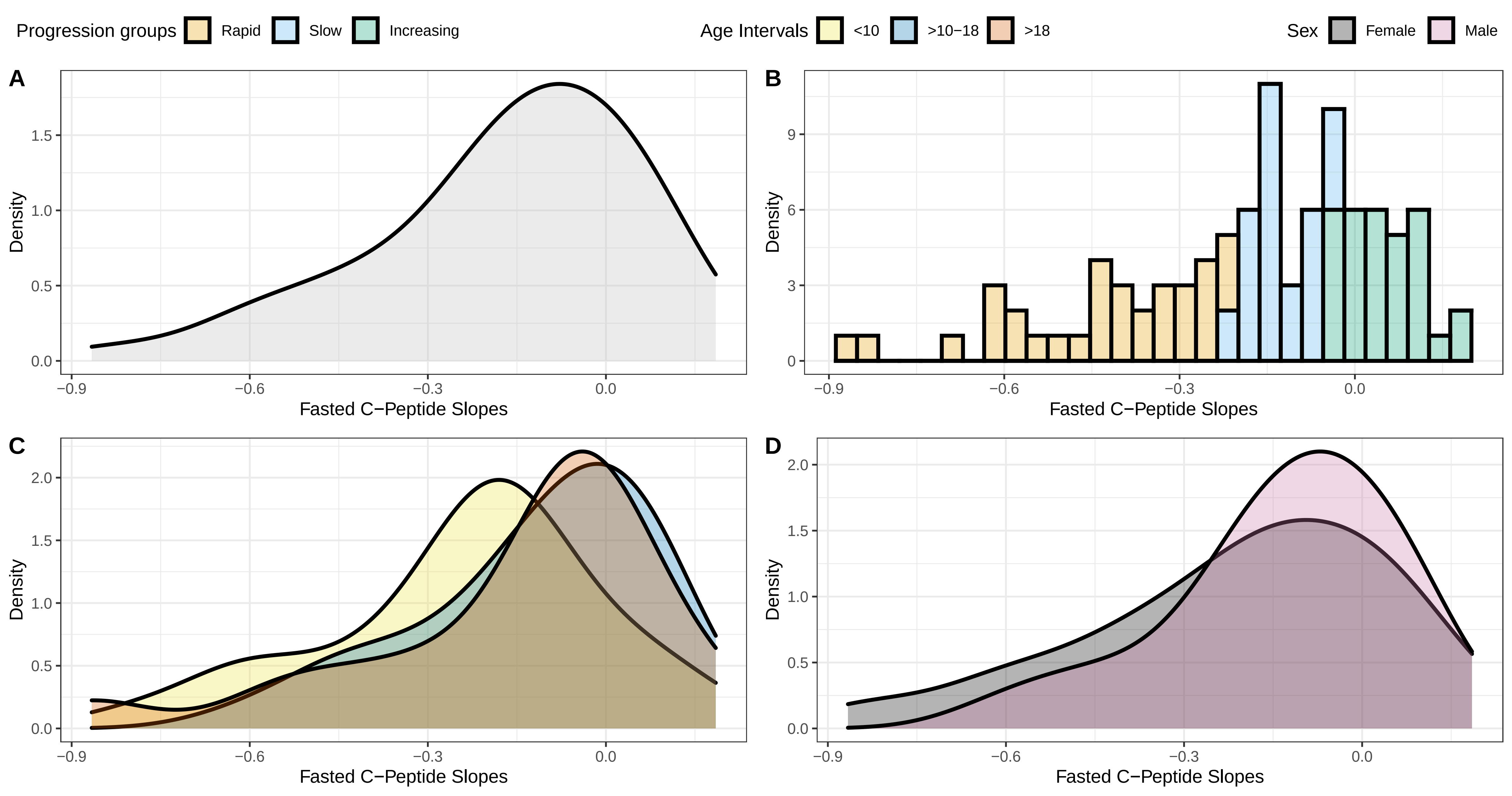

### Supplementary Figure 3

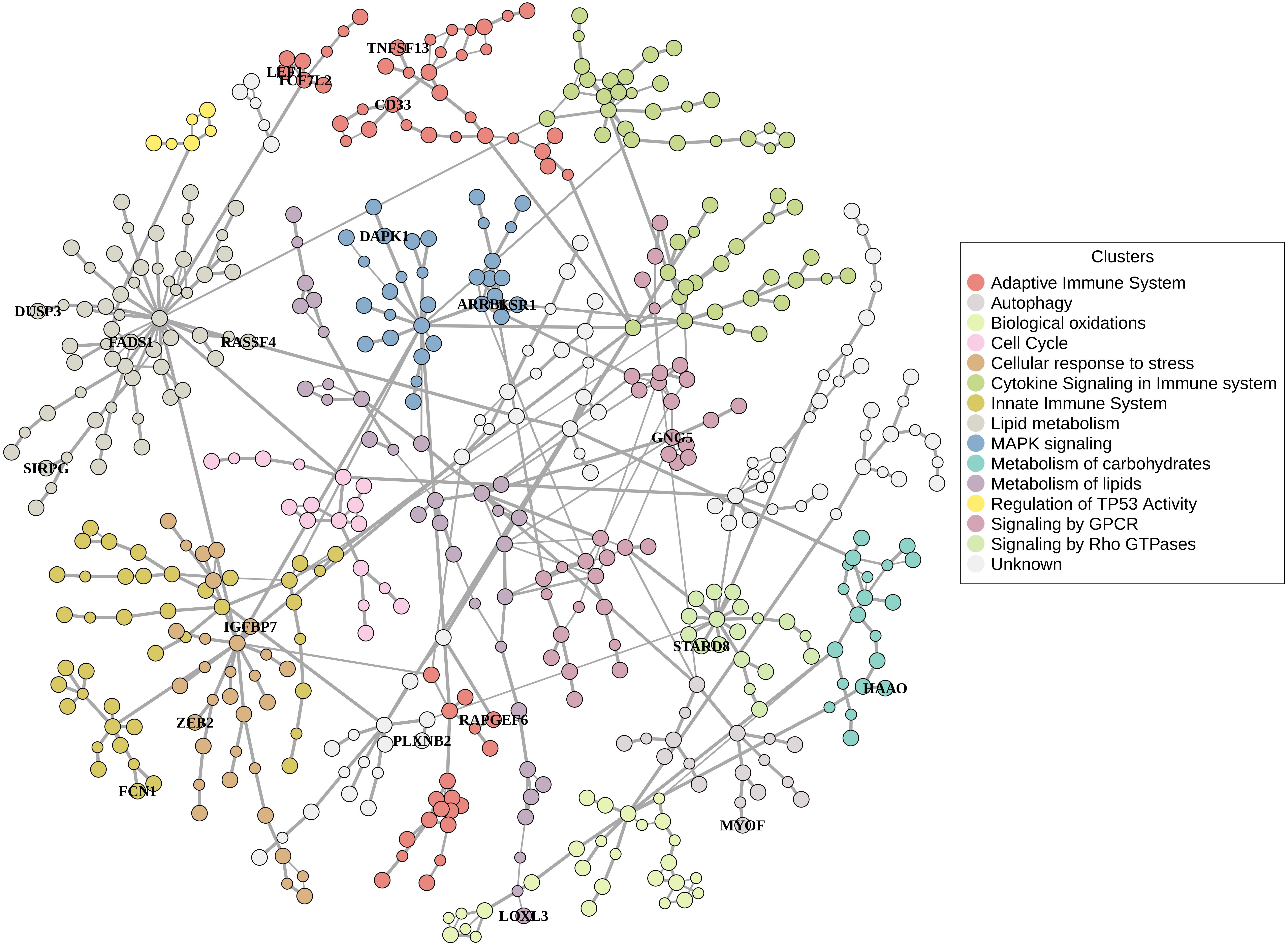

### Supplementary Figure 4

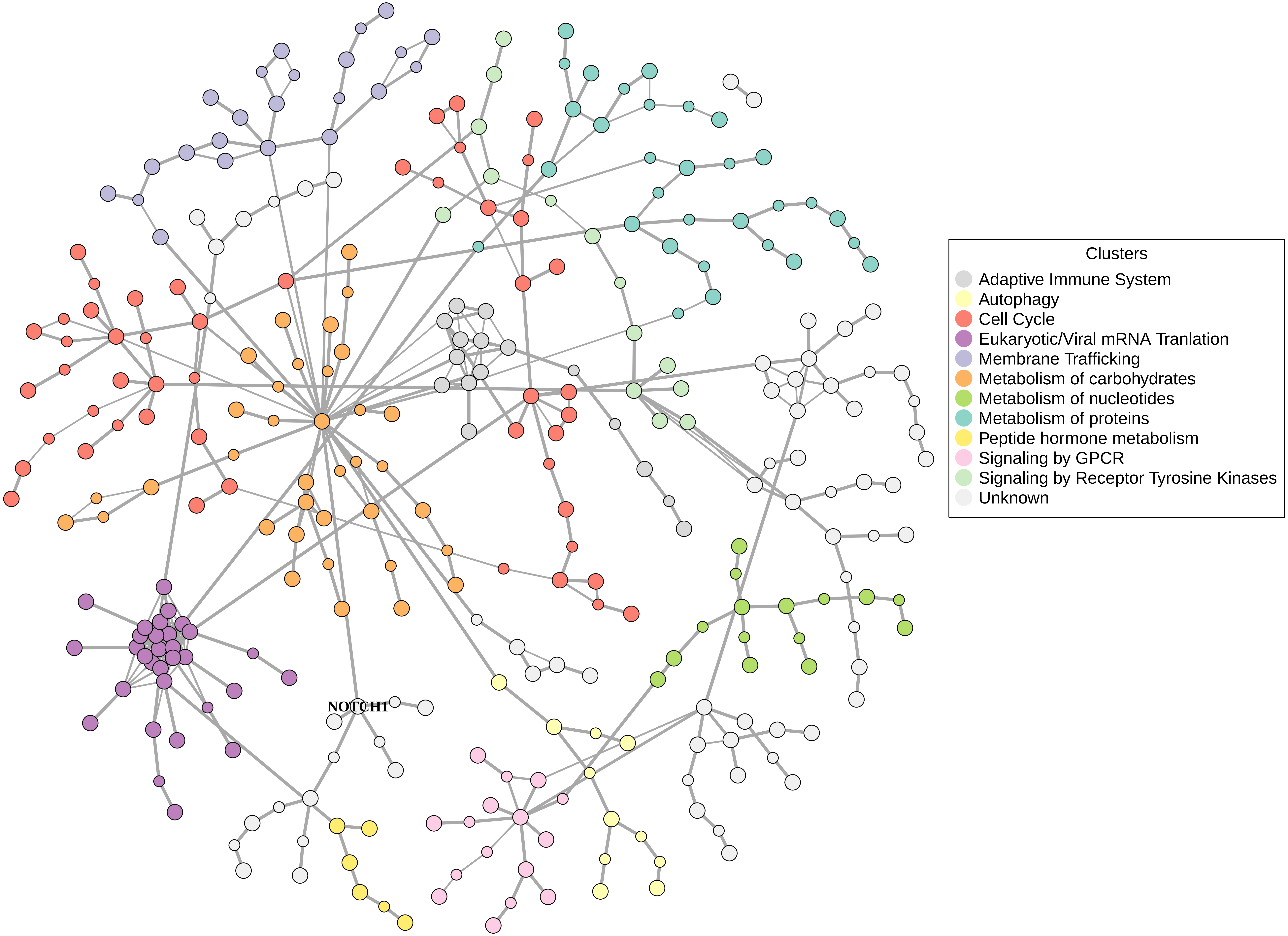

### Supplementary Figure 5

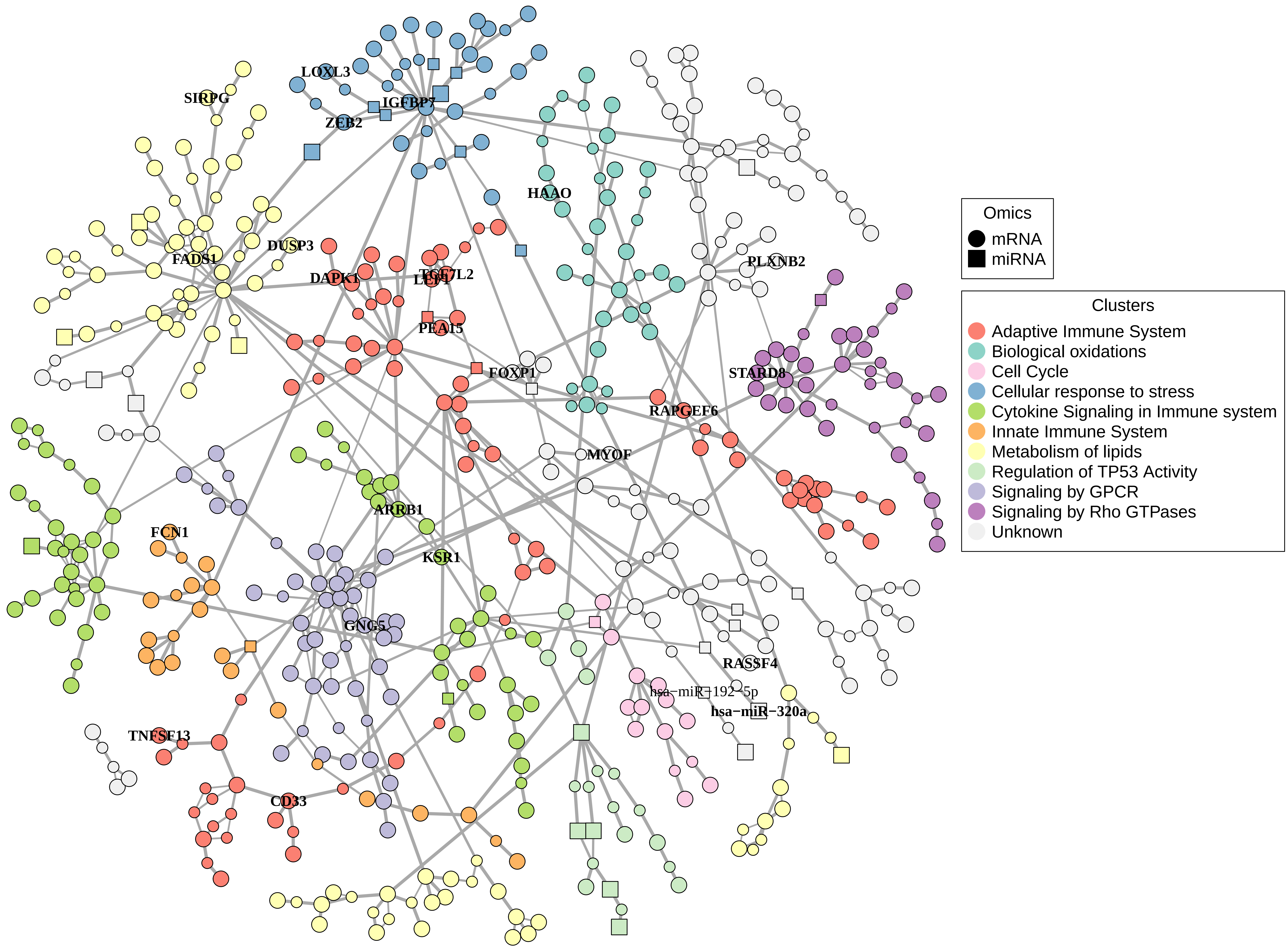

### Supplementary Figure 6

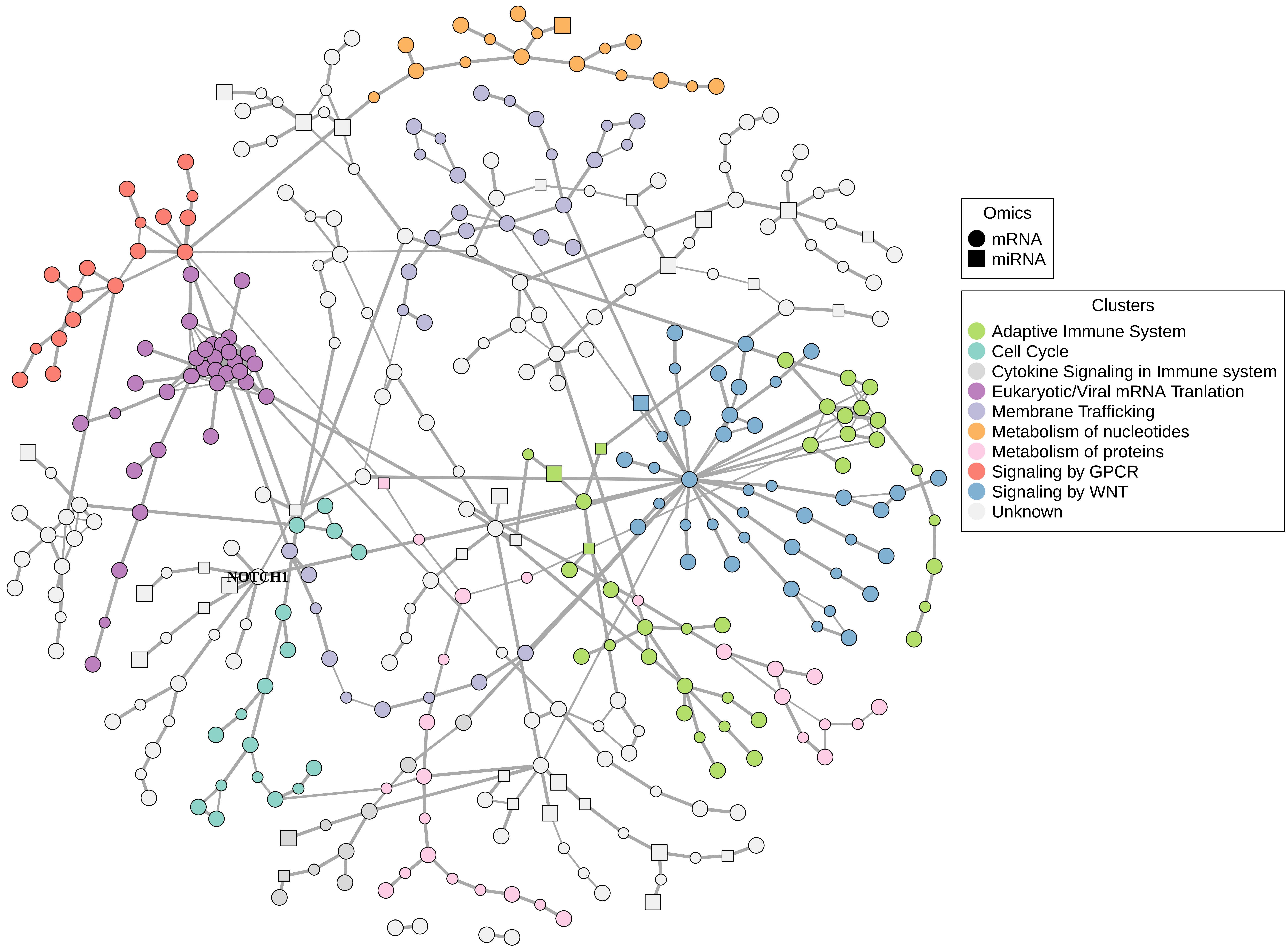

### Supplementary Figure 7

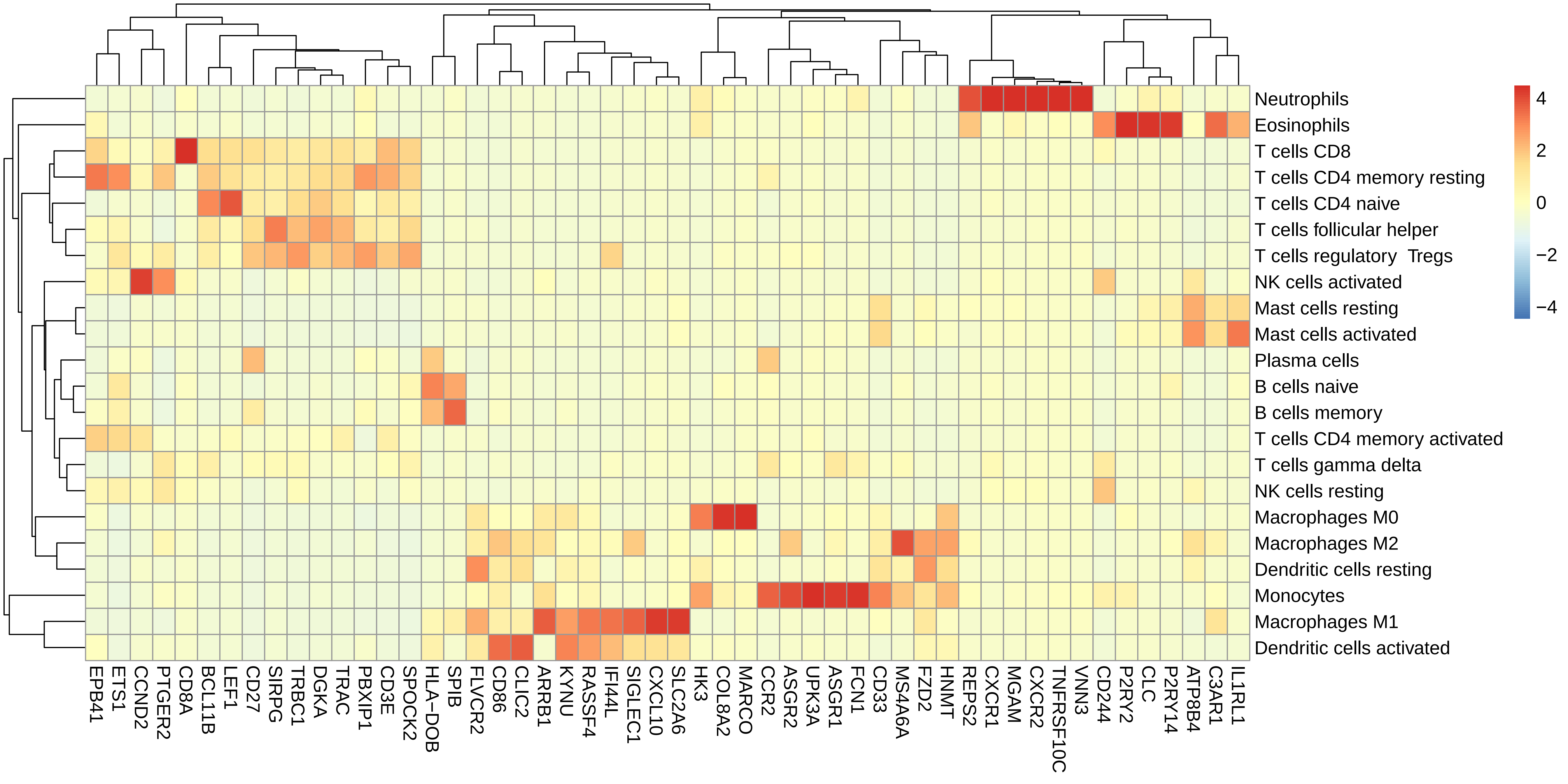
