## Supplementary Figure 8 for "Multi-omics analysis reveals drivers of loss of β-cell function after newly diagnosed autoimmune type 1 diabetes: An INNODIA^‡^ multicenter study"

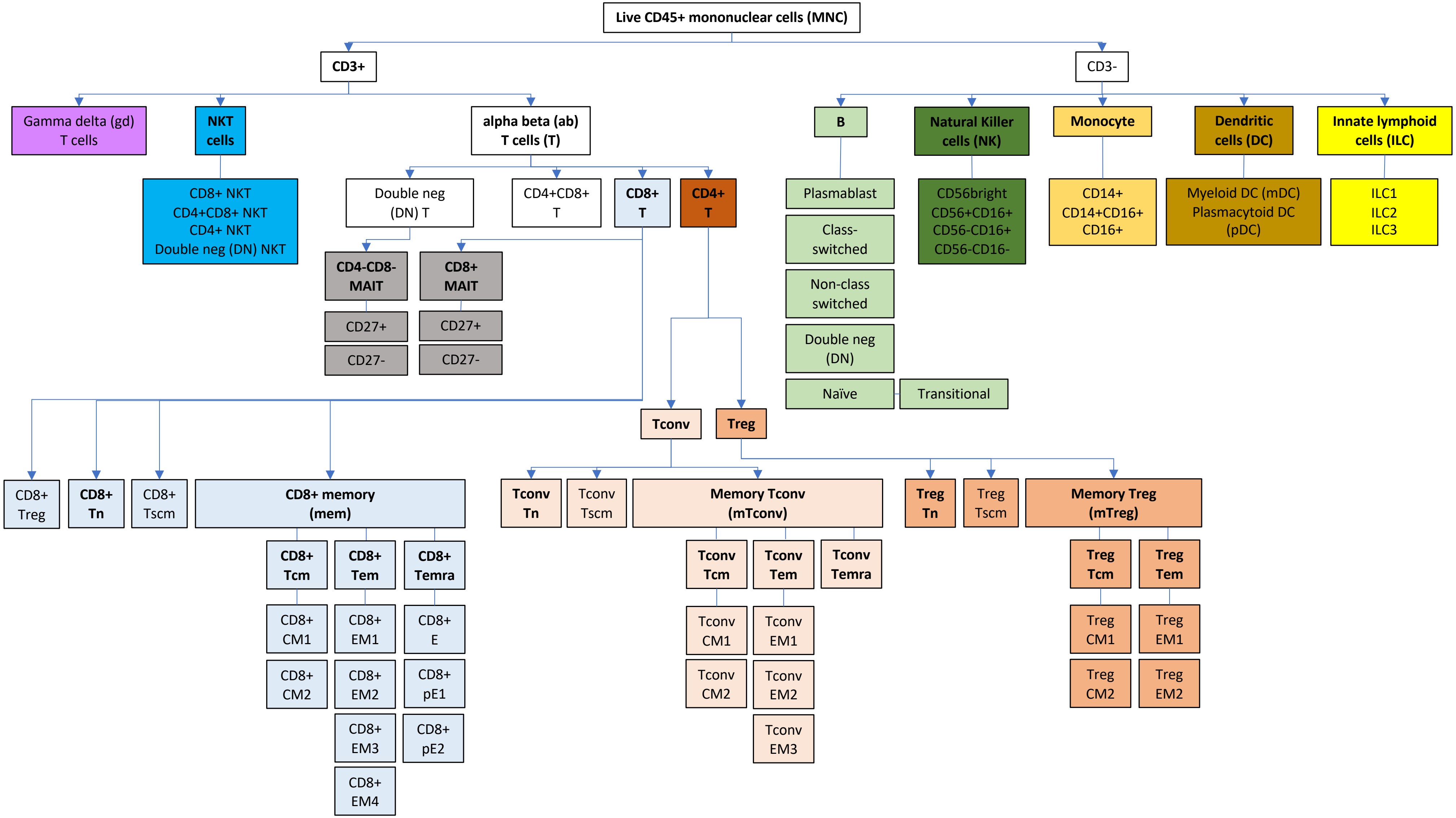
Live CD45+ mononuclear cells (MNC)
CD3+
CD3-
Gamma delta (gd) T cells
NKT cells
alpha beta (ab) T cells (T)
B
Natural Killer cells (NK)
Monocyte
Dendritic cells (DC)
Innate lymphoid cells (ILC)
CD8+ NKT
CD4+CD8+ NKT
CD4+ NKT
Double neg (DN) NKT
Double neg (DN) T
CD4+CD8+ T
CD8+ T
CD4+ T
Plasmablast
Class-switched
Non-class switched
Double neg (DN)
Naïve
Transitional
CD56bright
CD56+CD16+
CD56-CD16+
CD56-CD16-
CD14+
CD14+CD16+
CD16+
Myeloid DC (mDC)
Plasmacytoid DC (pDC)
ILC1
ILC2
ILC3
CD4-CD8- MAIT
CD8+ MAIT
CD27+
CD27-
CD27+
CD27-
Tconv
Treg
CD8+ Treg
CD8+ Tn
CD8+ Tscm
CD8+ memory (mem)
CD8+ Tcm
CD8+ Tem
CD8+ Temra
CD8+ CM1
CD8+ CM2
CD8+ EM1
CD8+ EM2
CD8+ EM3
CD8+ EM4
CD8+ E
CD8+ pE1
CD8+ pE2
Tconv Tn
Tconv Tscm
Memory Tconv (mTconv)
Tconv Tcm
Tconv Tem
Tconv Temra
Tconv CM1
Tconv CM2
Tconv EM1
Tconv EM2
Tconv EM3
Treg Tn
Treg Tscm
Memory Treg (mTreg)
Treg Tcm
Treg Tem
Treg CM1
Treg CM2
Treg EM1
Treg EM2

#### Activation, exhaustion and senescence markers

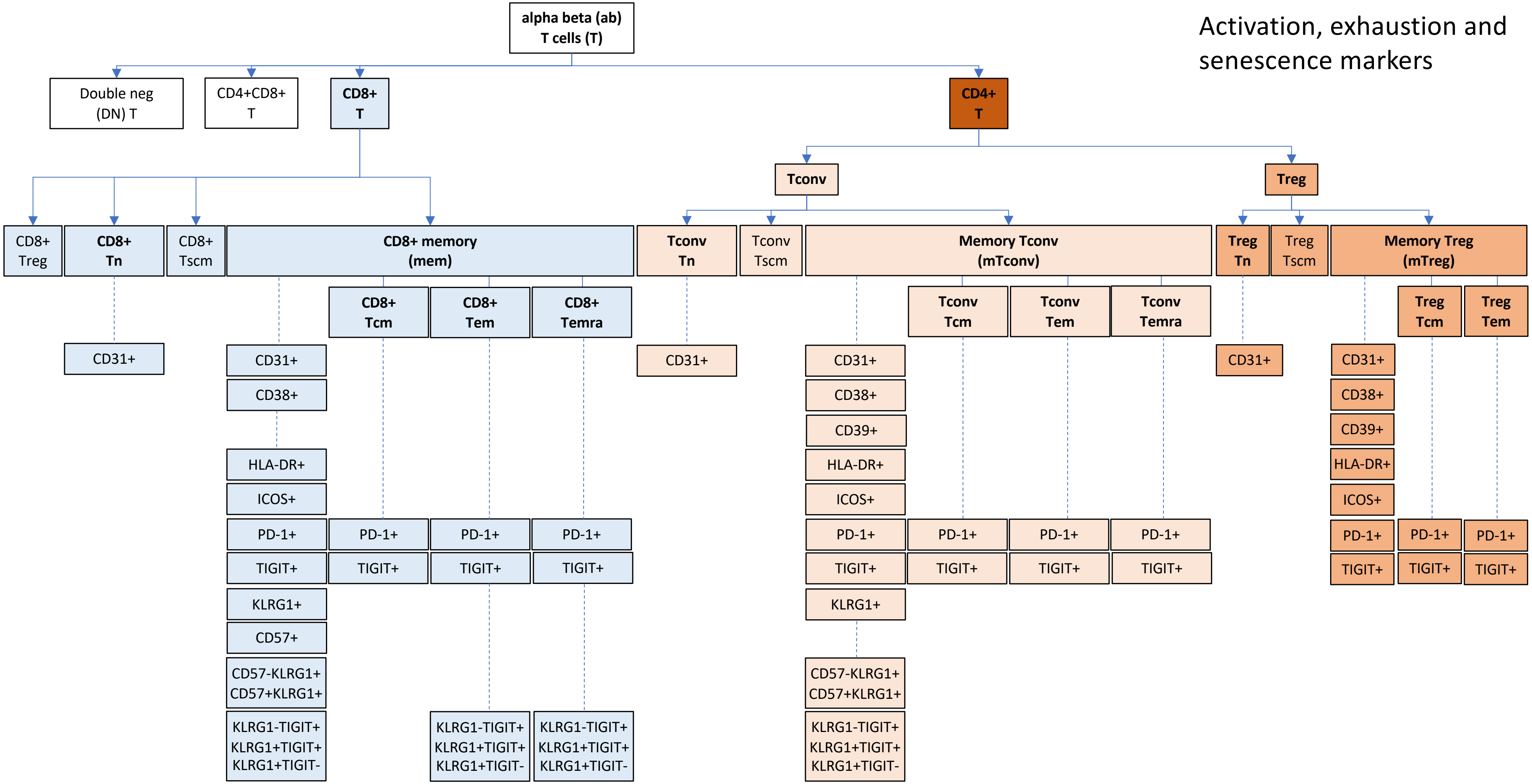

#### Helper T cells and Helper T cell-like Tregs

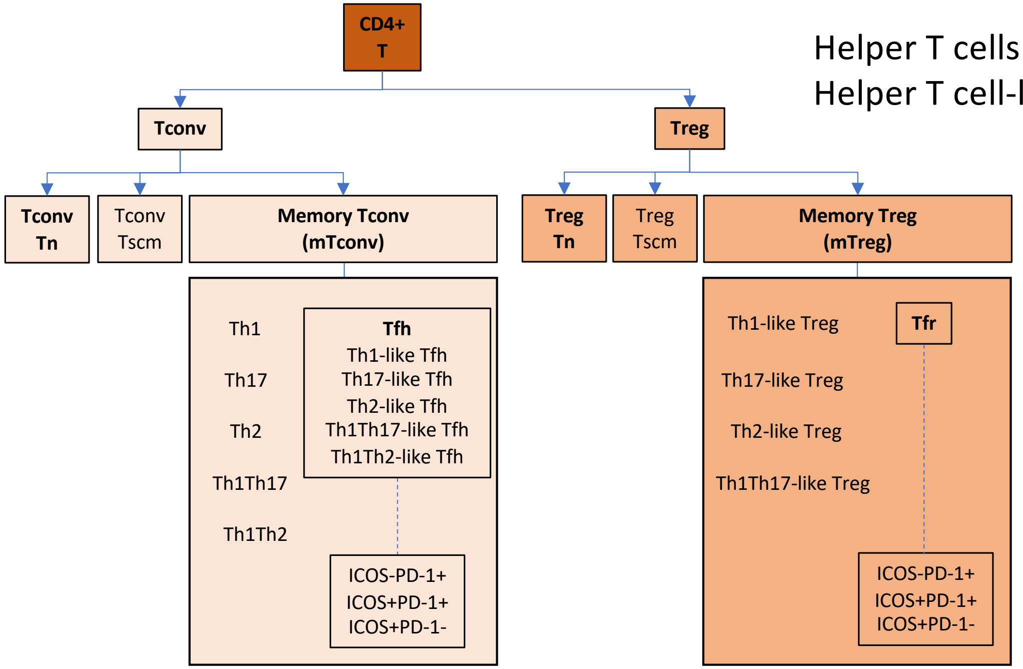

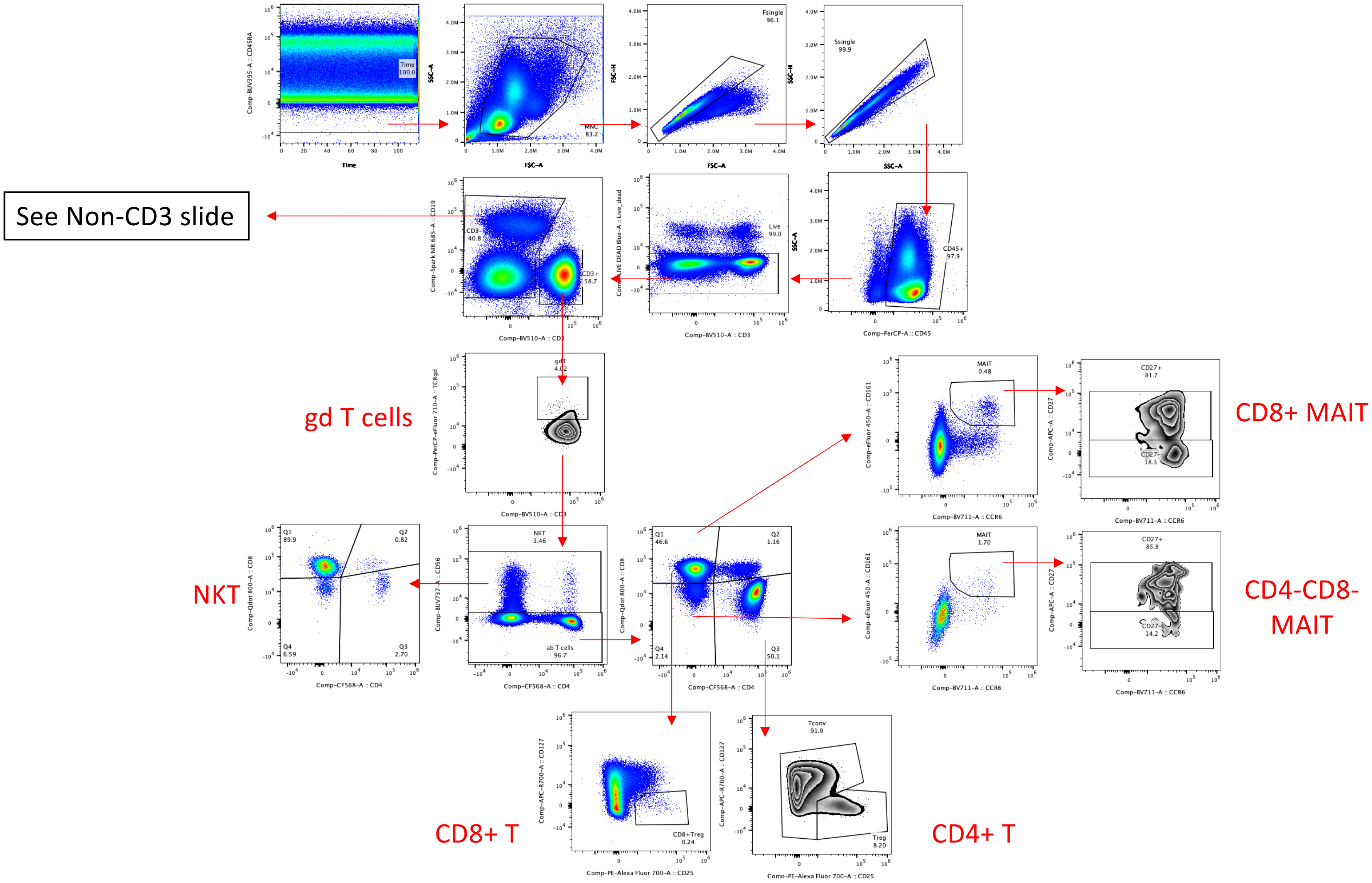
See Non-CD3 slide
gd T cells
NKT
CD8+ MAIT
CD4-CD8-
MAIT
CD8+ T
CD4+ T
Time
MNC
83.2
Fsingle
96.1
Ssingle
99.9
CD45+
97.9
Live
99.0
CD3-
40.8
CD3+
58.7
gdT
4.02
NKT
3.46
ab T cells
96.7
Q1
89.9
Q2
0.82
Q3
2.70
Q4
6.59
Q1
46.6
Q2
1.16
Q3
50.1
Q4
2.14
MAIT
0.48
MAIT
1.70
CD27+
81.7
CD27-
18.3
CD27+
85.8
CD27-
14.2
CD8+Treg
0.24
Tconv
91.9
Treg
8.20
Comp-BV510-A :: CD3
Comp-PerCP-A :: CD45
Comp-CF568-A :: CD4
Comp-BV711-A :: CCR6
Comp-PE-Alexa Fluor 700-A :: CD25

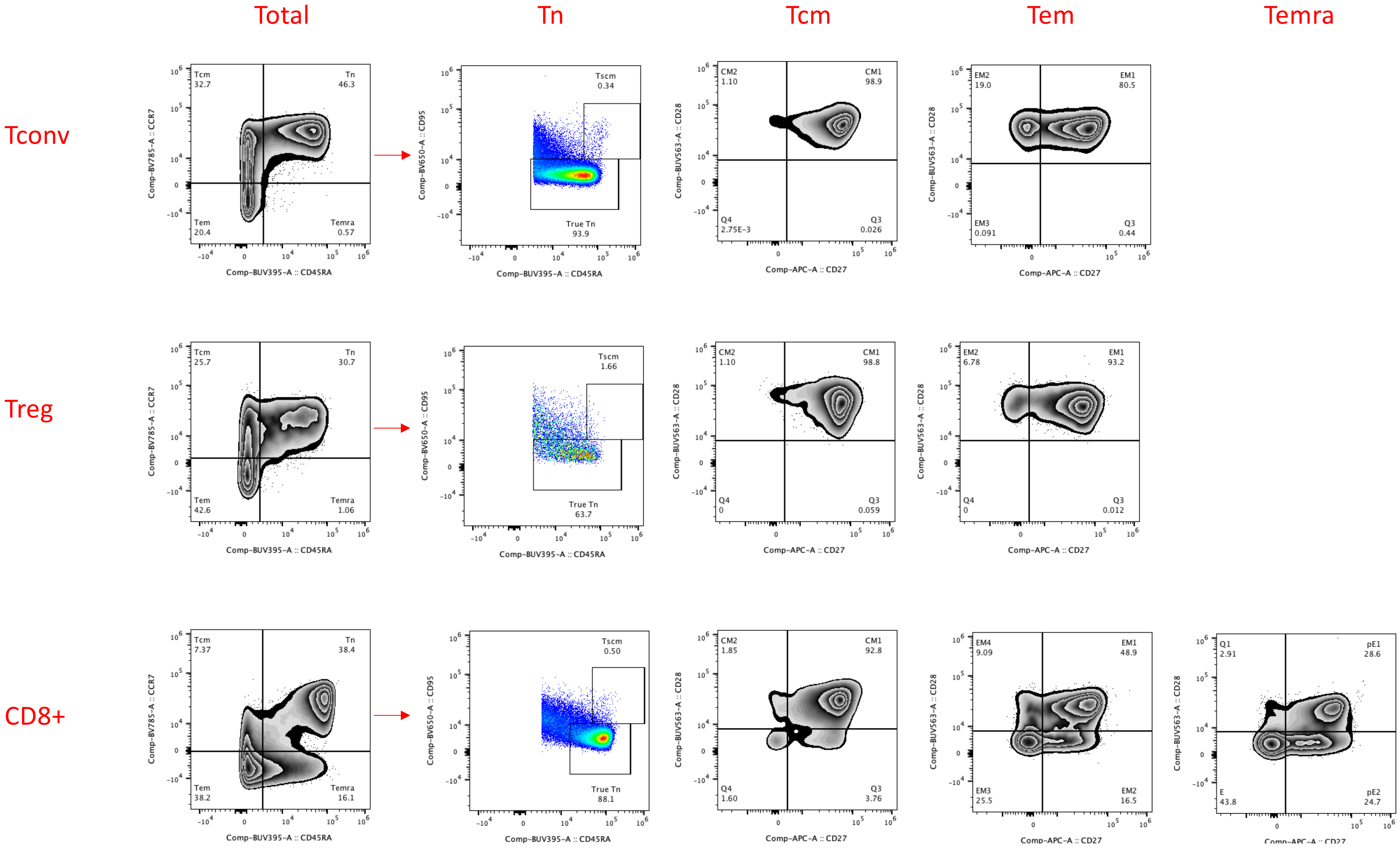
Total
Tn
Tcm
Tem
Temra
Tconv
Treg
CD8+
Tcm 32.7
Tn 46.3
Tem 20.4
Temra 0.57
Comp-BV785-A :: CCR7
Comp-BUV395-A :: CD45RA
Tscm 0.34
True Tn 93.9
Comp-BV650-A :: CD95
CM2 1.10
CM1 98.9
Q4 2.75E-3
Q3 0.026
Comp-BUV563-A :: CD28
Comp-APC-A :: CD27
EM2 19.0
EM1 80.5
EM3 0.091
Q3 0.44
Tcm 25.7
Tn 30.7
Tem 42.6
Temra 1.06
Tscm 1.66
True Tn 63.7
CM2 1.10
CM1 98.8
Q4 0
Q3 0.059
EM2 6.78
EM1 93.2
Q4 0
Q3 0.012
Tcm 7.37
Tn 38.4
Tem 38.2
Temra 16.1
Tscm 0.50
True Tn 88.1
CM2 1.85
CM1 92.8
Q4 1.60
Q3 3.76
EM4 9.09
EM1 48.9
EM3 25.5
EM2 16.5
Q1 2.91
pE1 28.6
E 43.8
pE2 24.7

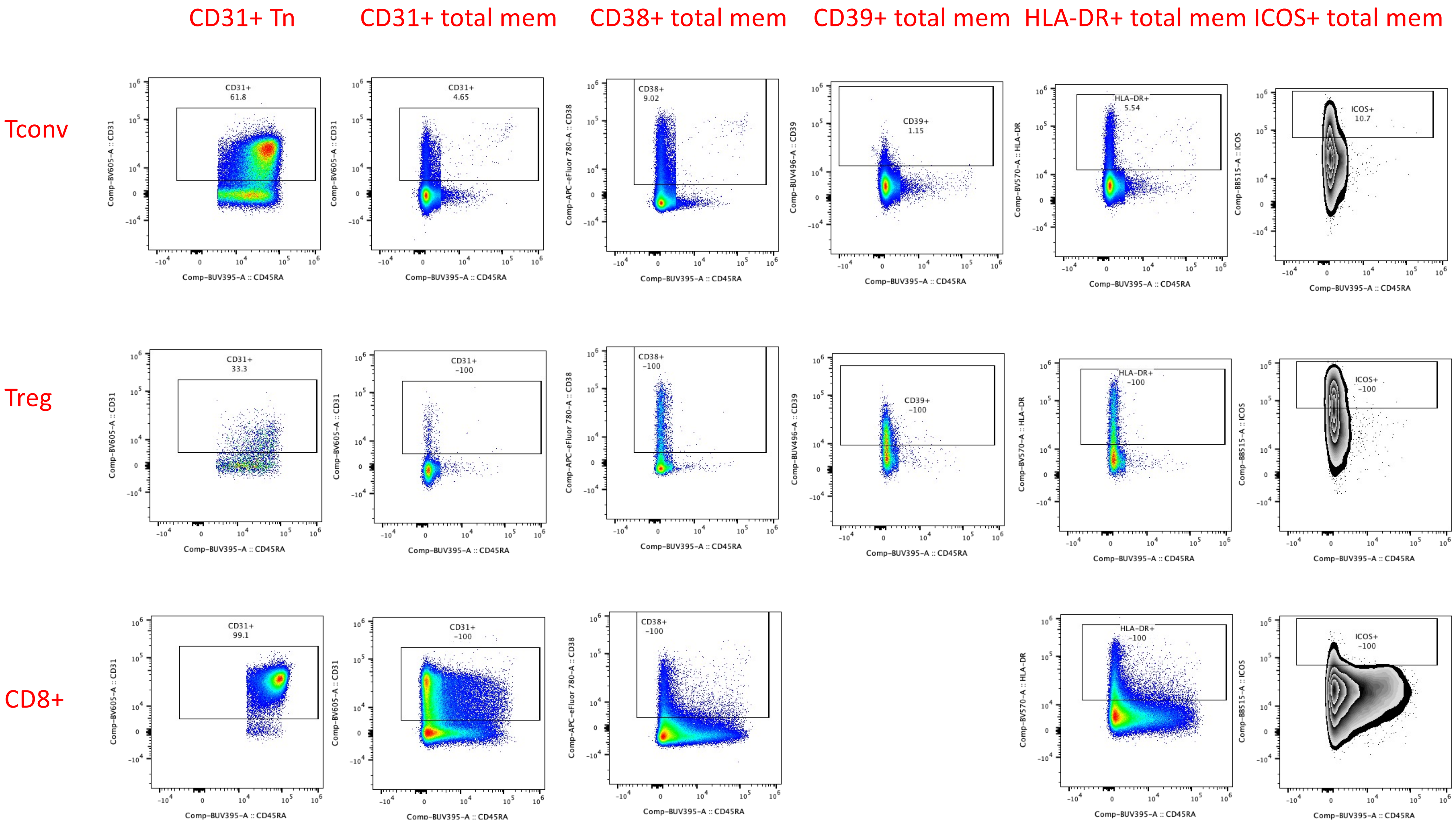
CD31+ Tn
CD31+ total mem
CD38+ total mem
CD39+ total mem
HLA-DR+ total mem
ICOS+ total mem
Tconv
Treg
CD8+
CD31+
61.8
4.65
CD38+
9.02
CD39+
1.15
HLA-DR+
5.54
ICOS+
10.7
33.3
~100
99.1
Comp-BV605-A :: CD31
Comp-APC-eFluor 780-A :: CD38
Comp-BUV496-A :: CD39
Comp-BV570-A :: HLA-DR
Comp-BB515-A :: ICOS
Comp-BUV395-A :: CD45RA

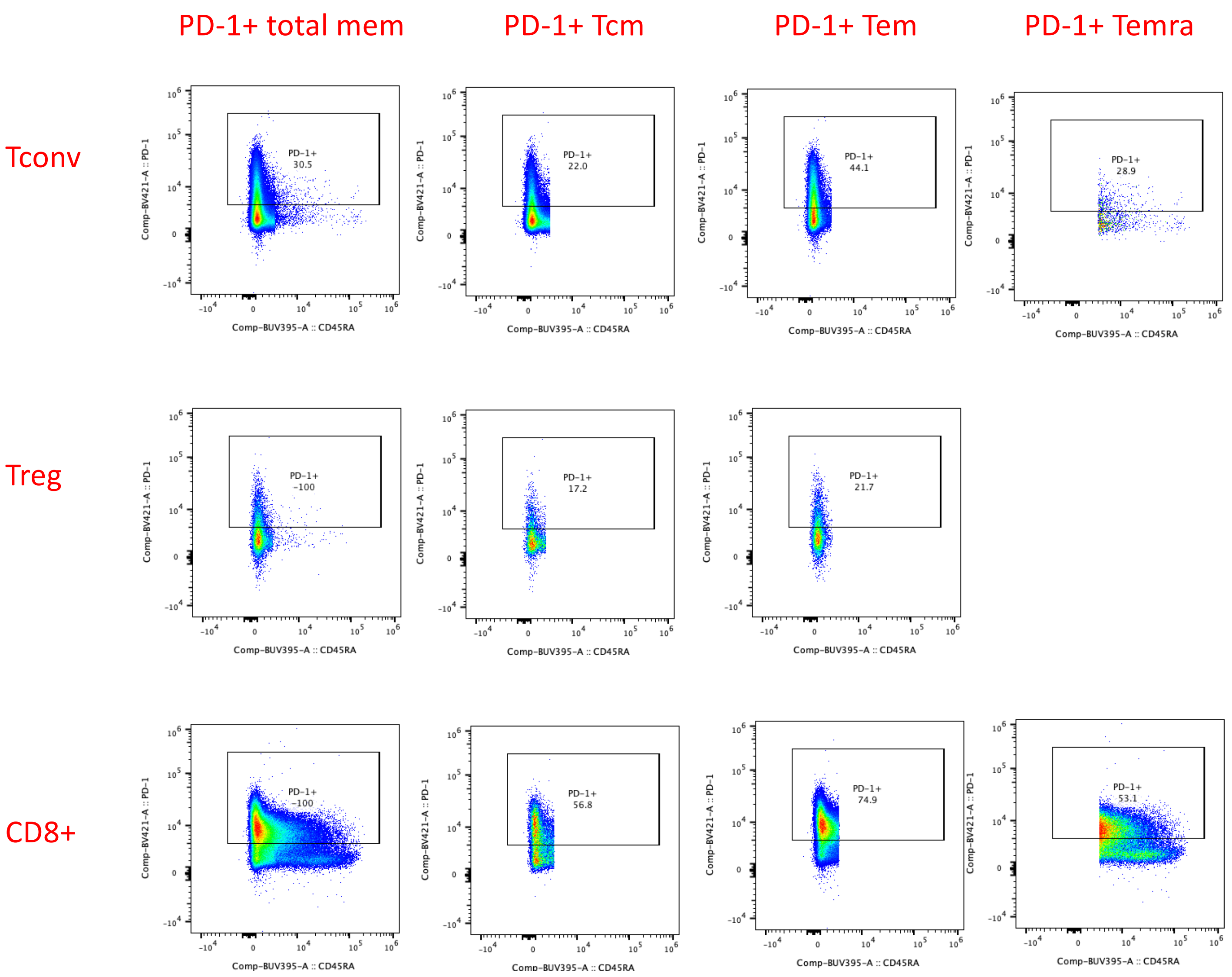
PD-1+ total mem
PD-1+ Tcm
PD-1+ Tem
PD-1+ Temra
Tconv
Treg
CD8+
PD-1+ 30.5
PD-1+ 22.0
PD-1+ 44.1
PD-1+ 28.9
PD-1+ ~100
PD-1+ 17.2
PD-1+ 21.7
PD-1+ ~100
PD-1+ 56.8
PD-1+ 74.9
PD-1+ 53.1
Comp-BV421-A :: PD-1
Comp-BUV395-A :: CD45RA

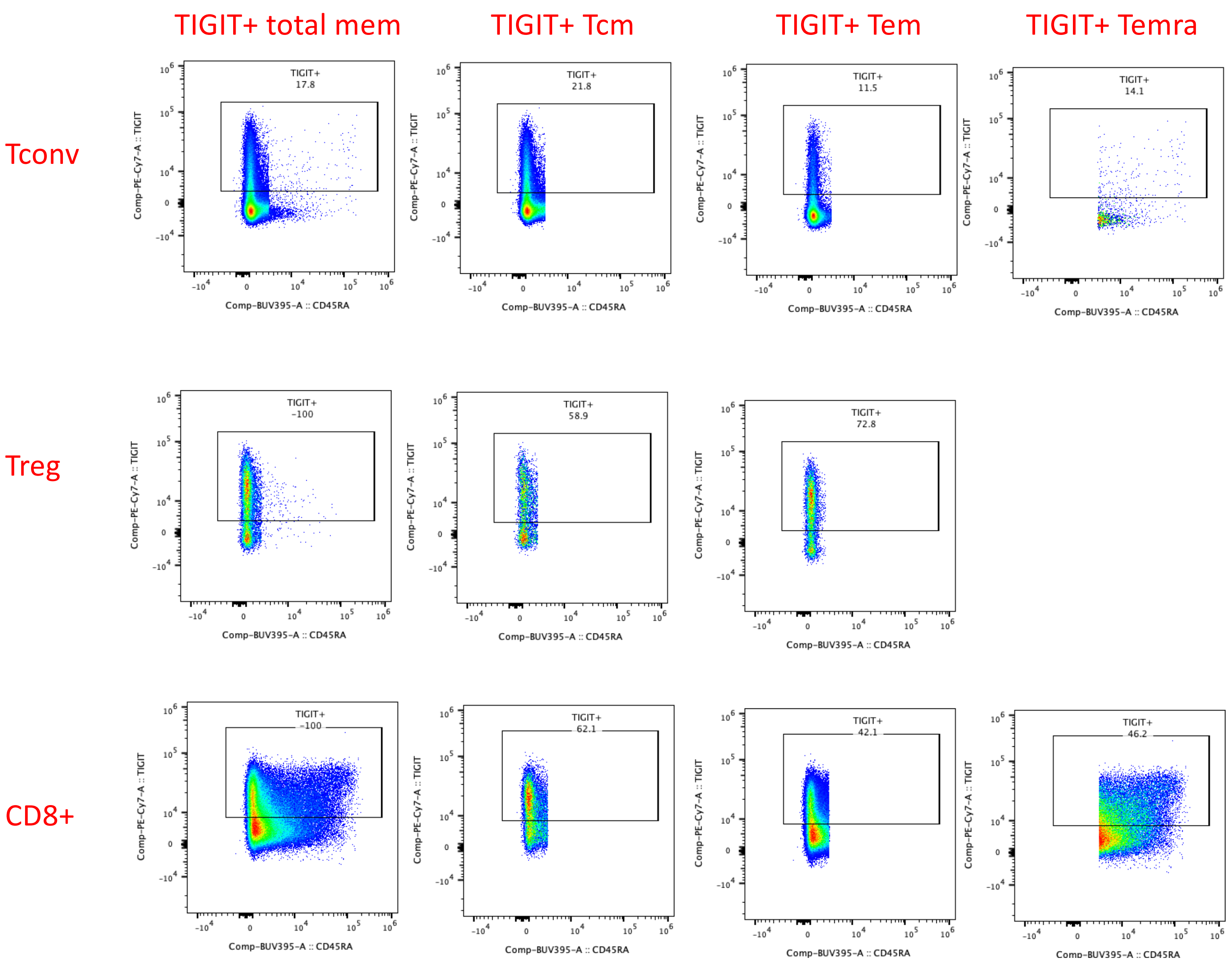
TIGIT+ total mem
TIGIT+ Tcm
TIGIT+ Tem
TIGIT+ Temra
Tconv
Treg
CD8+
TIGIT+
17.8
21.8
11.5
14.1
~100
58.9
72.8
~100
62.1
42.1
46.2
Comp-PE-Cy7-A :: TIGIT
Comp-BUV395-A :: CD45RA

KLRG1 & CD57 total mem | KLRG1 & TIGIT total mem | KLRG1 & TIGIT Tem | KLRG1 & TIGIT Temra

Tconv

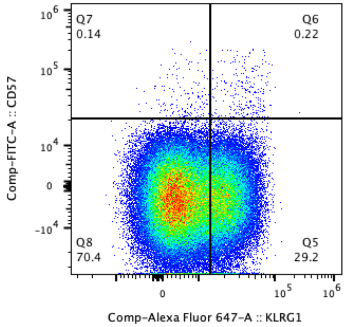


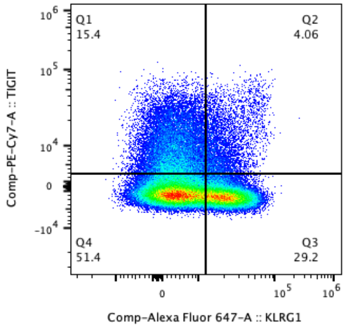


CD8+

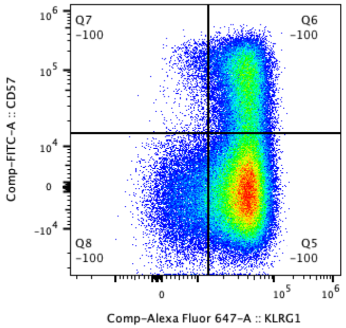


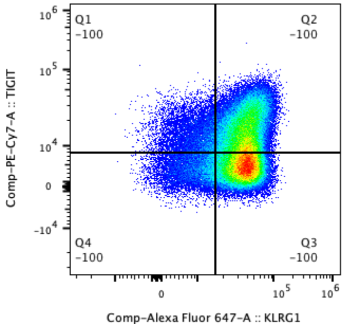


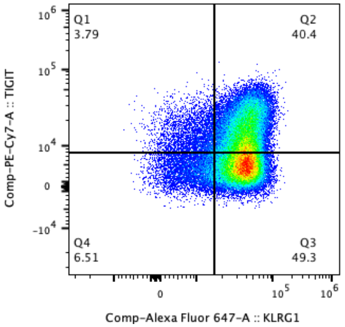


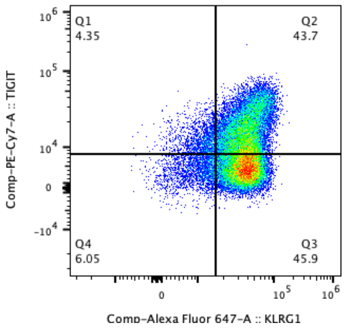

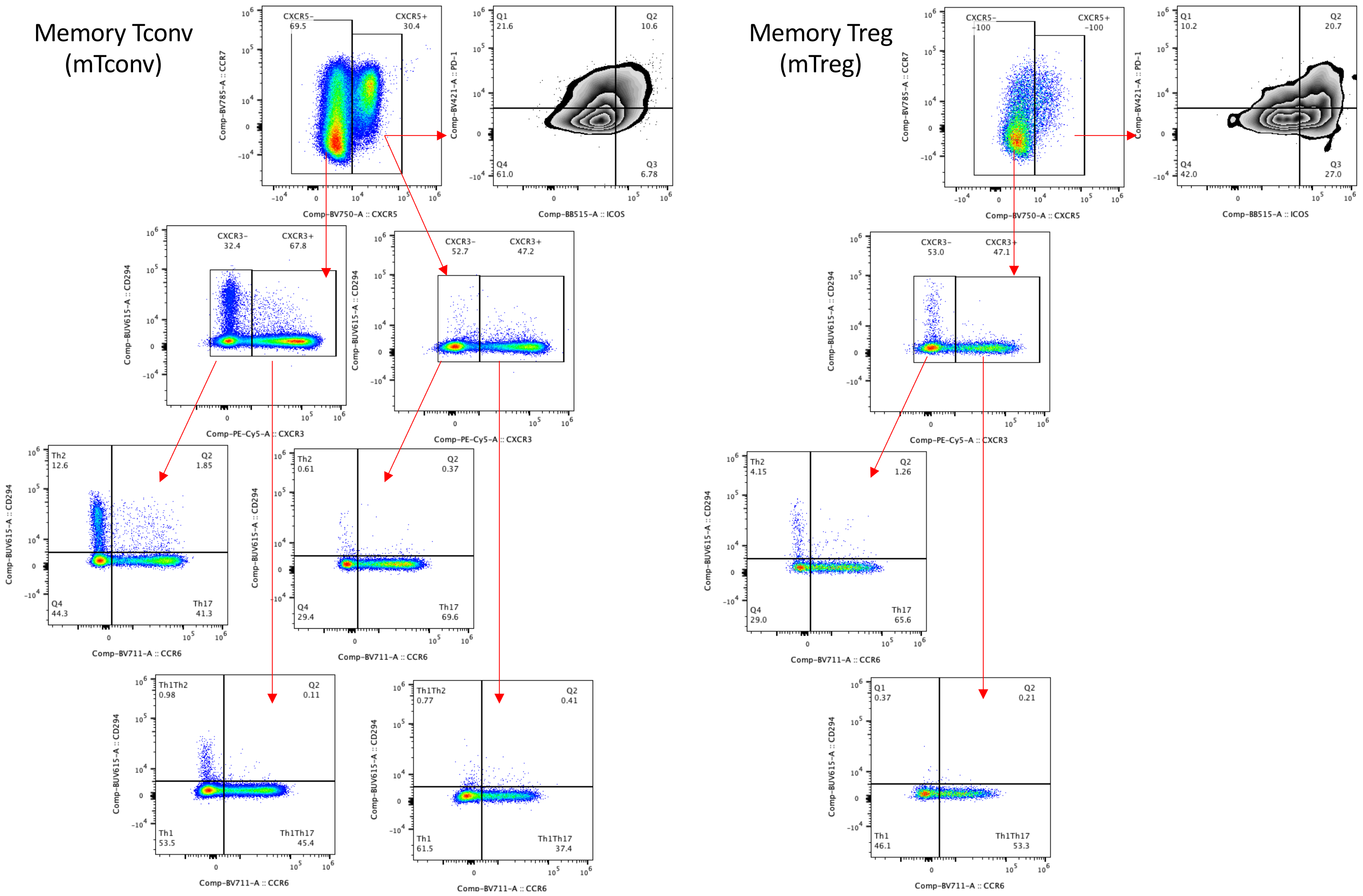
Memory Tconv (mTconv)
Memory Treg (mTreg)
CXCR5− 69.5
CXCR5+ 30.4
Comp-BV785-A :: CCR7
Comp-BV750-A :: CXCR5
Q1 21.6
Q2 10.6
Q3 6.78
Q4 61.0
Comp-BV421-A :: PD-1
Comp-BB515-A :: ICOS
CXCR3− 32.4
CXCR3+ 67.8
CXCR3− 52.7
CXCR3+ 47.2
Comp-BUV615-A :: CD294
Comp-PE-Cy5-A :: CXCR3
Th2 12.6
Q2 1.85
Q4 44.3
Th17 41.3
Th2 0.61
Q2 0.37
Q4 29.4
Th17 69.6
Comp-BV711-A :: CCR6
Th1Th2 0.98
Q2 0.11
Th1 53.5
Th1Th17 45.4
Th1Th2 0.77
Q2 0.41
Th1 61.5
Th1Th17 37.4
CXCR5− ~100
CXCR5+ ~100
Q1 10.2
Q2 20.7
Q3 27.0
Q4 42.0
CXCR3− 53.0
CXCR3+ 47.1
Th2 4.15
Q2 1.26
Q4 29.0
Th17 65.6
Q1 0.37
Q2 0.21
Th1 46.1
Th1Th17 53.3

### Non-CD3 slide

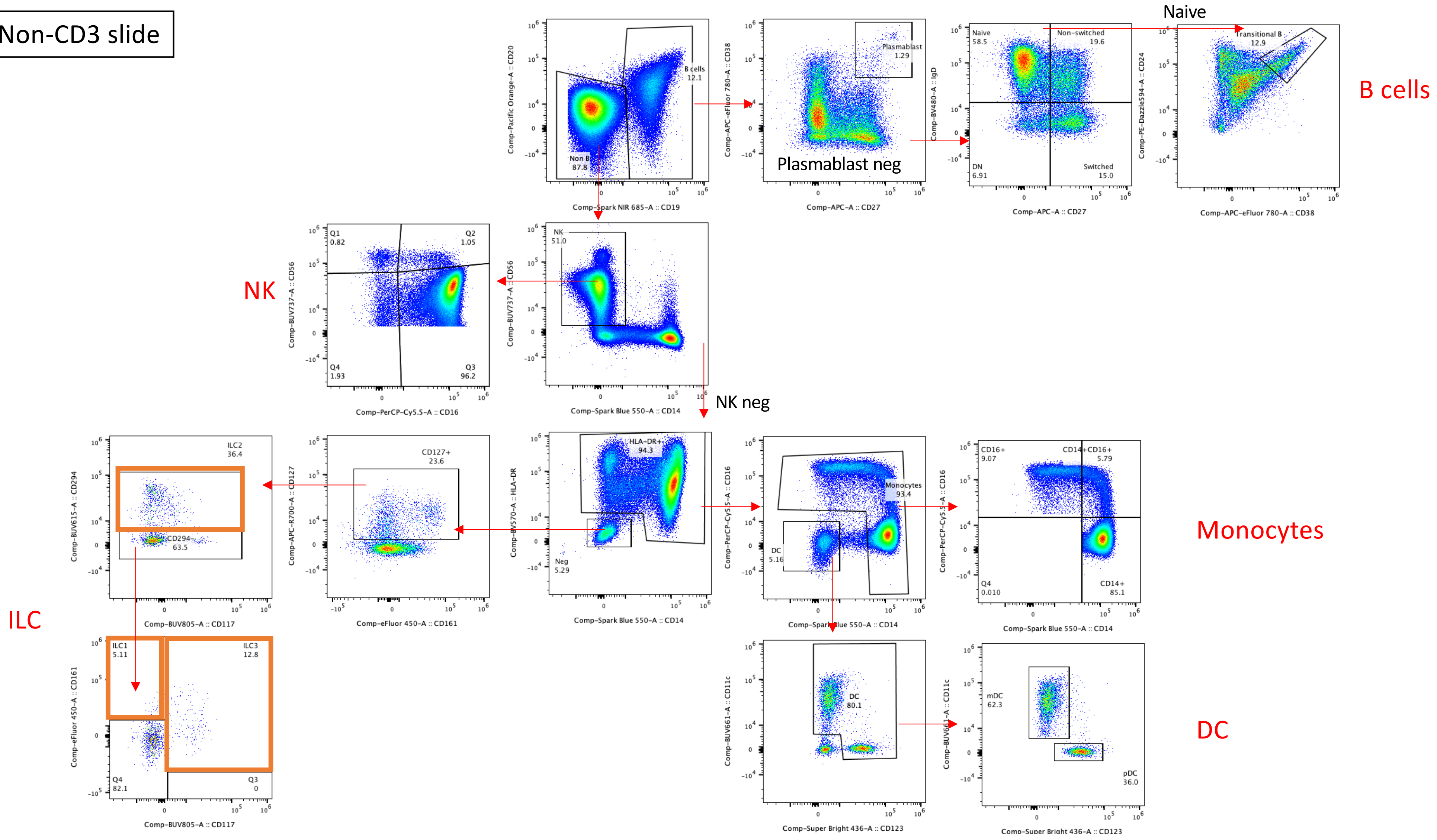
