## Supplementary Table 1 for "Multi-omics analysis reveals drivers of loss of β-cell function after newly diagnosed autoimmune type 1 diabetes: An INNODIA^‡^ multicenter study"

**Supplementary Table 1** - **Peptides and proteins measured using targeted proteomics.**

| Peptides | Protein Accession | Protein name | Gene |
|---|---|---|---|
| AGFAGDDAPR | C9JTX5 | Actin, cytoplasmic 1 (Fragment) | ACTB |
| AVFPSIVGR | C9JTX5 | Actin, cytoplasmic 1 (Fragment) | ACTB |
| LASPGFPGEYANDQER | O00187 | Mannan-binding lectin serine protease 2 | MASP2 |
| VLATLCGQESTDTER | O00187 | Mannan-binding lectin serine protease 2 | MASP2 |
| VTEPISAESGEQVER | O14791 | Apolipoprotein L1 | APOL1 |
| EATLQDCPSGPWGK | O43866 | CD5 antigen-like | CD5L |
| IWLDNVR | O43866 | CD5 antigen-like | CD5L |
| AFLLTPR | O95445 | Apolipoprotein M | APOM |
| SLTSCLDSK | O95445 | Apolipoprotein M | APOM |
| TLVVHEK | P00441 | Superoxide dismutase [Cu-Zn] | SOD1 |
| DDEEFIESNK | P00450 | Ceruloplasmin | CP |
| DIASGLIGPLIICK | P00450 | Ceruloplasmin | CP |
| TATSEYQTFFNPR | P00734 | Prothrombin | F2 |
| GLTLHLK | P00736 | Complement C1r subcomponent | C1R |
| HYEGSTVPEK | P00738 | Haptoglobin (Zonulin) | HP |
| VVLHPNYSQVDIGLIK | P00738 | Haptoglobin (Zonulin) | HP |
| NPANPVQ | H0Y300 | Haptoglobin (Zonulin) | HP |
| NPANPVQR | H0Y300 | Haptoglobin (Zonulin) | HP |
| FGSGYVSGWGR | P00740 | Coagulation factor IX | F9 |
| SALVLQYLR | P00740 | Coagulation factor IX | F9 |
| NGPLSCGQR | P00748 | Coagulation factor XII | F12 |
| AAVYHHFISDGVR | P01024 | Complement C3 | C3 |
| ENSQYQPIK | P01031 | Complement C5 | C5 |
| TDAPDLPEENQAR | P01031 | Complement C5 | C5 |
| QVVAGLNFR | P01042 | Kininogen-1 | KNG1 |
| TVGSDTFYSFK | P01042 | Kininogen-1 | KNG1 |
| GIVEECCFR | P01344 | Insulin-like growth factor II | IGF2 |
| FVYHLSDLCK | P01591 | Insulin-like growth factor II | IGJ |
| SSEDPNEDIVER | P01591 | Immunoglobulin J chain | IGJ |
| QVGSGVTTDQVQAEAK | P01871 | Immunoglobulin heavy constant mu | IGHM |
| YAATSQVLLPSK | P01871 | Immunoglobulin heavy constant mu | IGHM |
| ICVCDNGK | P02452 | Collagen alpha-1(I) chain | COL1A1 |
| VLCDDVICDETK | P02452 | Collagen alpha-1(I) chain | COL1A1 |
| DYVSQFEGSALGK | P02647 | Apolipoprotein A-I | APOA1 |
| THLAPYSDELR | P02647 | Apolipoprotein A-I | APOA1 |
| LEEQAQQIR | P02649 | Apolipoprotein E | APOE |
| LGPLVEQGR | P02649 | Apolipoprotein E | APOE |
| EFGNTLEDK | P02654 | Apolipoprotein C-I | APOC1 |
| EWFSETFQK | P02654 | Apolipoprotein C-I | APOC1 |
| TAAQNLYEK | P02655 | Apolipoprotein C-II | APOC2 |
| TYLPAVDEK | P02655 | Apolipoprotein C-II | APOC2 |

| | | | |
|---|---|---|---|
| DYWSTVK | P02656 | Apolipoprotein C-III | APOC3 |
| GWVTDGFSSLK | P02656 | Apolipoprotein C-III | APOC3 |
| ESSSHHPGIAEFPSR | P02671 | Fibrinogen alpha chain | FGA |
| GSESGIFTNTK | P02671 | Fibrinogen alpha chain | FGA |
| AFVFPK | P02741 | C-reactive protein | CRP |
| ESDTSYVSLK | P02741 | C-reactive protein | CRP |
| GYSIFSYATK | P02741 | C-reactive protein | CRP |
| ALPTTYEK | P02748 | Complement component C9 | C9 |
| FTPTETNK | P02748 | Complement component C9 | C9 |
| LSPIYNLVPVK | P02748 | Complement component C9 | C9 |
| AADDTWEPFASGK | P02766 | Transthyretin | TTR |
| TSESGELHGLTTEEEFVEGIYK | P02766 | Transthyretin | TTR |
| YLYEIAR | P02768 | Albumin | ALB |
| HLSLLTTLSNR | P02774 | Vitamin D-binding protein | GC |
| VLEPTLK | P02774 | Vitamin D-binding protein | GC |
| NIQSLEVIGK | P02775 | Platelet basic protein | PPBP |
| DGAGDVAFVK | P02787 | Serotransferrin | TF |
| EDPQTFYYAVAVVK | P02787 | Serotransferrin | TF |
| DSVTGTLPK | P03952 | Plasma kallikrein | KLKB1 |
| IAYGTQGSSGYSLR | P03952 | Plasma kallikrein | KLKB1 |
| VSEGNHDIALIK | P03952 | Plasma kallikrein | KLKB1 |
| EEIIYECDK | P04003 | C4b-binding protein alpha chain | C4BPA |
| GSSVIHCDADSK | P04003 | C4b-binding protein alpha chain | C4BPA |
| YTCLPGYVR | P04003 | C4b-binding protein alpha chain | C4BPA |
| ADVLTTGAGNPVGDK | P04040 | Catalase | CAT |
| FSTVAGESGSADTVR | P04040 | Catalase | CAT |
| ENFAGEATLQR | P04114 | Apolipoprotein B-100 | APOB |
| EVGTVLSQVYSK | P04114 | Apolipoprotein B-100 | APOB |
| ITENDIQIALDDAK | P04114 | Apolipoprotein B-100 | APOB |
| ADLFYDVEALDLESPK | P04196 | Histidine-rich glycoprotein | HRG |
| DGYLFQLLR | P04196 | Histidine-rich glycoprotein | HRG |
| ATWSGAVLAGR | P04217 | Alpha-1B-glycoprotein | A1BG |
| SGLSTGWTQLSK | P04217 | Alpha-1B-glycoprotein | A1BG |
| ILAGPAGDSNVVK | P04275 | von Willebrand factor | VWF |
| VTVFPIGIGDR | P04275 | von Willebrand factor | VWF |
| GALQNIIPASTGAAK | P04406 | Glyceraldehyde-3-phosphate dehydrogenase | GAPDH |
| LVINGNPITIFQER | P04406 | Glyceraldehyde-3-phosphate dehydrogenase | GAPDH |
| APQTGIVDECCFR | P05019 | Insulin-like growth factor I | IGF1 |
| GFYFNKPTGYGSSSR | P05019 | Insulin-like growth factor I | IGF1 |
| LLDSLPSDTR | P05155 | Plasma protease C1 inhibitor | SERPING1 |
| TLYSSSPR | P05155 | Plasma protease C1 inhibitor | SERPING1 |
| IANVFTNAFR | P05164 | Myeloperoxidase | MPO |
| NALALFVLPK | P05543 | Thyroxine-binding globulin | SERPINA7 |
| TLEAQLTPR | P05546 | Heparin cofactor 2 | SERPIND1 |
| AGALNSNDAFVLK | P06396 | Gelsolin | GSN |

| | | | |
|---|---|---|---|
| TPSAAYLWVGTGASEAEK | P06396 | Gelsolin | GSN |
| AVISPGFDVFAK | P06681 | Complement C2 | C2 |
| HAFILQDTK | P06681 | Complement C2 | C2 |
| ISASAEELR | P06727 | Apolipoprotein A-IV | APOA4 |
| LAPLAEDVR | P06727 | Apolipoprotein A-IV | APOA4 |
| LTQLNLDR | P07359 | Platelet glycoprotein Ib alpha chain | GP1BA |
| LTSLPLGALR | P07359 | Platelet glycoprotein Ib alpha chain | GP1BA |
| SLPVSDSVLSGFEQR | P07360 | Complement component C8 gamma chain | C8G |
| VQEAHLTEDQIFYFPK | P07360 | Complement component C8 gamma chain | C8G |
| STGGAPTFNVTVTK | P07737 | Profilin-1 | PFN1 |
| HLVALSPK | P08185 | Corticosteroid-binding globulin | SERPINA6 |
| ITQDAQLK | P08185 | Corticosteroid-binding globulin | SERPINA6 |
| AFQVWSDVTPLR | P08253 | 72 kDa type IV collagenase | MMP2 |
| GTYSTTVTGR | P08519 | Apolipoprotein | LPA |
| NPDAVAAPYCYTR | P08519 | Apolipoprotein | LPA |
| CTSTGWIPAPR | P08603 | Complement factor H | CFH |
| IDVHLVPDR | P08603 | Complement factor H | CFH |
| DFLQSLK | P08697 | Alpha-2-antiplasmin | SERPINF2 |
| LFGPDLK | P08697 | Alpha-2-antiplasmin | SERPINF2 |
| AQETSGEEISK | P08833 | Insulin-like growth factor-binding protein 1 | IGFBP1 |
| LHLDYIGPCK | P09486 | SPARC | SPARC |
| TFDSSCHFFATK | P09486 | SPARC | SPARC |
| EDTPNSVWEPAK | P09871 | Complement C1s subcomponent | C1S |
| FFGHGAEDSLADQAANEWGR | P0DJI8 | Serum amyloid A-1 protein | SAA1 |
| GPGGVWAAEAISDAR | P0DJI8 | Serum amyloid A-1 protein | SAA1 |
| GAEDSLADQAANK | P0DJI9 | Serum amyloid A-2 protein | SAA2 |
| GPGGAWAAEVISNAR | P0DJI9 | Serum amyloid A-2 protein | SAA2 |
| CNPDSNSANCLEEK | P10124 | Serglycin | SRGN |
| LRTDLFPK | P10124 | Serglycin | SRGN |
| ASSIIDELFQDR | P10909 | Clusterin | CLU |
| ITPSYVAFTPEGER | P11021 | Endoplasmic reticulum chaperone BiP | HSPA5 |
| NELESYAYSLK | P11021 | Endoplasmic reticulum chaperone BiP | HSPA5 |
| FQASVATPR | P11226 | Mannose-binding protein C | MBL2 |
| GFVVAGPSR | P13671 | Complement component C6 | C6 |
| IGESIELTCPK | P13671 | Complement component C6 | C6 |
| SLGPALLLLQK | P14780 | Matrix metalloproteinase-9 | MMP9 |
| SVDIWLR | P15151 | Poliovirus receptor | PVR |
| VLAKPQNTAEVQK | P15151 | Poliovirus receptor | PVR |
| FECQPGYR | P16109 | P-selectin | SELP |
| LEGPNNVECTTSGR | P16109 | P-selectin | SELP |
| LEGEVFFATR | P16112 | Aggrecan core protein | ACAN |
| ETEYGPCR | P17936 | Insulin-like growth factor-binding protein 3 | IGFBP3 |
| FLNVLSPR | P17936 | Insulin-like growth factor-binding protein 3 | IGFBP3 |
| SAGSVESPSVSSTHR | P17936 | Insulin-like growth factor-binding protein 3 | IGFBP3 |

| | | | |
|---|---|---|---|
| LEGEACGVYTPR | P18065 | Insulin-like growth factor-binding protein 2 | IGFBP2 |
| LIQGAPTIR | P18065 | Insulin-like growth factor-binding protein 2 | IGFBP2 |
| GSFALSFPVESDVAPIAR | P20742 | Pregnancy zone protein | PZP |
| ISEITNIVSK | P20742 | Pregnancy zone protein | PZP |
| AGQSAAGAAPGGGVDTR | P21333 | Filamin-A | FLNA |
| VEPGLGADNSVVR | P21333 | Filamin-A | FLNA |
| WGDEHIPGSPYR | P21333 | Filamin-A | FLNA |
| FLVGPDGIPIMR | P22352 | Glutathione peroxidase 3 | GPX3 |
| AGEVQEPELR | P25311 | Zinc-alpha-2-glycoprotein | AZGP1 |
| EIPAWVPFDPAAQITK | P25311 | Zinc-alpha-2-glycoprotein | AZGP1 |
| YSLTYIYTGLSK | P25311 | Zinc-alpha-2-glycoprotein | AZGP1 |
| SPLNDFQVLR | P26927 | Hepatocyte growth factor-like protein | MST1 |
| TPFDYCALR | P26927 | Hepatocyte growth factor-like protein | MST1 |
| GPGGVWAAK | P35542 | Serum amyloid A-4 protein | SAA4 |
| YLYAR | P35542 | Serum amyloid A-4 protein | SAA4 |
| LAELPADALGPLQR | P35858 | Insulin-like growth factor-binding protein complex acid labile subunit | IGFALS |
| LEALPNSLLAPLGR | P35858 | Insulin-like growth factor-binding protein complex acid labile subunit | IGFALS |
| LEYLLLSR | P35858 | Insulin-like growth factor-binding protein complex acid labile subunit | IGFALS |
| LSSGLVTAALYGR | P43251 | Biotinidase | BTD |
| WNPCLEPHR | P43251 | Biotinidase | BTD |
| AESPEVCFNEESPK | P43652 | Afamin | AFM |
| DADPDTFFAK | P43652 | Afamin | AFM |
| GQCIINSNK | P43652 | Afamin | AFM |
| FNALQYLR | P51884 | Lumican | LUM |
| ILGPLSYSK | P51884 | Lumican | LUM |
| AVSPPAR | P54108 | Cysteine-rich secretory protein 3 | CRISP3 |
| YEDLYSNCK | P54108 | Cysteine-rich secretory protein 3 | CRISP3 |
| AWFLESK | P55056 | Apolipoprotein C-IV | APOC4 |
| ELLETVVNR | P55056 | Apolipoprotein C-IV | APOC4 |
| WSLVR | P55056 | Apolipoprotein C-IV | APOC4 |
| IPACIAGER | P59666 | Neutrophil defensin 3 | DEFA3 |
| YGTCIYQGR | P59666 | Neutrophil defensin 3 | DEFA3 |
| SAVTALWGK | P68871 | Hemoglobin subunit beta | HBB |
| VNVDEVGGEALGR | P68871 | Hemoglobin subunit beta | HBB |
| ATYIQNYR | Q01459 | Di-N-acetylchitobiase | CTBS |
| STDTSCVNPPTVQNAHILSR | Q03591 | Complement factor H-related protein 1 | CFHR1 |
| TGESAEFVCK | Q03591 | Complement factor H-related protein 1 | CFHR1 |
| LEACESLTR | Q04756 | Hepatocyte growth factor activator | HGFAC |
| VANYVDWINDR | Q04756 | Hepatocyte growth factor activator | HGFAC |
| LTLLAPLNSVFK | Q15582 | Transforming growth factor-beta-induced protein ig-h3 | TGFBI |
| GFPGIQGR | Q15848 | Adiponectin | ADIPOQ |
| IFYNQQNHYDGSTGK | Q15848 | Adiponectin | ADIPOQ |

| | | | |
|---|---|---|---|
| FLPEGCQPLVSSAVDR | Q86YW5 | Trem-like transcript 1 protein | TREML1 |
| VSLNILPPEEEEETHK | Q86YW5 | Trem-like transcript 1 protein | TREML1 |
| TLIDFVK | Q8WXD2 | Secretogranin-3 | SCG3 |
| VEYQCQSYYELQGSK | Q92496 | Complement factor H-related protein 4 | CFHR4 |
| VYLPWSR | Q92496 | Complement factor H-related protein 4 | CFHR4 |
| AIHLDLEEYR | Q96KN2 | Beta-Ala-His dipeptidase | CNDP1 |
| ALEQDLPVNIK | Q96KN2 | Beta-Ala-His dipeptidase | CNDP1 |
| AGLLRPDYALLGHR | Q96PD5 | N-acetylmuramoyl-L-alanine amidase | PGLYRP2 |
| TFTLLDPK | Q96PD5 | N-acetylmuramoyl-L-alanine amidase | PGLYRP2 |
| IFSQETLTK | Q9UHG3 | Prenylcysteine oxidase 1 | PCYOX1 |
| SDFYDIVLVATPLNR | Q9UHG3 | Prenylcysteine oxidase 1 | PCYOX1 |
| LLGLSLAGK | Q9Y5Y7 | Lymphatic vessel endothelial hyaluronic acid receptor 1 | LYVE1 |
| LGGNETQVR | MSRT1 | Isotopically labelled retention time std. | |
| ADEGISFR | MSRT1 | Isotopically labelled retention time std. | |
| AEFAEVSK | MSRT1 | Isotopically labelled retention time std. | |
| AVQQPDGLAVLGIFLK | MSRT1 | Isotopically labelled retention time std. | |
| DISLSDYK | MSRT1 | Isotopically labelled retention time std. | |
| DQGGELLSLR | MSRT1 | Isotopically labelled retention time std. | |
| GLFIIDDK | MSRT1 | Isotopically labelled retention time std. | |
| LVNEVTEFAK | MSRT1 | Isotopically labelled retention time std. | |
| SGFSSVSVSR | MSRT1 | Isotopically labelled retention time std. | |
| TDELFQIEGLKEELAYLR | MSRT1 | Isotopically labelled retention time std. | |
| YWGVASFLQK | MSRT1 | Isotopically labelled retention time std. | |
