## Supplementary Table 2 for "Multi-omics analysis reveals drivers of loss of β-cell function after newly diagnosed autoimmune type 1 diabetes: An INNODIA^‡^ multicenter study"

### Gene List

#### HTG EdgeSeq
#### miRNA Whole Transcriptome Assay

##### Rat

| | | | | | | | | | | | |
|---|---|---|---|---|---|---|---|---|---|---|---|
| let-7a-2-3p | miR-122-5p | miR-132-3p | miR-145-5p | miR-181d-5p | miR-199a-5p | miR-222-3p | miR-296-3p | miR-30d-3p | miR-369-3p | miR-484 | miR-873-5p |
| let-7a-5p | miR-124-3p | miR-132-5p | miR-146a-5p | miR-183-5p | miR-19a-3p | miR-22-3p | miR-296-5p | miR-30d-5p | miR-369-5p | miR-485-5p | miR-874-3p |
| let-7b-5p | miR-124-5p | miR-133a-3p | miR-148b-3p | miR-184 | miR-19b-3p | miR-22-5p | miR-29a-3p | miR-30e-3p | miR-370-3p | miR-487b-3p | miR-92b-3p |
| let-7c-5p | miR-1249 | miR-133a-5p | miR-150-5p | miR-185-5p | miR-200a-3p | miR-23a-3p | miR-29a-5p | miR-30e-5p | miR-377-5p | miR-488-5p | miR-935 |
| let-7d-3p | miR-125a-3p | miR-134-5p | miR-152-3p | miR-186-5p | miR-200b-5p | miR-23a-5p | miR-29b-2-5p | miR-324-5p | miR-379-5p | miR-490-5p | miR-93-5p |
| let-7d-5p | miR-125a-5p | miR-135a-5p | miR-153-3p | miR-187-3p | miR-204-5p | miR-23b-3p | miR-29b-3p | miR-32-5p | miR-381-5p | miR-493-5p | miR-96-5p |
| let-7e-3p | miR-125b-1-3p | miR-135b-3p | miR-154-3p | miR-188-5p | miR-208a-5p | miR-24-3p | miR-29c-3p | miR-330-5p | miR-382-3p | miR-532-3p | miR-99a-5p |
| let-7e-5p | miR-125b-5p | miR-135b-5p | miR-154-5p | miR-18a-5p | miR-20a-5p | miR-25-3p | miR-29c-5p | miR-331-3p | miR-382-5p | miR-539-5p | miR-99b-3p |
| let-7f-5p | miR-127-3p | miR-136-3p | miR-15b-3p | miR-190a-5p | miR-20b-5p | miR-26a-5p | miR-301a-3p | miR-339-5p | miR-410-3p | miR-542-3p | miR-99b-5p |
| let-7i-5p | miR-128-3p | miR-136-5p | miR-15b-5p | miR-192-5p | miR-210-3p | miR-26b-3p | miR-30a-3p | miR-340-5p | miR-410-5p | miR-568 | |
| miR-100-5p | miR-129-2-3p | miR-138-5p | miR-16-5p | miR-194-5p | miR-21-5p | miR-26b-5p | miR-30a-5p | miR-342-3p | miR-411-5p | miR-652-3p | |
| miR-106b-3p | miR-129-5p | miR-140-5p | miR-17-5p | miR-195-5p | miR-216a-5p | miR-27a-3p | miR-30b-5p | miR-34a-5p | miR-425-5p | miR-708-5p | |
| miR-106b-5p | miR-1306-5p | miR-141-3p | miR-181a-5p | miR-196a-5p | miR-216b-5p | miR-27a-5p | miR-30c-1-3p | miR-34c-5p | miR-433-3p | miR-758-3p | |
| miR-10a-3p | miR-130a-3p | miR-142-5p | miR-181b-5p | miR-196b-5p | miR-219a-2-3p | miR-27b-3p | miR-30c-2-3p | miR-361-5p | miR-433-5p | miR-759 | |
| miR-10a-5p | miR-130b-3p | miR-144-3p | miR-181c-5p | miR-199a-3p | miR-221-3p | miR-28-5p | miR-30c-5p | miR-363-5p | miR-455-5p | miR-761 | |

##### Mouse

| | | | | | | | | | | | |
|---|---|---|---|---|---|---|---|---|---|---|---|
| let-7a-5p | miR-1247-5p | miR-135b-5p | miR-16-5p | miR-194-5p | miR-20a-5p | miR-25-3p | miR-302d-3p | miR-339-5p | miR-409-5p | miR-495-3p | miR-759 |
| let-7b-5p | miR-1251-3p | miR-138-5p | miR-17-5p | miR-196a-5p | miR-20b-5p | miR-26a-1-3p | miR-30a-3p | miR-340-3p | miR-410-3p | miR-504-5p | miR-761 |
| let-7c-5p | miR-125a-3p | miR-140-5p | miR-181a-5p | miR-196b-3p | miR-210-3p | miR-26a-5p | miR-30a-5p | miR-340-5p | miR-410-5p | miR-532-3p | miR-874-3p |
| let-7d-3p | miR-125a-5p | miR-141-3p | miR-181b-5p | miR-196b-5p | miR-212-5p | miR-26b-3p | miR-30b-5p | miR-342-3p | miR-411-3p | miR-532-5p | miR-875-5p |
| let-7d-5p | miR-125b-1-3p | miR-142-5p | miR-181c-5p | miR-199a-5p | miR-214-5p | miR-26b-5p | miR-30c-1-3p | miR-34a-5p | miR-411-5p | miR-539-5p | miR-877-5p |
| let-7e-3p | miR-125b-5p | miR-143-3p | miR-181d-5p | miR-19a-3p | miR-216a-5p | miR-27a-3p | miR-30c-2-3p | miR-34c-5p | miR-425-5p | miR-542-3p | miR-92b-3p |
| let-7e-5p | miR-127-3p | miR-144-3p | miR-183-3p | miR-19b-1-5p | miR-216b-5p | miR-27a-5p | miR-30c-5p | miR-361-5p | miR-431-3p | miR-551b-3p | miR-93-3p |
| let-7f-1-3p | miR-127-5p | miR-146a-5p | miR-183-5p | miR-19b-3p | miR-218-2-3p | miR-27b-3p | miR-30d-3p | miR-363-3p | miR-431-5p | miR-568 | miR-93-5p |
| let-7f-5p | miR-128-3p | miR-146b-5p | miR-185-5p | miR-200a-3p | miR-218-5p | miR-27b-5p | miR-30d-5p | miR-367-3p | miR-433-3p | miR-574-3p | miR-9-3p |
| let-7g-5p | miR-129-1-3p | miR-148a-3p | miR-186-5p | miR-200a-5p | miR-219a-2-3p | miR-296-3p | miR-30e-3p | miR-369-3p | miR-433-5p | miR-574-5p | miR-9-5p |
| let-7i-5p | miR-129-2-3p | miR-148b-3p | miR-187-3p | miR-200b-3p | miR-219b-3p | miR-296-5p | miR-30e-5p | miR-369-5p | miR-450a-5p | miR-590-3p | miR-96-5p |
| miR-100-5p | miR-129-5p | miR-149-5p | miR-188-3p | miR-200b-5p | miR-221-3p | miR-29a-3p | miR-324-5p | miR-370-3p | miR-451a | miR-590-5p | miR-99a-5p |
| miR-106b-3p | miR-1298-5p | miR-150-5p | miR-188-5p | miR-200c-3p | miR-222-3p | miR-29a-5p | miR-32-5p | miR-374b-5p | miR-455-5p | miR-615-3p | miR-99b-3p |
| miR-106b-5p | miR-130a-3p | miR-152-3p | miR-18a-5p | miR-200c-5p | miR-223-3p | miR-29b-3p | miR-328-3p | miR-377-3p | miR-484 | miR-652-3p | miR-99b-5p |
| miR-10a-3p | miR-130b-3p | miR-153-3p | miR-190a-5p | miR-204-3p | miR-22-3p | miR-29c-3p | miR-330-5p | miR-377-5p | miR-485-5p | miR-654-3p | |
| miR-10a-5p | miR-132-3p | miR-154-3p | miR-191-5p | miR-204-5p | miR-22-5p | miR-29c-5p | miR-331-3p | miR-379-3p | miR-487b-3p | miR-671-3p | |
| miR-10b-5p | miR-133a-3p | miR-154-5p | miR-192-3p | miR-205-5p | miR-23a-3p | miR-301a-3p | miR-331-5p | miR-379-5p | miR-490-5p | miR-671-5p | |
| miR-122-5p | miR-134-5p | miR-15a-5p | miR-192-5p | miR-208a-3p | miR-23a-5p | miR-301a-5p | miR-335-3p | miR-381-3p | miR-491-3p | miR-708-5p | |
| miR-124-3p | miR-135a-5p | miR-15b-3p | miR-193a-3p | miR-208a-5p | miR-23b-3p | miR-302a-3p | miR-335-5p | miR-382-5p | miR-491-5p | miR-744-3p | |
| miR-124-5p | miR-135b-3p | miR-15b-5p | miR-193b-5p | miR-208b-3p | miR-24-3p | miR-302b-3p | miR-338-3p | miR-409-3p | miR-494-3p | miR-744-5p | |

##### Human

| | | | | | | | | | | | |
|---|---|---|---|---|---|---|---|---|---|---|---|
| let-7a-2-3p | miR-105-3p | miR-1197 | miR-1236-5p | miR-1257 | miR-1273e | miR-129-2-3p | miR-130a-3p | miR-138-1-3p | miR-147b | miR-15b-5p | miR-187-3p |
| let-7a-3p | miR-105-5p | miR-1199-3p | miR-1237-3p | miR-1258 | miR-1273g-5p | miR-1292-5p | miR-130a-5p | miR-138-2-3p | miR-148a-3p | miR-16-1-3p | miR-187-5p |
| let-7a-5p | miR-106a-3p | miR-1200 | miR-1237-5p | miR-125a-3p | miR-1273h-5p | miR-1293 | miR-130b-3p | miR-138-5p | miR-148b-3p | miR-16-5p | miR-188-3p |
| let-7b-5p | miR-106a-5p | miR-1202 | miR-1238-3p | miR-125a-5p | miR-127-3p | miR-1294 | miR-130b-5p | miR-139-3p | miR-149-3p | miR-17-3p | miR-188-5p |
| let-7c-3p | miR-106b-3p | miR-1204 | miR-1238-5p | miR-125b-1-3p | miR-1275 | miR-1295b-3p | miR-1321 | miR-139-5p | miR-149-5p | miR-17-5p | miR-18a-3p |
| let-7c-5p | miR-106b-5p | miR-1205 | miR-1243 | miR-125b-2-3p | miR-127-5p | miR-129-5p | miR-1322 | miR-140-5p | miR-150-3p | miR-181a-2-3p | miR-18a-5p |
| let-7d-3p | miR-107 | miR-1206 | miR-124-3p | miR-125b-5p | miR-1276 | miR-1296-3p | miR-1323 | miR-141-3p | miR-150-5p | miR-181a-3p | miR-18b-3p |
| let-7d-5p | miR-10a-3p | miR-1207-3p | miR-1244 | miR-1261 | miR-1277-5p | miR-1296-5p | miR-132-3p | miR-141-5p | miR-151a-3p | miR-181a-5p | miR-18b-5p |
| let-7e-3p | miR-10a-5p | miR-1207-5p | miR-1245b-3p | miR-1262 | miR-1278 | miR-1297 | miR-1324 | miR-142-5p | miR-152-3p | miR-181b-3p | miR-1908-5p |
| let-7e-5p | miR-10b-3p | miR-1208 | miR-124-5p | miR-1263 | miR-1279 | miR-1298-3p | miR-132-5p | miR-143-3p | miR-153-3p | miR-181b-5p | miR-1909-3p |
| let-7f-1-3p | miR-10b-5p | miR-1224-5p | miR-1247-3p | miR-126-3p | miR-128-1-5p | miR-1298-5p | miR-133a-3p | miR-143-5p | miR-153-5p | miR-181c-3p | miR-1909-5p |
| let-7f-2-3p | miR-1178-3p | miR-1225-3p | miR-1247-5p | miR-1264 | miR-1282 | miR-1299 | miR-133a-5p | miR-144-3p | miR-1537-3p | miR-181c-5p | miR-190a-3p |
| let-7f-5p | miR-1178-5p | miR-1225-5p | miR-1249 | miR-1265 | miR-128-2-5p | miR-1301-3p | miR-133b | miR-144-5p | miR-1537-5p | miR-181d-3p | miR-190a-5p |
| let-7g-3p | miR-1179 | miR-122-5p | miR-1250-3p | miR-126-5p | miR-1283 | miR-1301-5p | miR-1343-5p | miR-145-3p | miR-1538 | miR-181d-5p | miR-190b |
| let-7g-5p | miR-1180-3p | miR-1226-3p | miR-1250-5p | miR-1267 | miR-128-3p | miR-1302 | miR-134-3p | miR-145-5p | miR-1539 | miR-182-3p | miR-1910-3p |
| let-7i-5p | miR-1180-5p | miR-1227-3p | miR-1251-3p | miR-1269a | miR-1285-5p | miR-1303 | miR-134-5p | miR-1468-3p | miR-154-3p | miR-182-5p | miR-1910-5p |
| miR-1 | miR-1182 | miR-1228-3p | miR-1252-3p | miR-1269b | miR-1287-3p | miR-1304-3p | miR-135a-3p | miR-1468-5p | miR-154-5p | miR-183-3p | miR-1911-3p |
| miR-100-3p | miR-1183 | miR-1229-3p | miR-1252-5p | miR-1270 | miR-1287-5p | miR-1304-5p | miR-135a-5p | miR-146a-3p | miR-155-3p | miR-183-5p | miR-1911-5p |
| miR-100-5p | miR-1184 | miR-1229-5p | miR-1253 | miR-1271-5p | miR-1288-3p | miR-1305 | miR-135b-3p | miR-146a-5p | miR-155-5p | miR-184 | miR-1912 |
| miR-101-3p | miR-1185-1-3p | miR-1231 | miR-1254 | miR-1272 | miR-1289 | miR-1306-3p | miR-135b-5p | miR-146b-3p | miR-1587 | miR-185-3p | miR-1913 |
| miR-101-5p | miR-1185-2-3p | miR-1233-3p | miR-1255b-2-3p | miR-1273a | miR-1290 | miR-1306-5p | miR-136-3p | miR-146b-5p | miR-15a-3p | miR-185-5p | miR-191-3p |
| miR-103a-2-5p | miR-1185-5p | miR-1234-3p | | miR-1273c | miR-1291 | miR-1307-3p | miR-136-5p | miR-1471 | miR-15a-5p | miR-186-3p | miR-1914-3p |
| miR-103a-3p | miR-1193 | miR-1236-3p | miR-1256 | miR-1273d | miR-129-1-3p | miR-1307-5p | miR-137 | miR-147a | miR-15b-3p | miR-186-5p | miR-1914-5p |

### Gene List

#### HTG EdgeSeq
#### miRNA Whole Transcriptome Assay

##### Human Continued

| | | | | | | | | | | | |
|---|---|---|---|---|---|---|---|---|---|---|---|
| miR-1915-3p | miR-2115-3p | miR-27b-3p | miR-3130-3p | miR-3189-5p | miR-3607-3p | miR-3682-5p | miR-383-5p | miR-4255 | miR-432-3p | miR-4471 | miR-4524b-5p |
| miR-1915-5p | miR-2115-5p | miR-27b-5p | miR-3131 | miR-3191-3p | miR-3607-5p | miR-3683 | miR-384 | miR-425-5p | miR-4324 | miR-4472 | miR-4525 |
| miR-191-5p | miR-211-5p | miR-28-5p | miR-3132 | miR-3191-5p | miR-3609 | miR-3684 | miR-3907 | miR-4256 | miR-4325 | miR-4473 | miR-452-5p |
| miR-192-3p | miR-2116-3p | miR-2861 | miR-3133 | miR-3192-3p | miR-3611 | miR-3685 | miR-3908 | miR-4257 | miR-432-5p | miR-4474-3p | miR-4527 |
| miR-192-5p | miR-2116-5p | miR-2909 | miR-3134 | miR-3192-5p | miR-3612 | miR-3686 | miR-3909 | miR-4259 | miR-4326 | miR-4475 | miR-4528 |
| miR-193a-3p | miR-2117 | miR-296-3p | miR-3135a | miR-3193 | miR-3613-3p | miR-3687 | miR-3910 | miR-4261 | miR-4327 | miR-4476 | miR-4530 |
| miR-193a-5p | miR-212-3p | miR-296-5p | miR-3136-5p | miR-3197 | miR-3613-5p | miR-3688-5p | miR-3912-3p | miR-4262 | miR-4328 | miR-4477a | miR-4531 |
| miR-193b-3p | miR-212-5p | miR-297 | miR-3137 | miR-3198 | miR-361-3p | miR-3689a-3p | miR-3912-5p | miR-4263 | miR-4329 | miR-4477b | miR-4533 |
| miR-193b-5p | miR-21-3p | miR-298 | miR-3138 | miR-3199 | miR-3614-3p | miR-3689a-5p | miR-3913-5p | miR-4264 | miR-4330 | miR-4478 | miR-4534 |
| miR-194-3p | miR-214-5p | miR-299-3p | miR-3140-3p | miR-3200-5p | miR-3614-5p | miR-3689b-3p | miR-3914 | miR-4265 | miR-433-3p | miR-448 | miR-4535 |
| miR-194-5p | miR-215-3p | miR-299-5p | miR-3140-5p | miR-3201 | miR-361-5p | miR-3689d | miR-3915 | miR-4266 | miR-433-5p | miR-4480 | miR-4536-5p |
| miR-195-3p | miR-215-5p | miR-29a-3p | miR-3141 | miR-3202 | miR-3616-3p | miR-3689f | miR-3916 | miR-4267 | miR-4417 | miR-4481 | miR-4537 |
| miR-195-5p | miR-21-5p | miR-29a-5p | miR-3142 | miR-320a | miR-3616-5p | miR-3690 | miR-3917 | miR-4268 | miR-4419a | miR-4482-3p | miR-4538 |
| miR-196a-3p | miR-216a-3p | miR-29b-1-5p | miR-3143 | miR-320b | miR-3617-3p | miR-3691-3p | miR-3919 | miR-4269 | miR-4419b | miR-4482-5p | miR-4539 |
| miR-196a-5p | miR-216a-5p | miR-29b-2-5p | miR-3144-3p | miR-320c | miR-3617-5p | miR-3691-5p | miR-3920 | miR-4270 | miR-4420 | miR-4483 | miR-4540 |
| miR-196b-3p | miR-216b-3p | miR-29b-3p | miR-3144-5p | miR-320d | miR-3618 | miR-3692-3p | miR-3921 | miR-4271 | miR-4421 | miR-4484 | miR-454-3p |
| miR-196b-5p | miR-216b-5p | miR-29c-3p | miR-3145-3p | miR-320e | miR-3619-3p | miR-3692-5p | miR-3922-3p | miR-4272 | miR-4422 | miR-4486 | miR-454-5p |
| miR-197-3p | miR-217 | miR-29c-5p | miR-3145-5p | miR-323a-3p | miR-3619-5p | miR-369-3p | miR-3922-5p | miR-4273 | miR-4423-3p | miR-4487 | miR-455-3p |
| miR-197-5p | miR-218-1-3p | miR-300 | miR-3146 | miR-323a-5p | miR-3620-3p | miR-369-5p | miR-3923 | miR-4274 | miR-4423-5p | miR-4489 | miR-455-5p |
| miR-1976 | miR-218-2-3p | miR-301a-3p | miR-3147 | miR-323b-3p | miR-362-3p | miR-370-3p | miR-3924 | miR-4275 | miR-4424 | miR-4490 | miR-4632-3p |
| miR-198 | miR-218-5p | miR-301a-5p | miR-3148 | miR-323b-5p | miR-362-5p | miR-370-5p | miR-3925-5p | miR-4276 | miR-4425 | miR-4491 | miR-4632-5p |
| miR-199a-3p | miR-219a-1-3p | miR-301b | miR-3149 | miR-32-3p | miR-363-3p | miR-3713 | miR-3926 | miR-4277 | miR-4426 | miR-4493 | miR-4636 |
| miR-199a-5p | miR-219a-2-3p | miR-302a-3p | miR-3150b-5p | miR-324-3p | miR-363-5p | miR-3714 | miR-3927-5p | miR-4278 | miR-4427 | miR-4494 | miR-4637 |
| miR-199b-5p | miR-219b-3p | miR-302a-5p | miR-3152-3p | miR-324-5p | miR-3646 | miR-371b-3p | miR-3928-3p | miR-4279 | miR-4428 | miR-4495 | miR-4638-5p |
| miR-19a-3p | miR-221-3p | miR-302b-3p | miR-3152-5p | miR-325 | miR-3648 | miR-371b-5p | miR-3928-5p | miR-4280 | miR-4429 | miR-4496 | miR-4639-3p |
| miR-19a-5p | miR-221-5p | miR-302b-5p | miR-3153 | miR-32-5p | miR-3649 | miR-372-3p | miR-3929 | miR-4282 | miR-4430 | miR-4498 | miR-4639-5p |
| miR-19b-1-5p | miR-222-3p | miR-302c-3p | miR-3154 | miR-326 | miR-3650 | miR-372-5p | miR-3934-3p | miR-4283 | miR-4431 | miR-4499 | miR-4640-3p |
| miR-19b-2-5p | miR-222-5p | miR-302c-5p | miR-3155a | miR-328-3p | miR-3652 | miR-373-3p | miR-3934-5p | miR-4287 | miR-4432 | miR-449a | miR-4641 |
| miR-19b-3p | miR-223-3p | miR-302d-3p | miR-3155b | miR-329-3p | miR-3653 | miR-373-5p | miR-3935 | miR-4288 | miR-4433b-5p | miR-449b-3p | miR-4642 |
| miR-200a-3p | miR-223-5p | miR-302d-5p | miR-3156-5p | miR-329-5p | miR-3654 | miR-374a-3p | miR-3936 | miR-4289 | miR-4434 | miR-449b-5p | miR-4643 |
| miR-200a-5p | miR-22-3p | miR-302e | miR-3157-3p | miR-330-3p | miR-3655 | miR-374b-3p | miR-3937 | miR-429 | miR-4435 | miR-449c-3p | miR-4644 |
| miR-200b-3p | miR-224-3p | miR-302f | miR-3157-5p | miR-330-5p | miR-3657 | miR-374b-5p | miR-3938 | miR-4290 | miR-4436a | miR-449c-5p | miR-4645-3p |
| miR-200b-5p | miR-224-5p | miR-3064-3p | miR-3158-5p | miR-331-3p | miR-3658 | miR-374c-5p | miR-3939 | miR-4291 | miR-4436b-3p | miR-4500 | miR-4646-3p |
| miR-200c-3p | miR-22-5p | miR-3064-5p | miR-3159 | miR-331-5p | miR-3659 | miR-375 | miR-3940-5p | miR-4292 | miR-4437 | miR-4501 | miR-4646-5p |
| miR-200c-5p | miR-2276-3p | miR-30a-3p | miR-31-5p | miR-335-3p | miR-365a-3p | miR-376a-2-5p | miR-3941 | miR-4293 | miR-4438 | miR-4502 | miR-4647 |
| miR-202-3p | miR-2276-5p | miR-30a-5p | miR-3161 | miR-335-5p | miR-365a-5p | miR-376a-3p | miR-3942-5p | miR-4294 | miR-4439 | miR-4503 | miR-4648 |
| miR-202-5p | miR-2278 | miR-30b-3p | miR-3162-5p | miR-337-3p | miR-365b-5p | miR-376a-5p | miR-3943 | miR-4295 | miR-4440 | miR-4504 | miR-4649-3p |
| miR-203b-3p | miR-2355-3p | miR-30b-5p | miR-3163 | miR-337-5p | miR-3660 | miR-376b-3p | miR-3944-5p | miR-4296 | miR-4441 | miR-4505 | miR-4649-5p |
| miR-203b-5p | miR-2355-5p | miR-30c-1-3p | miR-3164 | miR-338-3p | miR-3661 | miR-376b-5p | miR-3945 | miR-4297 | miR-4442 | miR-4506 | miR-4650-3p |
| miR-204-3p | miR-2392 | miR-30c-2-3p | miR-3165 | miR-339-3p | miR-3662 | miR-376c-3p | miR-3973 | miR-4299 | miR-4444 | miR-4507 | miR-4652-3p |
| miR-204-5p | miR-23a-3p | miR-30c-5p | miR-3166 | miR-339-5p | miR-3663-5p | miR-376c-5p | miR-3974 | miR-4300 | miR-4446-3p | miR-4509 | miR-4652-5p |
| miR-2052 | miR-23a-5p | miR-30d-3p | miR-3167 | miR-33a-3p | miR-3664-5p | miR-377-3p | miR-3975 | miR-4301 | miR-4446-5p | miR-450a-1-3p | miR-4653-3p |
| miR-2053 | miR-23b-3p | miR-30d-5p | miR-3169 | miR-33a-5p | miR-3666 | miR-377-5p | miR-3976 | miR-4302 | miR-4447 | miR-450a-2-3p | miR-4653-5p |
| miR-205-3p | miR-23b-5p | miR-30e-3p | miR-3170 | miR-33b-3p | miR-3667-3p | miR-378a-3p | miR-3977 | miR-4303 | miR-4449 | miR-450a-5p | miR-4654 |
| miR-2054 | miR-23c | miR-30e-5p | miR-3171 | miR-33b-5p | miR-3667-5p | miR-378b | miR-3978 | miR-4304 | miR-4450 | miR-450b-3p | miR-4655-3p |
| miR-205-5p | miR-24-2-5p | miR-3115 | miR-3174 | miR-340-3p | miR-3668 | miR-378c | miR-409-3p | miR-4305 | miR-4451 | miR-450b-5p | miR-4655-5p |
| miR-206 | miR-24-3p | miR-3116 | miR-3175 | miR-340-5p | miR-3670 | miR-378d | miR-409-5p | miR-4306 | miR-4452 | miR-4510 | miR-4656 |
| miR-208a-3p | miR-2467-3p | miR-3117-5p | miR-3176 | miR-342-3p | miR-3671 | miR-378e | miR-410-3p | miR-4307 | miR-4453 | miR-4511 | miR-4657 |
| miR-208a-5p | miR-2467-5p | miR-3118 | miR-3177-3p | miR-342-5p | miR-3672 | miR-378f | miR-410-5p | miR-4308 | miR-4455 | miR-4512 | miR-4658 |
| miR-208b-3p | miR-25-3p | miR-3119 | miR-3177-5p | miR-345-3p | miR-367-3p | miR-378g | miR-411-3p | miR-4309 | miR-4456 | miR-4513 | miR-4659a-5p |
| miR-208b-5p | miR-25-5p | miR-3121-5p | miR-3179 | miR-345-5p | miR-3674 | miR-378h | miR-411-5p | miR-4310 | miR-4457 | miR-4514 | miR-4659b-5p |
| miR-20a-3p | miR-2681-3p | miR-3122 | miR-3180 | miR-346 | miR-3675-3p | miR-378i | miR-412-3p | miR-4311 | miR-4458 | miR-4515 | miR-466 |
| miR-20a-5p | miR-2681-5p | miR-3123 | miR-3180-3p | miR-34a-3p | miR-3675-5p | miR-378j | miR-412-5p | miR-4312 | miR-4459 | miR-4517 | miR-4660 |
| miR-20b-3p | miR-2682-3p | miR-3124-3p | miR-3182 | miR-34a-5p | miR-367-5p | miR-379-3p | miR-421 | miR-431-3p | miR-4460 | miR-4518 | miR-4661-5p |
| miR-20b-5p | miR-2682-5p | miR-3124-5p | miR-3184-3p | miR-34b-3p | miR-3677-5p | miR-379-5p | miR-422a | miR-4314 | miR-4461 | miR-4519 | miR-4662a-3p |
| miR-210-3p | miR-26a-1-3p | miR-3125 | miR-3185 | miR-34b-5p | miR-3678-3p | miR-380-3p | miR-424-3p | miR-4315 | miR-4462 | miR-451a | miR-4663 |
| miR-210-5p | miR-26a-2-3p | miR-3126-3p | miR-3186-3p | miR-34c-3p | miR-3679-3p | miR-380-5p | miR-424-5p | miR-431-5p | miR-4463 | miR-451b | miR-4664-3p |
| miR-2110 | miR-26a-5p | miR-3126-5p | miR-3186-5p | miR-34c-5p | miR-3679-5p | miR-381-3p | miR-4251 | miR-4317 | miR-4464 | miR-4520a-5p | miR-4664-5p |
| miR-2113 | miR-26b-3p | miR-3127-3p | miR-3187-3p | miR-3605-3p | miR-3680-3p | miR-381-5p | miR-4252 | miR-4318 | miR-4465 | miR-4521 | miR-4665-5p |
| miR-211-3p | miR-26b-5p | miR-3127-5p | miR-3187-5p | miR-3605-5p | miR-3680-5p | miR-382-3p | miR-4253 | miR-4319 | miR-4467 | miR-4522 | miR-4666a-5p |
| miR-2114-3p | miR-27a-3p | miR-3128 | miR-3188 | miR-3606-3p | miR-3681-5p | miR-382-5p | miR-425-3p | miR-4320 | miR-4468 | miR-452-3p | miR-4666b |
| miR-2114-5p | miR-27a-5p | miR-3129-3p | miR-3189-3p | miR-3606-5p | miR-3682-3p | miR-383-3p | miR-4254 | miR-4322 | miR-4470 | miR-4524a-5p | miR-4667-3p |

#### Human Continued

| | | | | | | | | | | | |
|---|---|---|---|---|---|---|---|---|---|---|---|
| miR-4667-5p | miR-4719 | miR-4770 | miR-495-5p | miR-513a-3p | miR-543 | miR-5581-3p | miR-577 | miR-6132 | miR-6511b-3p | miR-6735-3p | miR-6770-3p |
| miR-4668-3p | miR-4720-3p | miR-4771 | miR-496 | miR-513a-5p | miR-544a | miR-5581-5p | miR-578 | miR-6133 | miR-6513-3p | miR-6735-5p | miR-6770-5p |
| miR-4668-5p | miR-4720-5p | miR-4772-5p | miR-497-3p | miR-513b-3p | miR-544b | miR-5582-3p | miR-579-3p | miR-6134 | miR-6513-5p | miR-6736-3p | miR-6771-3p |
| miR-4669 | miR-4721 | miR-4773 | miR-498 | miR-513b-5p | miR-545-3p | miR-5582-5p | miR-579-5p | miR-614 | miR-651-3p | miR-6736-5p | miR-6771-5p |
| miR-4670-5p | miR-4722-3p | miR-4774-3p | miR-4999-5p | miR-513c-3p | miR-545-5p | miR-5583-5p | miR-580-3p | miR-615-3p | miR-6514-3p | miR-6737-3p | miR-6772-3p |
| miR-4671-3p | miR-4722-5p | miR-4774-5p | miR-499b-3p | miR-513c-5p | miR-548ab | miR-5584-3p | miR-580-5p | miR-616-3p | miR-6514-5p | miR-6737-5p | miR-6772-5p |
| miR-4671-5p | miR-4723-3p | miR-4775 | miR-5000-5p | miR-514a-3p | miR-548ad | miR-5584-5p | miR-581 | miR-6165 | miR-6515-3p | miR-6738-3p | miR-6773-3p |
| miR-4672 | miR-4723-5p | miR-4777-3p | miR-5001-3p | miR-514a-5p | miR-548ae | miR-5585-3p | miR-582-5p | miR-616-5p | miR-6515-5p | miR-6738-5p | miR-6773-5p |
| miR-4673 | miR-4724-3p | miR-4777-5p | miR-5001-5p | miR-514b-3p | miR-548ag | miR-5585-5p | miR-583 | miR-617 | miR-651-5p | miR-6739-3p | miR-6774-3p |
| miR-4675 | miR-4725-3p | miR-4778-3p | miR-5002-3p | miR-514b-5p | miR-548ak | miR-5586-3p | miR-584-3p | miR-618 | miR-6516-3p | miR-6739-5p | miR-6774-5p |
| miR-4676-3p | miR-4725-5p | miR-4779 | miR-5002-5p | miR-515-3p | miR-548al | miR-5586-5p | miR-584-5p | miR-619-3p | miR-6516-5p | miR-6740-3p | miR-6775-3p |
| miR-4677-3p | miR-4726-3p | miR-4780 | miR-5003-3p | miR-515-5p | miR-548an | miR-5587-3p | miR-585-3p | miR-620 | miR-652-3p | miR-6740-5p | miR-6775-5p |
| miR-4677-5p | miR-4726-5p | miR-4781-3p | miR-5003-5p | miR-516a-3p | miR-548ao-3p | miR-5588-5p | miR-586 | miR-621 | miR-652-5p | miR-6741-3p | miR-6776-3p |
| miR-4678 | miR-4727-3p | miR-4781-5p | miR-5004-3p | miR-516a-5p | miR-548ao-5p | miR-5589-3p | miR-587 | miR-622 | miR-653-3p | miR-6741-5p | miR-6776-5p |
| miR-4679 | miR-4727-5p | miR-4782-5p | miR-5004-5p | miR-516b-5p | miR-548ar-3p | miR-5589-5p | miR-588 | miR-623 | miR-654-3p | miR-6742-3p | miR-6778-3p |
| miR-4680-3p | miR-4728-3p | miR-4783-3p | miR-5006-3p | miR-517-5p | miR-548as-3p | miR-559 | miR-589-3p | miR-624-3p | miR-654-5p | miR-6742-5p | miR-6778-5p |
| miR-4680-5p | miR-4728-5p | miR-4784 | miR-5006-5p | miR-517a-3p | miR-548au-3p | miR-5590-5p | miR-589-5p | miR-624-5p | miR-655-3p | miR-6744-3p | miR-6779-3p |
| miR-4681 | miR-4729 | miR-4786-3p | miR-5007-3p | miR-517c-3p | miR-548av-5p | miR-561-3p | miR-590-3p | miR-625-5p | miR-655-5p | miR-6744-5p | miR-6779-5p |
| miR-4682 | miR-4730 | miR-4786-5p | miR-5007-5p | miR-5186 | miR-548aw | miR-561-5p | miR-590-5p | miR-626 | miR-656-3p | miR-6745 | miR-6780a-3p |
| miR-4683 | miR-4731-3p | miR-4788 | miR-5008-5p | miR-5187-3p | miR-548ax | miR-562 | miR-591 | miR-627-3p | miR-656-5p | miR-6746-5p | miR-6780a-5p |
| miR-4684-3p | miR-4731-5p | miR-4789-3p | miR-5009-3p | miR-5187-5p | miR-548ay-3p | miR-563 | miR-592 | miR-628-5p | miR-657 | miR-6747-3p | miR-6780b-3p |
| miR-4684-5p | miR-4732-3p | miR-4789-5p | miR-5009-5p | miR-5188 | miR-548ay-5p | miR-564 | miR-593-3p | miR-629-5p | miR-658 | miR-6747-5p | miR-6780b-5p |
| miR-4685-3p | miR-4732-5p | miR-4790-3p | miR-500a-3p | miR-5189-3p | miR-548az-5p | miR-566 | miR-593-5p | miR-630 | miR-660-3p | miR-6748-3p | miR-6781-3p |
| miR-4685-5p | miR-4733-3p | miR-4790-5p | miR-500a-5p | miR-5189-5p | miR-548b-3p | miR-567 | miR-595 | miR-631 | miR-660-5p | miR-6748-5p | miR-6781-5p |
| miR-4686 | miR-4734 | miR-4791 | miR-500b-3p | miR-518b | miR-548b-5p | miR-568 | miR-597-3p | miR-632 | miR-661 | miR-6750-3p | miR-6782-3p |
| miR-4687-3p | miR-4735-5p | miR-4793-3p | miR-500b-5p | miR-518c-3p | miR-548ba | miR-5680 | miR-597-5p | miR-633 | miR-662 | miR-6750-5p | miR-6782-5p |
| miR-4688 | miR-4736 | miR-4793-5p | miR-5010-3p | miR-5190 | miR-548d-5p | miR-5682 | miR-598-3p | miR-634 | miR-663b | miR-6751-3p | miR-6783-3p |
| miR-4689 | miR-4737 | miR-4794 | miR-5010-5p | miR-5191 | miR-548e-3p | miR-5684 | miR-598-5p | miR-635 | miR-664a-3p | miR-6751-5p | miR-6783-5p |
| miR-4690-3p | miR-4738-3p | miR-4795-3p | miR-5011-5p | miR-5192 | miR-548e-5p | miR-5685 | miR-600 | miR-637 | miR-664a-5p | miR-6752-5p | miR-6784-5p |
| miR-4690-5p | miR-4738-5p | miR-4795-5p | miR-501-3p | miR-5193 | miR-548g-3p | miR-5687 | miR-601 | miR-640 | miR-664b-3p | miR-6753-3p | miR-6785-3p |
| miR-4691-3p | miR-4742-3p | miR-4796-5p | miR-501-5p | miR-5194 | miR-548h-5p | miR-5688 | miR-604 | miR-641 | miR-664b-5p | miR-675-3p | miR-6786-3p |
| miR-4691-5p | miR-4742-5p | miR-4798-3p | miR-502-3p | miR-5195-3p | miR-548j-3p | miR-5689 | miR-605-3p | miR-642a-5p | miR-668-3p | miR-6754-3p | miR-6787-3p |
| miR-4692 | miR-4743-3p | miR-4798-5p | miR-502-5p | miR-5195-5p | miR-548k | miR-569 | miR-605-5p | miR-642b-5p | miR-668-5p | miR-6754-5p | miR-6787-5p |
| miR-4693-3p | miR-4743-5p | miR-4799-5p | miR-503-3p | miR-5196-3p | miR-548l | miR-5690 | miR-606 | miR-643 | miR-670-3p | miR-6755-3p | miR-6788-3p |
| miR-4693-5p | miR-4744 | miR-4800-3p | miR-503-5p | miR-5196-5p | miR-548m | miR-5691 | miR-6069 | miR-644a | miR-670-5p | miR-6755-5p | miR-6789-3p |
| miR-4694-5p | miR-4745-3p | miR-4800-5p | miR-504-3p | miR-5197-3p | miR-548s | miR-5692a | miR-607 | miR-645 | miR-671-3p | miR-675-5p | miR-6789-5p |
| miR-4695-3p | miR-4746-5p | miR-4801 | miR-504-5p | miR-5197-5p | miR-548t-5p | miR-5692b | miR-6071 | miR-646 | miR-6715b-3p | miR-6756-5p | miR-6790-5p |
| miR-4695-5p | miR-4747-3p | miR-4802-3p | miR-5047 | miR-519a-3p | miR-548v | miR-5692c | miR-6072 | miR-647 | miR-671-5p | miR-6757-3p | miR-6791-3p |
| miR-4696 | miR-4747-5p | miR-4802-5p | miR-505-3p | miR-519b-3p | miR-548w | miR-5693 | miR-6073 | miR-648 | miR-6716-3p | miR-6757-5p | miR-6791-5p |
| miR-4697-3p | miR-4748 | miR-4803 | miR-505-5p | miR-519c-3p | miR-548x-3p | miR-5694 | miR-6075 | miR-649 | miR-6716-5p | miR-6758-3p | miR-6792-3p |
| miR-4697-5p | miR-4750-5p | miR-4804-5p | miR-506-3p | miR-519d-3p | miR-548y | miR-5695 | miR-6076 | miR-6499-3p | miR-6717-5p | miR-6758-5p | miR-6792-5p |
| miR-4698 | miR-4751 | miR-484 | miR-506-5p | miR-519e-3p | miR-549a | miR-5696 | miR-6078 | miR-6499-5p | miR-6718-5p | miR-6759-3p | miR-6793-5p |
| miR-4699-3p | miR-4752 | miR-485-3p | miR-507 | miR-519e-5p | miR-550a-3-5p | miR-5697 | miR-6079 | miR-650 | miR-6719-3p | miR-6759-5p | miR-6794-3p |
| miR-4699-5p | miR-4753-3p | miR-485-5p | miR-508-3p | miR-520a-5p | miR-550a-5p | miR-5698 | miR-608 | miR-6500-3p | miR-6720-3p | miR-6760-3p | miR-6794-5p |
| miR-4700-5p | miR-4753-5p | miR-486-5p | miR-508-5p | miR-520b | miR-550b-2-5p | miR-5699-5p | miR-6080 | miR-6500-5p | miR-6720-5p | miR-6760-5p | miR-6795-3p |
| miR-4701-3p | miR-4756-5p | miR-487a-3p | miR-5087 | miR-520c-3p | miR-551a | miR-5700 | miR-6081 | miR-6501-3p | miR-6721-5p | miR-6761-3p | miR-6796-3p |
| miR-4701-5p | miR-4758-5p | miR-487a-5p | miR-5088-3p | miR-520d-3p | miR-551b-3p | miR-5701 | miR-6082 | miR-6501-5p | miR-6722-3p | miR-6761-5p | miR-6796-5p |
| miR-4703-5p | miR-4759 | miR-487b-3p | miR-5088-5p | miR-520e | miR-551b-5p | miR-5702 | miR-6083 | miR-6502-3p | miR-6722-5p | miR-6762-3p | miR-6797-3p |
| miR-4704-3p | miR-4760-3p | miR-487b-5p | miR-5089-3p | miR-520f-3p | miR-552-3p | miR-5703 | miR-6085 | miR-6502-5p | miR-6723-5p | miR-676-3p | miR-6797-5p |
| miR-4704-5p | miR-4760-5p | miR-488-3p | miR-5090 | miR-520g-5p | miR-552-5p | miR-5704 | miR-6086 | miR-6503-3p | miR-6726-3p | miR-6764-3p | miR-6798-5p |
| miR-4705 | miR-4761-3p | miR-488-5p | miR-5091 | miR-521 | miR-553 | miR-5705 | miR-6088 | miR-6503-5p | miR-6726-5p | miR-6764-5p | miR-6799-3p |
| miR-4706 | miR-4761-5p | miR-489-3p | miR-5093 | miR-522-3p | miR-554 | miR-5706 | miR-609 | miR-6504-5p | miR-6727-5p | miR-6765-5p | miR-6799-5p |
| miR-4707-3p | miR-4762-5p | miR-489-5p | miR-509-3-5p | miR-523-3p | miR-555 | miR-5707 | miR-610 | miR-6505-3p | miR-6728-3p | miR-676-5p | miR-6800-3p |
| miR-4707-5p | miR-4763-3p | miR-490-5p | miR-509-3p | miR-525-5p | miR-556-3p | miR-571 | miR-612 | miR-6505-5p | miR-6728-5p | miR-6766-3p | miR-6800-5p |
| miR-4709-3p | miR-4764-3p | miR-491-3p | miR-5094 | miR-532-3p | miR-556-5p | miR-572 | miR-6124 | miR-6506-3p | miR-6729-3p | miR-6766-5p | miR-6801-3p |
| miR-4709-5p | miR-4764-5p | miR-491-5p | miR-509-5p | miR-532-5p | miR-557 | miR-573 | miR-6126 | miR-6506-5p | miR-6730-5p | miR-6767-3p | miR-6801-5p |
| miR-4711-3p | miR-4765 | miR-492 | miR-510-3p | miR-539-3p | miR-5571-3p | miR-5739 | miR-6127 | miR-6507-5p | miR-6731-3p | miR-6767-5p | miR-6802-3p |
| miR-4712-5p | miR-4766-3p | miR-493-3p | miR-510-5p | miR-539-5p | miR-5571-5p | miR-574-3p | miR-6128 | miR-6508-5p | miR-6731-5p | miR-6768-3p | miR-6802-5p |
| miR-4713-3p | miR-4766-5p | miR-493-5p | miR-511-3p | miR-541-3p | miR-5572 | miR-574-5p | miR-6129 | miR-6509-5p | miR-6733-3p | miR-6768-5p | miR-6803-3p |
| miR-4713-5p | miR-4768-5p | miR-494-3p | miR-511-5p | miR-541-5p | miR-5579-5p | miR-575 | miR-613 | miR-6510-3p | miR-6733-5p | miR-6769a-3p | miR-6803-5p |
| miR-4714-5p | miR-4769-3p | miR-494-5p | miR-512-3p | miR-542-3p | miR-558 | miR-576-3p | miR-6130 | miR-6511a-3p | miR-6734-3p | miR-6769a-5p | miR-6804-3p |
| miR-4718 | miR-4769-5p | miR-495-3p | miR-512-5p | miR-542-5p | miR-5580-3p | miR-576-5p | miR-6131 | miR-6511a-5p | miR-6734-5p | miR-6769b-3p | miR-6805-5p |

#### Human Continued

| | | | | | | | | | | | |
|---|---|---|---|---|---|---|---|---|---|---|---|
| miR-6806-3p | miR-6821-5p | miR-6837-3p | miR-6854-3p | miR-6872-3p | miR-6888-3p | miR-7114-3p | miR-744-3p | miR-7848-3p | miR-8069 | miR-877-5p | miR-93-3p |
| miR-6806-5p | miR-6822-3p | miR-6837-5p | miR-6854-5p | miR-6872-5p | miR-6888-5p | miR-7114-5p | miR-744-5p | miR-7850-5p | miR-8070 | miR-885-3p | miR-934 |
| miR-6807-3p | miR-6822-5p | miR-6838-3p | miR-6855-3p | miR-6873-3p | miR-6889-3p | miR-7-1-3p | miR-7515 | miR-7851-3p | miR-8071 | miR-885-5p | miR-935 |
| miR-6807-5p | miR-6823-3p | miR-6839-5p | miR-6855-5p | miR-6873-5p | miR-6889-5p | miR-7150 | miR-758-3p | miR-7852-3p | miR-8074 | miR-887-3p | miR-93-5p |
| miR-6808-5p | miR-6823-5p | miR-6840-3p | miR-6856-3p | miR-6874-3p | miR-6890-3p | miR-7151-3p | miR-758-5p | miR-7853-5p | miR-8076 | miR-887-5p | miR-936 |
| miR-6809-3p | miR-6824-5p | miR-6840-5p | miR-6856-5p | miR-6874-5p | miR-6890-5p | miR-7151-5p | miR-759 | miR-7854-3p | miR-8077 | miR-888-3p | miR-937-3p |
| miR-6809-5p | miR-6825-3p | miR-6841-3p | miR-6857-3p | miR-6875-5p | miR-6891-3p | miR-7152-3p | miR-7-5p | miR-7856-5p | miR-8078 | miR-888-5p | miR-937-5p |
| miR-6810-3p | miR-6825-5p | miR-6841-5p | miR-6857-5p | miR-6876-3p | miR-6892-3p | miR-7152-5p | miR-760 | miR-7973 | miR-8079 | miR-889-3p | miR-938 |
| miR-6810-5p | miR-6826-5p | miR-6842-3p | miR-6859-3p | miR-6877-3p | miR-6892-5p | miR-7153-3p | miR-761 | miR-7974 | miR-8080 | miR-889-5p | miR-939-5p |
| miR-6811-3p | miR-6827-3p | miR-6842-5p | miR-6859-5p | miR-6877-5p | miR-6893-3p | miR-7153-5p | miR-764 | miR-7976 | miR-8081 | miR-890 | miR-9-3p |
| miR-6811-5p | miR-6827-5p | miR-6843-3p | miR-6861-3p | miR-6878-3p | miR-6893-5p | miR-7154-3p | miR-765 | miR-7978 | miR-8082 | miR-891a-3p | miR-941 |
| miR-6812-5p | miR-6828-3p | miR-6844 | miR-6862-3p | miR-6878-5p | miR-6894-5p | miR-7154-5p | miR-766-3p | miR-802 | miR-8083 | miR-891a-5p | miR-942-3p |
| miR-6813-3p | miR-6828-5p | miR-6845-5p | miR-6862-5p | miR-6880-5p | miR-6895-3p | miR-7155-5p | miR-766-5p | miR-8052 | miR-8084 | miR-891b | miR-943 |
| miR-6813-5p | miR-6829-5p | miR-6846-3p | miR-6863 | miR-6881-3p | miR-6895-5p | miR-7156-3p | miR-767-3p | miR-8053 | miR-8085 | miR-892a | miR-944 |
| miR-6814-3p | miR-6830-3p | miR-6846-5p | miR-6864-3p | miR-6881-5p | miR-708-5p | miR-7156-5p | miR-767-5p | miR-8054 | miR-8086 | miR-892b | miR-95-3p |
| miR-6814-5p | miR-6830-5p | miR-6847-3p | miR-6864-5p | miR-6882-3p | miR-7106-3p | miR-7157-3p | miR-769-3p | miR-8055 | miR-8087 | miR-892c-3p | miR-95-5p |
| miR-6815-3p | miR-6831-3p | miR-6847-5p | miR-6865-5p | miR-6882-5p | miR-7106-5p | miR-7157-5p | miR-769-5p | miR-8056 | miR-8088 | miR-892c-5p | miR-9-5p |
| miR-6815-5p | miR-6831-5p | miR-6848-3p | miR-6866-3p | miR-6883-3p | miR-7107-3p | miR-7158-3p | miR-7702 | miR-8057 | miR-8089 | miR-920 | miR-96-3p |
| miR-6816-3p | miR-6832-3p | miR-6848-5p | miR-6866-5p | miR-6883-5p | miR-7107-5p | miR-7159-3p | miR-7703 | miR-8058 | miR-873-3p | miR-921 | miR-96-5p |
| miR-6817-3p | miR-6832-5p | miR-6849-3p | miR-6867-3p | miR-6884-3p | miR-7108-5p | miR-7159-5p | miR-7705 | miR-8059 | miR-873-5p | miR-922 | miR-98-3p |
| miR-6817-5p | miR-6833-3p | miR-6849-5p | miR-6867-5p | miR-6884-5p | miR-7109-3p | miR-7160-3p | miR-770-5p | miR-8060 | miR-874-3p | miR-924 | miR-99a-5p |
| miR-6818-3p | miR-6833-5p | miR-6851-3p | miR-6868-3p | miR-6885-3p | miR-7109-5p | miR-7160-5p | miR-7706 | miR-8061 | miR-874-5p | miR-92a-1-5p | miR-99b-3p |
| miR-6818-5p | miR-6834-3p | miR-6851-5p | miR-6868-5p | miR-6885-5p | miR-711 | miR-7161-3p | miR-7843-3p | miR-8062 | miR-875-3p | miR-92a-2-5p | miR-99b-5p |
| miR-6819-3p | miR-6834-5p | miR-6852-3p | miR-6869-5p | miR-6886-3p | miR-7111-3p | miR-7161-5p | miR-7843-5p | miR-8063 | miR-875-5p | miR-92a-3p | |
| miR-6819-5p | miR-6835-3p | miR-6852-5p | miR-6870-3p | miR-6886-5p | miR-7111-5p | miR-7162-3p | miR-7844-5p | miR-8065 | miR-876-3p | miR-92b-3p | |
| miR-6820-3p | miR-6835-5p | miR-6853-3p | miR-6870-5p | miR-6887-3p | miR-7112-3p | miR-7162-5p | miR-7845-5p | miR-8067 | miR-876-5p | miR-92b-5p | |
| miR-6821-3p | miR-6836-5p | miR-6853-5p | miR-6871-5p | miR-6887-5p | miR-7113-5p | miR-7-2-3p | miR-7846-3p | miR-8068 | miR-877-3p | miR-933 | |

**HTG Molecular Diagnostics, Inc.**
Call 1-877-289-2615
www.htgmolecular.com

**For Research Use Only. Not for use in diagnostic procedures.**
All trademarks are the property of HTG Molecular and its subsidiaries
unless otherwise specified.
