## Supplementary Table 5 for "Multi-omics analysis reveals drivers of loss of β-cell function after newly diagnosed autoimmune type 1 diabetes: An INNODIA^‡^ multicenter study"

**Supplementary Table 5** Internal standards used in lipidomics analysis.

| Name | Abbreviation |
|---|---|
| 1,2-diheptadecanoyl-sn-glycero-3-phosphoethanolamine | PE(17:0/17:0) |
| N-heptadecanoyl-D-erythro-sphingosylphosphorylcholine | SM(d18:1/17:0) |
| N-heptadecanoyl-D-erythro-sphingosine | Cer(d18:1/17:0) |
| 1,2-diheptadecanoyl-sn-glycero-3-phosphocholine | PC(17:0/17:0) |
| 1-heptadecanoyl-2-hydroxy-sn-glycero-3-phosphocholine | LPC(17:0) |
| 1-palmitoyl-d31-2-oleoyl-sn-glycero-3-phosphocholine | PC(16:0/d31/18:1) |
| 1,2-Dimyristoyl-sn-glycero-3-phospho(choline-d13) | PC(14:0/d13) |
| Tripalmitin-1,1,1-13C3 | TG(16:0/16:0/16:0)-13C3 |
| Trioctanoin-1,1,1-13C3 | TG(8:0/8:0/8:0)-13C3 |
