## Supplementary Table 6 for "Multi-omics analysis reveals drivers of loss of β-cell function after newly diagnosed autoimmune type 1 diabetes: An INNODIA^‡^ multicenter study"

**Supplementary Table 6** Multi-FACS flow panel antibodies and reagents

| Steps/mastermix | Marker/reagent | Fluorochome | Clone |
|---|---|---|---|
| **Viability stain** | Live/dead blue | Live/dead blue | N/A |
| **Block Fc receptor** | Human TruStain FcX (Fc Receptor Blocking Solution) | N/A | N/A |
| **Mastermix 1** | Brilliant stain buffer plus | N/A | N/A |
| | CD183/CXCR3 | PE-CY5 | 1C6/CXCR3 |
| | CD117 | BUV805 | 104D2 |
| | CD294/CRTH2 | BUV615 | BM16 |
| | CD161 | eFluor450 | HP-3G10 |
| **Mastermix 2** | Brilliant stain buffer plus | N/A | N/A |
| | CD185/CXCR5 | Brilliant Violet 750 | RF8B2 |
| | ICOS | BB515 | DX29 |
| | CCR7 | Brilliant Violet 785 | G043H7 |
| | CD196/CCR6 | Brilliant Violet 711 | G034E3 |
| **Mastermix 3** | Brilliant stain buffer plus | N/A | N/A |
| | CD16 | PerCP-Cy5.5 | 3G8 |
| | CD20 | Pacific Orange | HI47 |
| | CD95 | Brilliant Violet 650 | DX2 |
| | CD27 | APC | O323 |
| | CD38 | APC-eFluor780 | HIT2 |
| | CD24 | PE/Dazzle™ 594 | ML5 |
| | TIGIT | PE-Cy7 | A15153G |
| | CD25 | PE-Af700 | CD25-3G10 |
| | TCRgd | PerCP-eFluor710 | B1.1 |
| | CD127 | APC-R700 | HIL-7R-M21 |
| | CD45 | PerCP | H130 |
| | CD123 | Super Bright 436 | 6H6 |
| | KLRG1 | Alexa Fluor® 647 | SA231A2 |
| | CD4 | CF568 | C4/206 |
| | CD19 | Spark NIR 685 | HIB19 |
| | CD8 | Qdot 800 | 3B5 |
| | CD57 | FITC | HNK-1 |
| | CD14 | Spark blue 550 | 63D3 |
| | CD39 | BUV496 | TU66 |
| | CD28 | BUV563 | CD28.2 |
| | PD-1 | Brilliant Violet 421 | EH12.2H7 |
| | HLA-DR | Brilliant Violet 570 | L243 |
| | CD3 | Brilliant Violet 510 | OKT3 |
| | CD56 | BUV737 | NCAM16.2 |
| | CD31 | Brilliant Violet 605 | WM59 |
| | CD11c | BUV661 | B-ly6 |
| | IgD | Brilliant Violet 480 | IA6-2 |
| | CD45RA | BUV395 | 5H9 |
